# Intrapartum antibiotic prophylaxis to prevent Group B streptococcal infections in newborn infants: a systematic review and meta-analysis comparing various strategies

**DOI:** 10.1101/2024.06.17.24309017

**Authors:** Timothy J.R. Panneflek, Gea F. Hasperhoven, Yamikani Chimwaza, Connor Allen, Tina Lavin, Arjan B. te Pas, Vincent Bekker, Thomas van den Akker

## Abstract

**Background:** Early-onset Group B Streptococcus (EOGBS) leads to substantial morbidity and mortality in newborn infants. Intrapartum antibiotic prophylaxis (IAP) prevents EOGBS infection, but IAP strategies vary. The approach to the provision of IAP can be risk-based, universal or a combination of the two strategies. Previous systematic reviews reported that universal strategies might be most optimal in lowering EOGBS infection, but there is no consensus. Therefore, we aimed to provide up-to-date evidence on effectiveness of different strategies by comparing perinatal outcomes.

**Methods:** A systematic search for EOGBS prevention strategies was performed in MEDLINE, Embase and Web of Science. Studies were included if they reported on different strategies and outcomes of interest, including EOGBS infection, IAP administration and antimicrobial resistance. Summary data was extracted from published reports. Study quality was assessed using the ROBINS-I tool. Random-effects meta-analyses were used to determine risk ratios (RR) and 95%-confidence intervals. PROSPERO registration CRD42023411806.

**Findings:** A total of 6050 records were identified, of which 72 observational studies were included for synthesis with more than 10 million live births. Meta-analysis demonstrated that implementation of any strategy (n=34 studies, RR 0.46 (0.36-0.60)), risk-based strategies (n=11 studies, RR 0.65 (0.48-0.87)), or universal strategies (n=16 studies, RR 0.37 (0.25-0.55)) was associated with a reduced risk of EOGBS infection compared to no strategy. In direct comparison, universal strategies were associated with a reduced risk of EOGBS compared to a risk-based strategy (n=17 studies, RR 0.41 (0.30-0.55)), while the proportion of women receiving IAP did not differ between risk-based (16%) and universal (21%) strategies (n=9 studies, RR 1.29 (0.95-1.75)). There was no antimicrobial resistance of EOGBS isolates to penicillin or ampicillin (n=11 studies).

**Interpretation:** Any IAP strategy could reduce the risk of EOGBS infection without evidence of increasing antimicrobial resistance. Universal strategies give the largest reduction in the EOGBS burden, while not exposing a significantly higher proportion of pregnancies to IAP compared to risk-based strategies.

**Funding:** UNDP-UNFPA-UNICEF-WHO-World Bank Special Programme of Research, Development and Research Training in Human Reproduction, a cosponsored programme executed by the World Health Organisation.

## Introduction

Early-onset Group B Streptococcus (EOGBS) infection, comprising sepsis, pneumonia and meningitis, are a leading cause of neonatal morbidity and mortality, with a worldwide incidence of 0.41 per 1000 live births and a mortality rate of 4-10% in high-income countries.^1,2^ EOGBS infections are defined as the presence of Group B Streptococcus (GBS) within 7 days of birth in normally sterile fluids, such as blood and cerebrospinal fluid.^3,4^ Infants may be infected antenatally by vertical transmission in colonised pregnant women.^5^ Globally, GBS colonisation of the recto-vaginal tract is 18% among all pregnant women at any point during pregnancy.^6^ Therefore, prevention of the sequelae of GBS colonisation contributes importantly to perinatal health.

Currently, prevention of vertical transmission consists of administration of intrapartum antibiotic prophylaxis (IAP) at least 4 hours before birth, which is associated with a reduced risk of EOGBS infection.^7,8^ IAP is administered according to various screening strategies applied in different settings. Risk-factor based screening (‘risk-based’) strategies are used in some settings, where IAP is administered according to the presence of any risk factor for EOGBS during pregnancy.^9^ In universal microbiology-based screening (‘universal’) strategies GBS colonisation is determined in all pregnant women antenatally and IAP is administered to women with GBS colonisation.^7^ In addition, IAP can be administered with any combination of elements from risk-based and universal strategies (‘other’).^10,11^

Previous systematic reviews and meta-analyses reported that universal strategies were associated with a lower incidence of EOGBS infection compared to risk-based strategies or having no strategy.^12,13^ However, IAP strategies still vary considerably around the world, which is partly due to discussion concerning increased antibiotic use, antimicrobial resistance development and risk of a possible increase in non-GBS early-onset sepsis (EOS), caused by pathogens like *E. coli*.^14–16^ Moreover, even when implementing universal strategies, there is no consensus of timing of GBS determination.

In order to contribute to consensus on GBS prevention strategies, this review elaborates on previous work, while incorporating up-to-date evidence on a variety of outcomes needed for evidence-based and nuanced policy-making. We evaluated the effectiveness of different prevention strategies, by comparing maternal and neonatal infectious morbidity and mortality, the frequency of IAP administration, and the presence of antimicrobial resistance. Furthermore, we evaluated timing of GBS determination in universal strategies to prevent EOGBS infection in newborns.

## Methods

### Protocol and Registration

The Preferred Reporting Items for Systematic Reviews and Meta-Analyses (PRISMA) guidelines and Cochrane Handbook for Systematic Reviews were used to conduct and report this systematic review and meta-analysis.^17,18^ The protocol was registered in PROSPERO, with ID CRD42023411806.^19^

### Eligibility criteria

#### Study designs

We included any randomised and non-randomised study with human participants that reported on any of the outcomes of interest (specified below) when comparing/describing at least two different strategies for IAP administration to prevent EOGBS infections, including no strategy. Non-randomised studies included non-randomised interventional studies (e.g. quasi-randomised controlled trials) and uncontrolled observational studies (e.g. cohort studies). Studies were eligible regardless of sample size, year, country or language of publication and temporal data collection.

Studies were excluded when:

- only qualitative data was presented;
- strategies coincided;
- data was already published in other articles^1^;
- data was based on models, not actual cases;
- the full-text article could not be obtained, or;
- the article was categorised as case report, case series, letter to the editor, commentary, or conference abstract.

#### Participants

Participants were pregnant women and neonates.

#### Interventions

The intervention consisted of strategies for IAP administration (including no strategy). Risk-based strategies were defined as strategies where IAP was administered according to the presence of any risk factor for EOGBS during pregnancy, such as a history of an infant with EOGBS infection, presence of GBS bacteriuria during pregnancy, or presence of intrapartum fever, preterm labour or prolonged rupture of membranes >18 hours.^9^ Universal strategies were defined as strategies where GBS colonisation was determined using microbiological testing and IAP was administered to all women positive for GBS colonisation.^7^ Other strategies were defined as strategies where IAP was administered based any combination of elements from risk-based and universal strategies. ‘Other’ strategies usually consisted of strategies where both risk-based and universal strategies were implemented in parallel and selective strategies that only treat pregnant women positive for GBS colonisation with the presence of least one risk factor.^10,11^ Lastly, IAP was sometimes found to be administered without any official screening (‘no’) strategy on an individual basis and at the discretion of the attending physician.^12,20^

#### Outcome measures

Review outcomes were selected based on the critical and important outcomes used in the WHO recommendations on the prevention and treatment of maternal peripartum infections.^21^ Outcomes listed below were used as primary outcomes for the review. Protocol outcomes

were specified into the following outcomes:

- Incidence of EOGBS infection per 1000 live births or pregnant women (as defined by study authors).
- Incidence of non-GBS early-onset sepsis (EOS) per 1000 live births or pregnant women.
- Incidence of all EOS per 1000 live births or pregnant women.
- EOGBS-related mortality rate per 1000 live births or pregnant women.
- IAP administration (%).
- Antimicrobial resistance in EOS isolates (%).
- Maternal peripartum infection (%). (<u>Supplementary file 2</u>)

#### Information sources

Records were obtained through a systematic search of MEDLINE, Embase, and Web of Science databases. Additional publications were obtained manually by searching reference lists of relevant records and reviews.

#### Search strategy

The full search strategy used in MEDLINE is presented in <u>Supplementary file 1</u>. Search terms used included pregnancy, newborn, infant, Group B streptococcus, screening, culture-based, polymerase chain reaction, risk-based, prevention, guidelines, and early onset. In the final search, articles (regardless of language) were included until 2024. The last queries were run in May 2024.

#### Selection process

Potential studies were identified from the search strategy by double (independent) review (TJRP, GFH, TL, CA, YC, VB). Any disagreements were resolved by a third author (TJRP, GFH, TL, CA, YC, VB, respectively) and reviewers were not blinded. All titles and abstracts were placed in EndNote version X20 (Clarivate Analytics, Philadelphia, PA, USA) to automatically and manually deduplicate the list of studies. Afterwards, all articles were screened for eligibility via title and abstract in COVIDENCE (Veritas Health Innovation, Melbourne, Australia). Selected studies were subject to full-text screening. Full-text screening was done using aforementioned selection criteria.

#### Data collection process

Two authors (TJRP, GFH) independently extracted the following data: study design, study population and outcome measures using a preformed data extraction sheet.

#### Data items

Data items consisted of general information about the article, on the study population, IAP strategies, and outcome information. Outcome measures were manually calculated using raw data from the article. Missing data were reported as ‘No data’ in the tables.

#### Study risk of bias assessment

Risk of bias was assessed via the Risk of Bias in Non-randomised Studies of Interventions (ROBINS-I) tool.^22^ Studies were assessed by two independent reviewers (TJRP, GFH), and any discrepancies were handled through discussion until consensus. Seven domains of bias were assessed: confounding, selection of participants into the study, classification of interventions, deviations from intended intervention, missing data, measurement of outcomes and selection of the reported results. Each domain was scored as low, moderate, serious, critical, or unknown risk of bias. Confounding factors relevant to this review were specified before the studies were assessed for risk of bias.

#### Effect measures

EOGBS infection, non-GBS EOS and all EOS incidences were presented as cases per 1000 live births or pregnant women. EOGBS-related mortality rate was presented as deaths per 1000 live births or pregnant women. IAP administration incidence was presented as the percentage of pregnant women receiving IAP. Antimicrobial resistance incidence was presented as the percentage of resistance with respect to all EOS isolates examined. Incidence rates were calculated using the data provided in the studies.

Incidence of all early-onset infections and administration of IAP were compared between strategies with pooled risk ratios (RR) and 95%-Confidence intervals (CIs). EOGBS-related mortality rate (per 1000 live births or pregnant women) and IAP administration (%) incidence were also reported as a pooled incidence of the random-effects meta-analyses with a random intercept logistic regression model. Individual study 95%-CIs were reported with normal approximation confidence intervals. Meta-regression output was presented as β-coefficient and corresponding p-value.

#### Synthesis methods

Studies were only included in data synthesis if they reported on at least two different strategies (including no strategy) and if raw data was provided on the incidence of the outcomes of interest. The category ‘other strategies’ was not included in the meta-analysis due to heterogeneity in the strategies used, but these studies were included in the ‘any’ vs. ‘no’ strategy comparison. We used R version 4.2.1 (R: The R Project for Statistical Computing (r-project.org)) within Studio version 4.2.1 (R Studio, Boston, MA, USA, 2022) to combine studies, synthesize data, and create funnel plots. Heterogeneity was assessed by i) a Chi^2^-test for variation between studies and ii) the I^2^ statistic, which described the proportion of variation that is due to heterogeneity.

As we anticipated marked heterogeneity between studies that might influence treatment effect, in terms of study population, baseline infection incidence and timing of GBS determination, we performed subgroup analyses in addition to random-effects meta-regression. Subgroup analyses included comparisons in EOGBS infection incidence in studies that reported on term case incidence. Meta-regression analyses were performed post-hoc with an inverse variance method for all EOGBS comparisons that included at least 10 studies. We sought to delineate baseline EOGBS infection incidence (in cases per 1000 live births or pregnant women) and timing of GBS determination. Meta-regression analyses for timing of GBS determination were performed with two different methods in two different comparisons. In the comparison between universal and no strategy, the timing of GBS determination was compared between early determination before 33 weeks’ gestation and late determination at 33-37 weeks’ gestation. In the comparison between universal and risk-based strategies, timing of determination was compared between (antepartum) determination at 35-37 weeks’ gestation and (intrapartum) GBS colonisation determined within two days of birth.

We performed sensitivity analyses to assess the robustness of the synthesised results by excluding all studies with a serious risk of bias in the comparisons for EOGBS infection incidence. A p-value <0.05 was considered significant.

#### Reporting bias assessment

Publication bias analyses were carried out via visual inspection of the funnel plots and an Egger’s test for funnel plot asymmetry for meta-analyses, including at least ten studies.^23^

### Certainty assessment

Certainty of the outcomes was assessed using the GRADE approach of the Cochrane Handbook for Systematic Reviews of Interventions.^24^ We critically evaluated study limitations, consistency of effect, imprecision, indirectness, and publication bias in the outcomes. An overall judgement of certainty assessment ranged from very low to high certainty evidence. Study limitations were evaluated with degree of risk of bias. Inconsistency was based on I^2^ and overlap of 95%-CI estimates. Indirectness was not relevant to this review. Imprecision was based on 95%-CI estimate of outcome. Publication bias was assessed as listed above.

## Results

### Study selection

The selection process is illustrated in a flow diagram in Figure 1. The three databases overall provided 11,025 records, which were reduced to 6,613 after removing duplicates. Title and abstract screening was done in all 6,613 records: 5,624 did not meet inclusion criteria, and the remaining 669 articles were reviewed in full-text. Ultimately, data on 82 articles^25–106^ were presented and 72 articles^25–96^ were included in the synthesis of the systematic review. Reasons for exclusions are listed in Figure 1. No randomised controlled trials on this subject were found.

**Figure 1:**
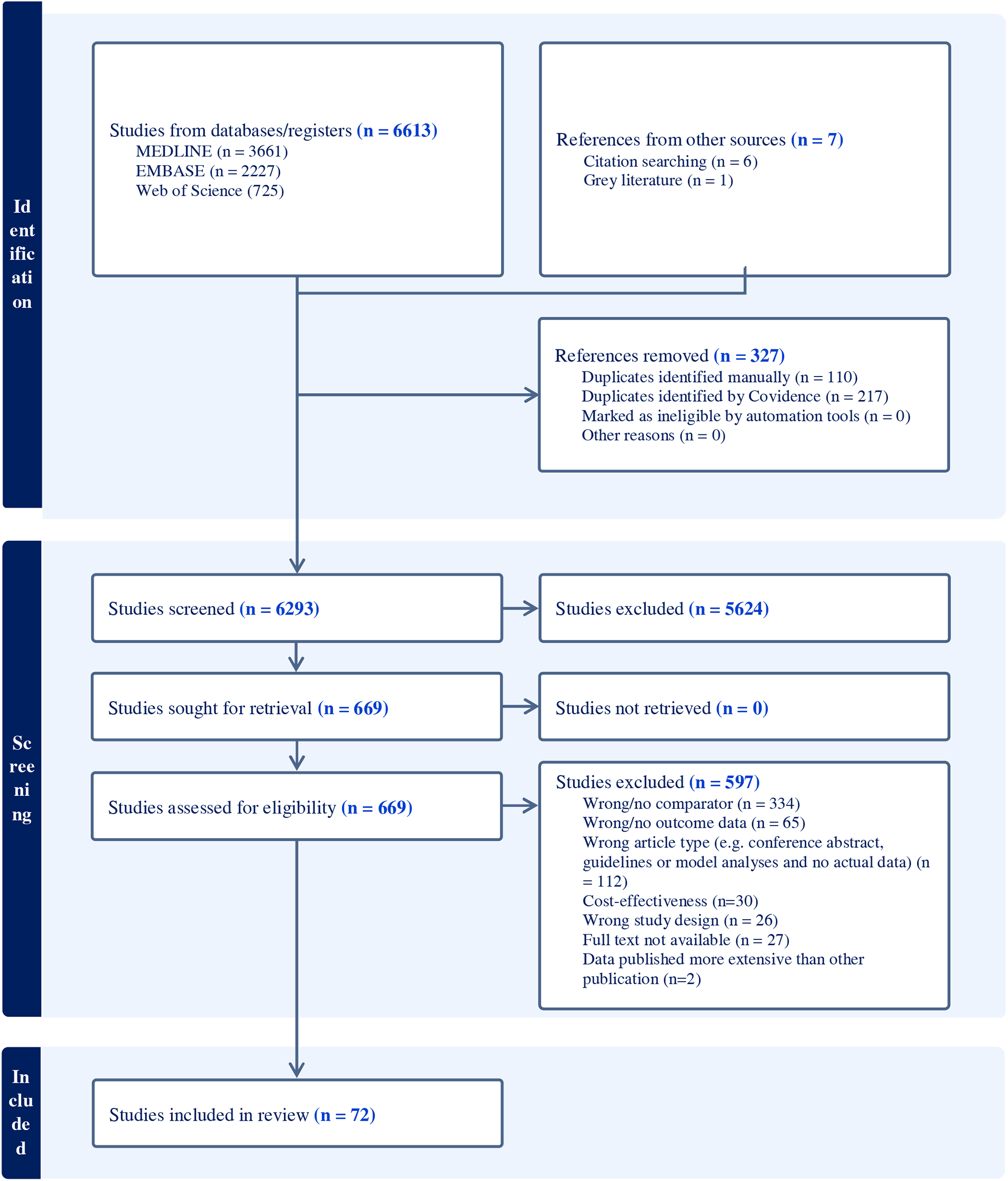
PRISMA study inclusion flowchart PRISMA flowchart of retrieved and included studies.

### Study characteristics

Overall, the 72 included studies included more than 10 million live births/deliveries from 20 different countries (Table 1). A total of 55 observational studies reported on EOGBS infection incidence, 18 on non-GBS EOS, 19 on all EOS, 2 on timing of GBS determination, 19 on EOGBS-related mortality, 16 on IAP administration and 16 on antimicrobial resistance. Two studies reported on timing of GBS determination. Study characteristics and results of individual studies are displayed in Table 1 and <u>Supplementary file 2</u>.

**Table 1:**
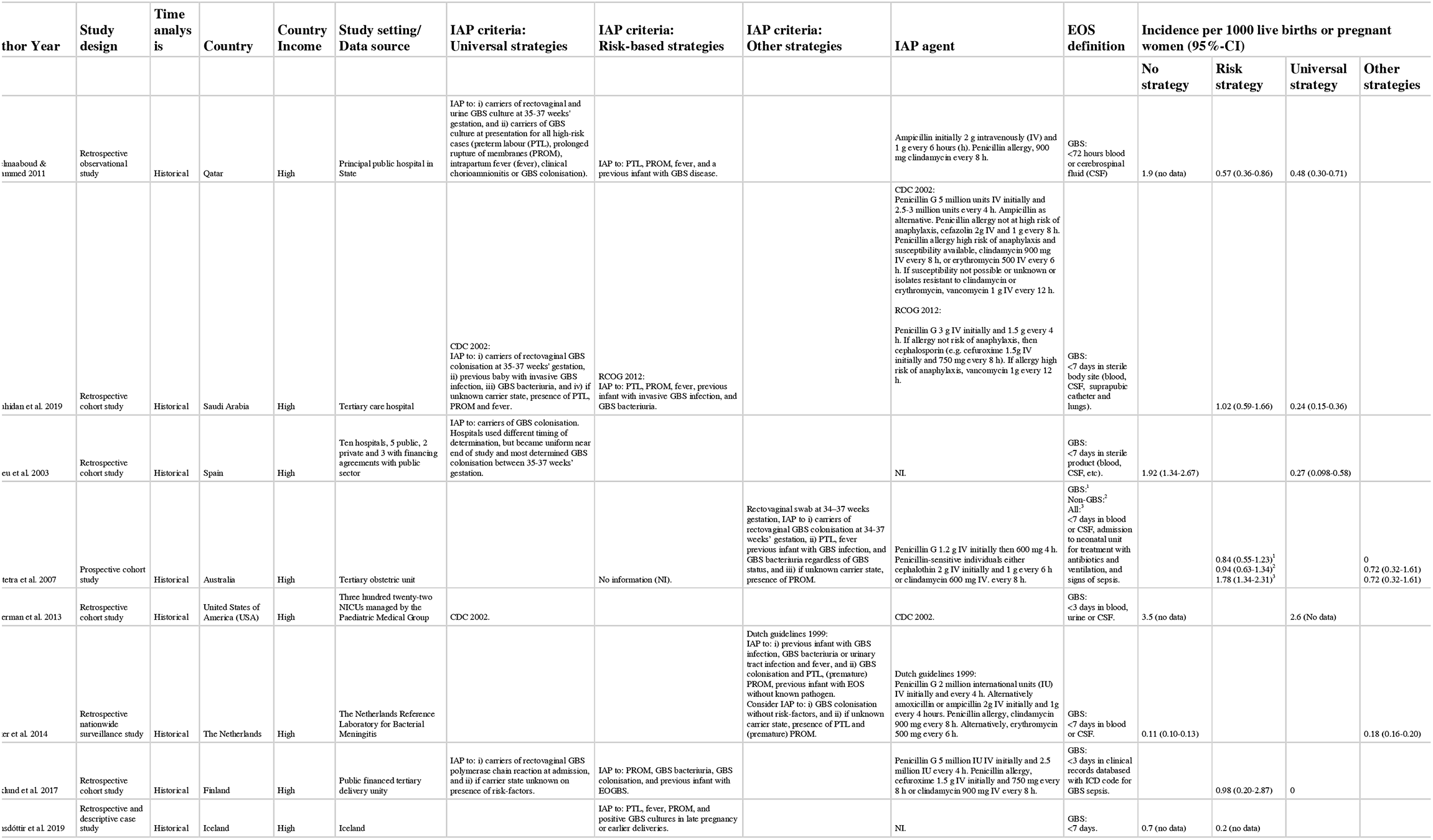
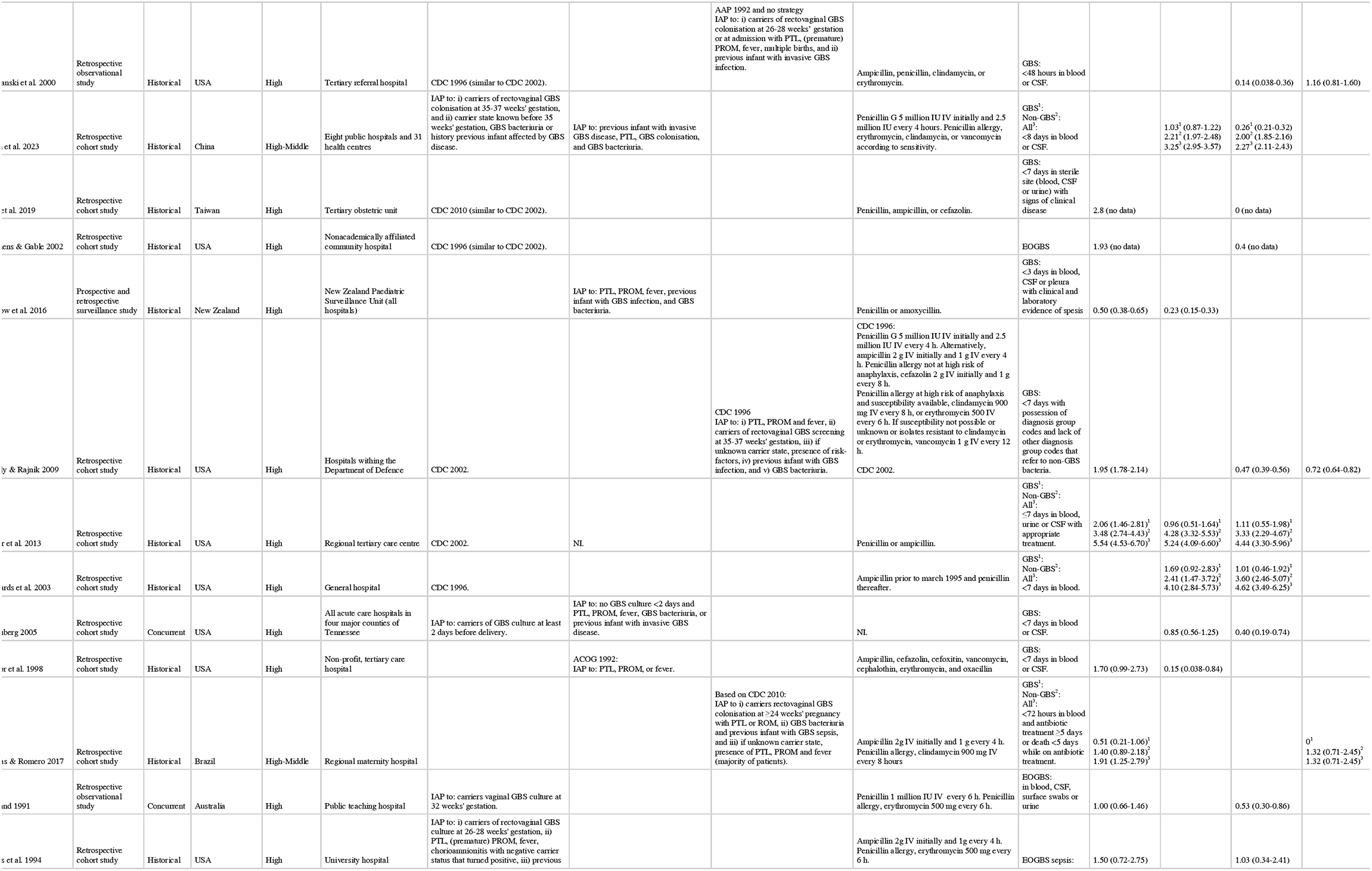
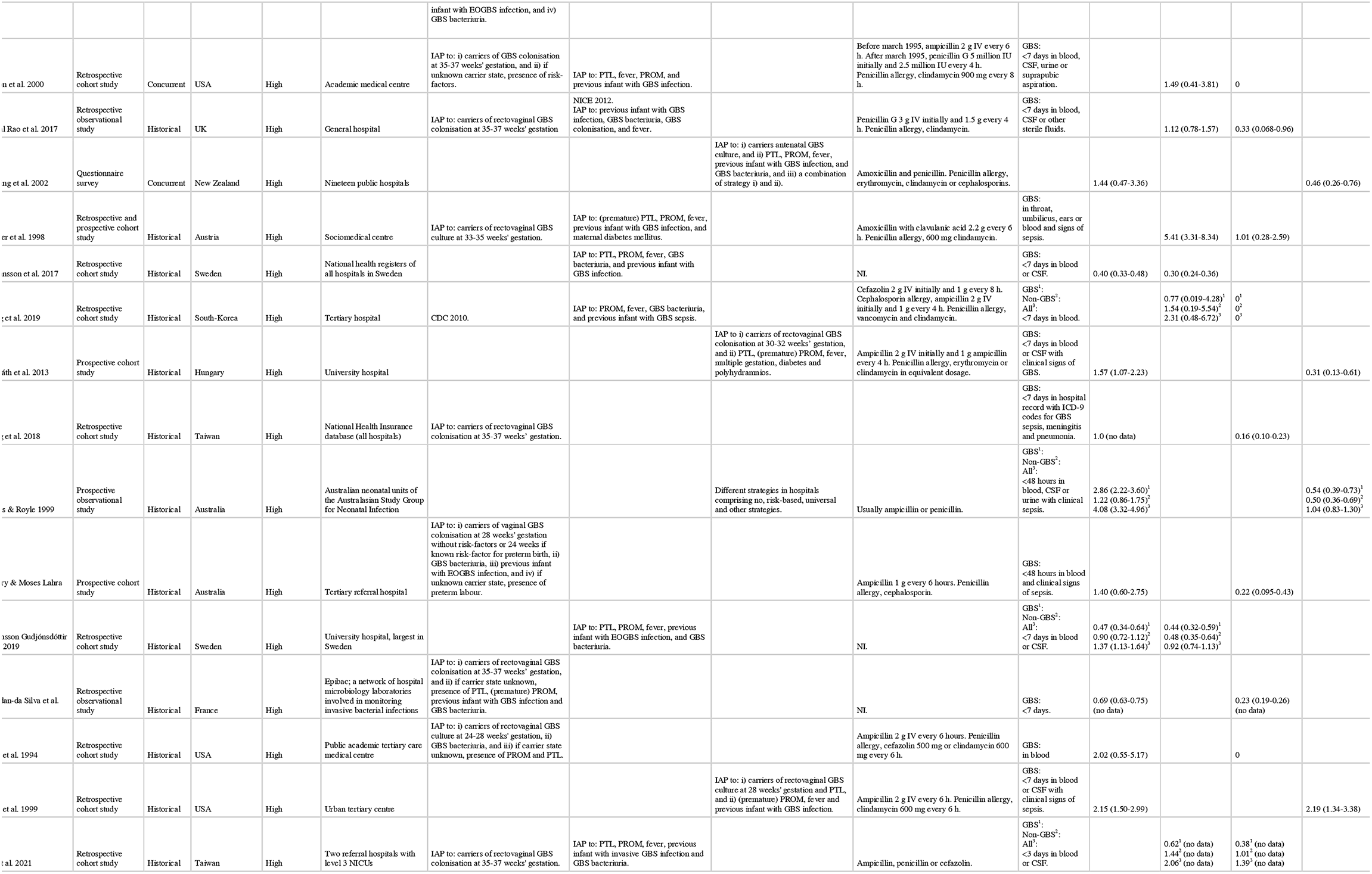
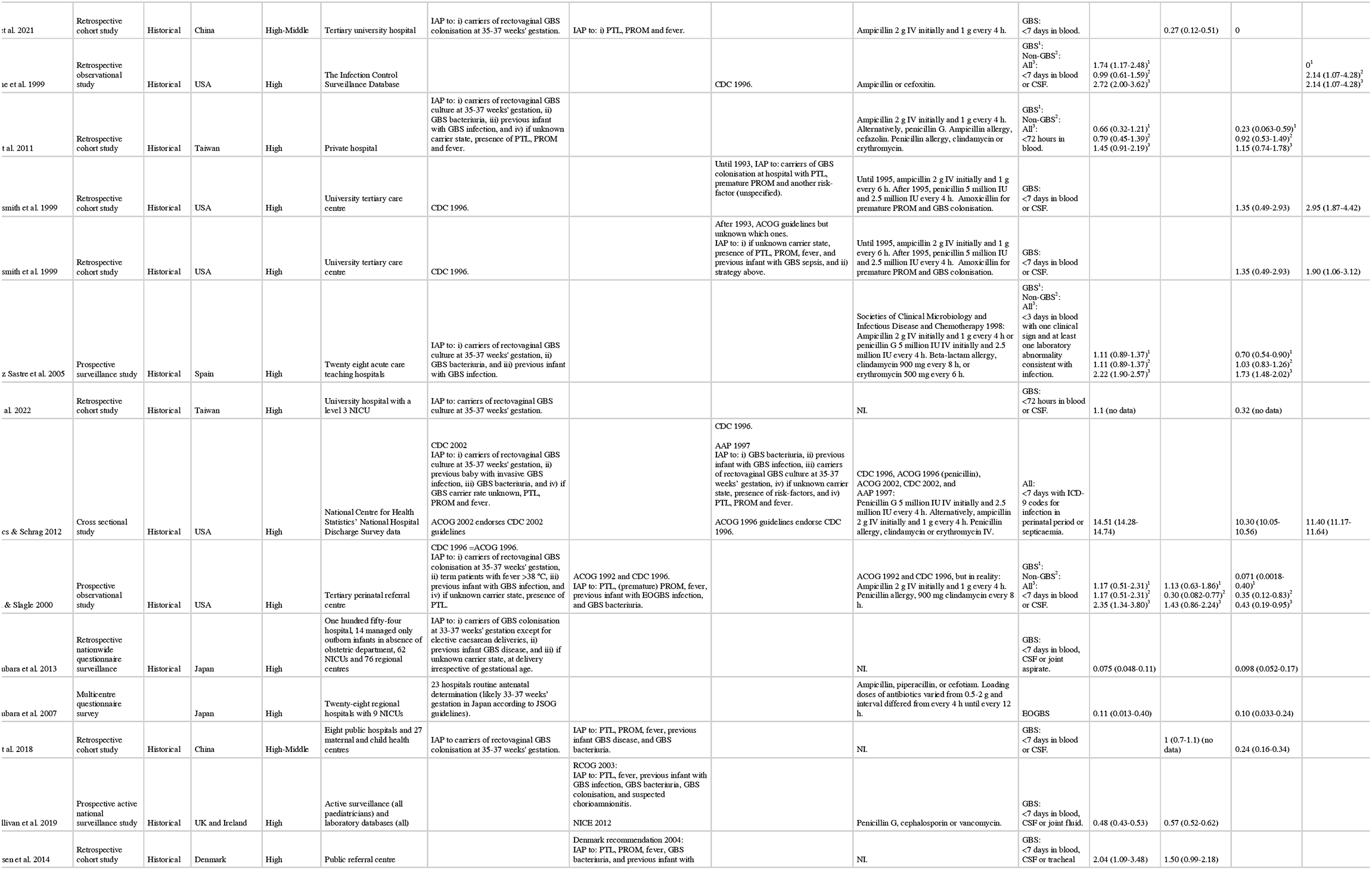
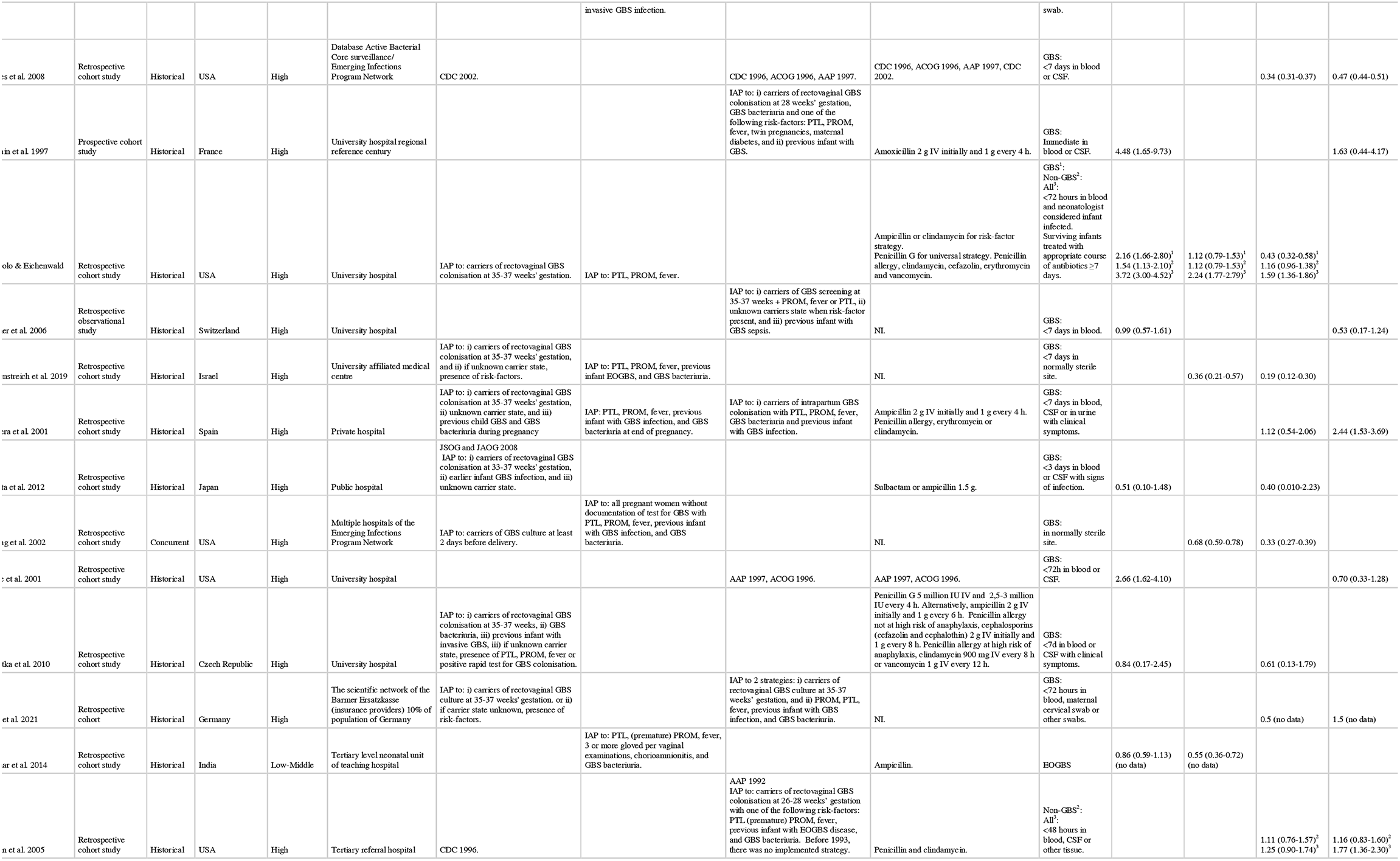
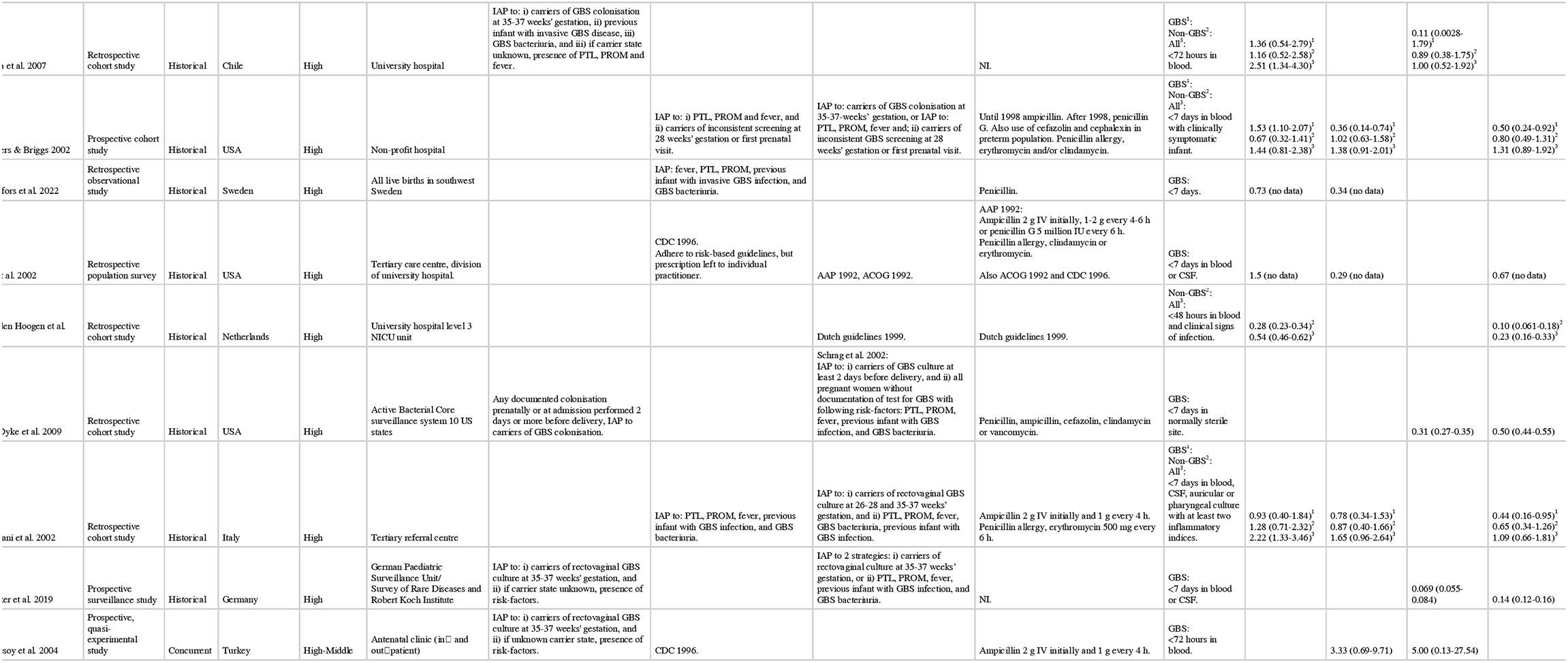
Early-onset infection.

### Risk of bias within studies

Overall risk of bias was critical in 3 studies, serious in 43 studies, moderate-serious in 3 studies, moderate in 22 studies and low in 1 study (<u>Supplementary file 3</u>). Many studies had problems in the domain of confounding due to a lack of data on the demographics for the total population or issues with retrospective outcome measurements, which were not standardised.

### Synthesis of results

#### EOGBS infection

Any strategy (i.e. risk-based, universal or ‘other’) was associated with a reduced risk of EOGBS infection compared to no strategy (n=34 studies, RR 0.46 (0.36-0.60), I^2^=93%, Chi^2^-test p<0.001) (Figure 2 and <u>Supplementary files 4, 5 and 6</u>). Similarly, risk-based (n=11 studies, RR 0.65 (0.48-0.87)) and universal strategies (n=16 studies, RR 0.37 (0.25-0.55)) were also associated with reduced risk of EOGBS infection as compared to no strategy (Figure 3, 4 and <u>Supplementary file 4, 5 and 6</u>). In direct comparison, universal strategies were significantly associated with a reduced risk of EOGBS infection compared to risk-based strategies (n=17 studies, RR 0.41 (0.30-0.55), Figure 5 and <u>Supplementary file 4, 5 and 6</u>). Results were similar for studies reporting on term incidence and after excluding studies with a serious risk of bias (sensitivity analyses) (<u>Supplementary file 3, 4, 5 and 6</u>).

**Figure 2:**
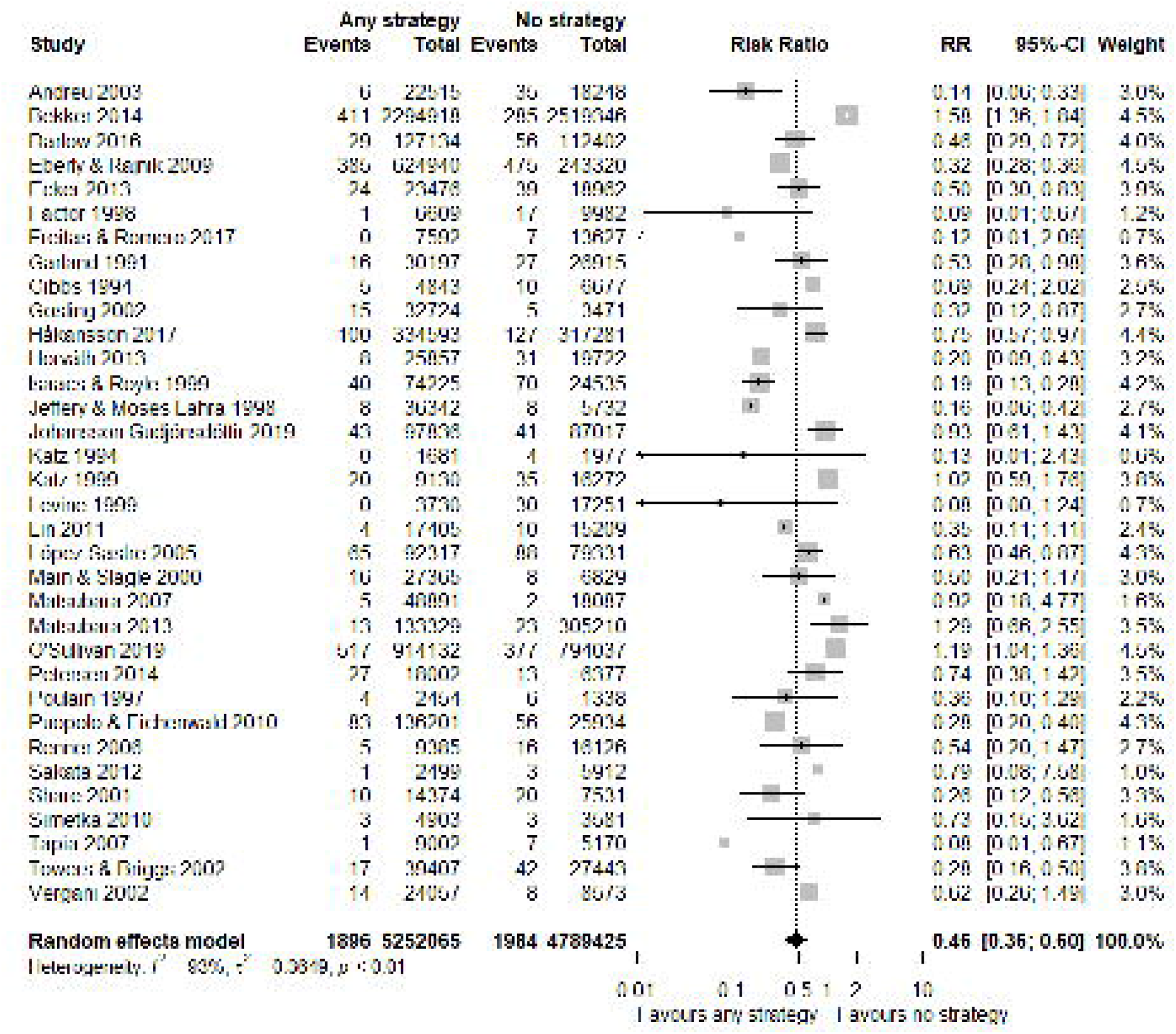
Forest plot of EOGBS infection: any vs no strategy Forest plot of risk ratio (with 95%-CI) of EOGBS infection in any strategy versus no strategy.

**Figure 3:**
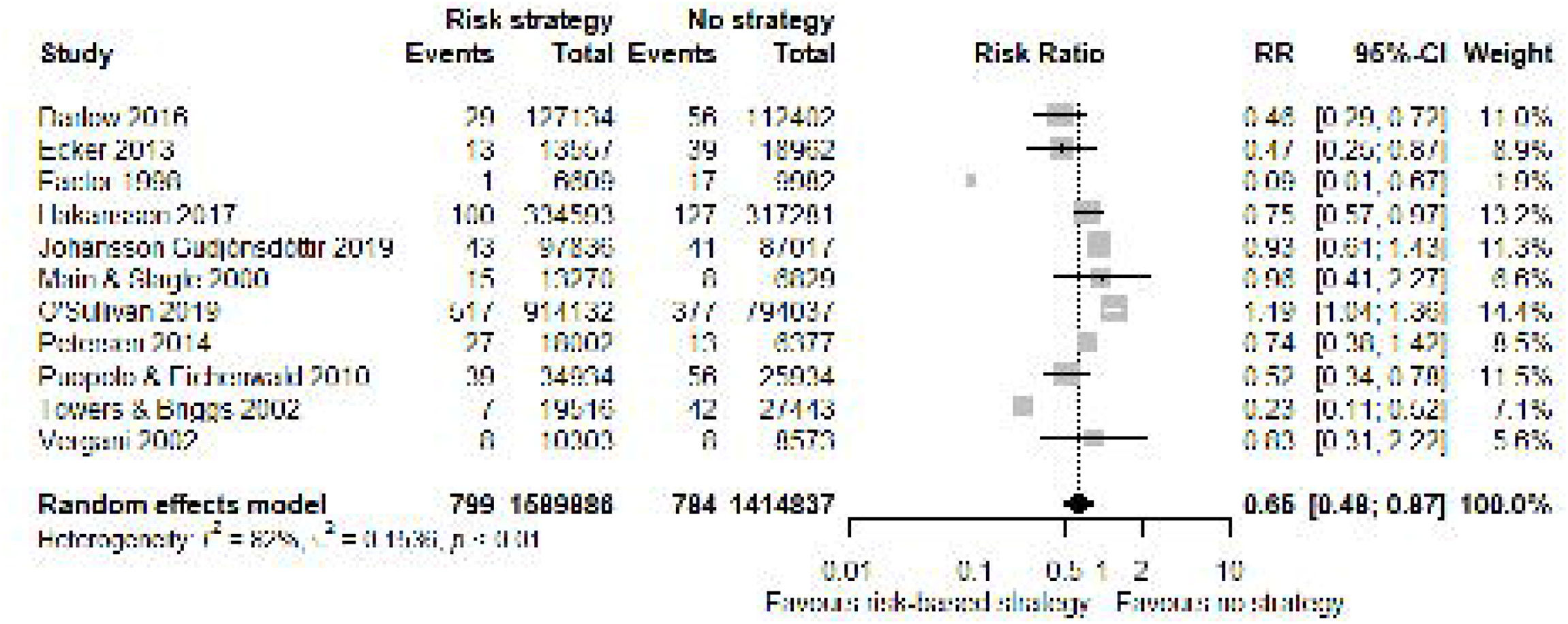
Forest plot of EOGBS infection: risk vs. no strategy Forest plot of risk ratio (with 95%-CI) of EOGBS infection in risk-based strategies versus no strategy.

**Figure 4:**
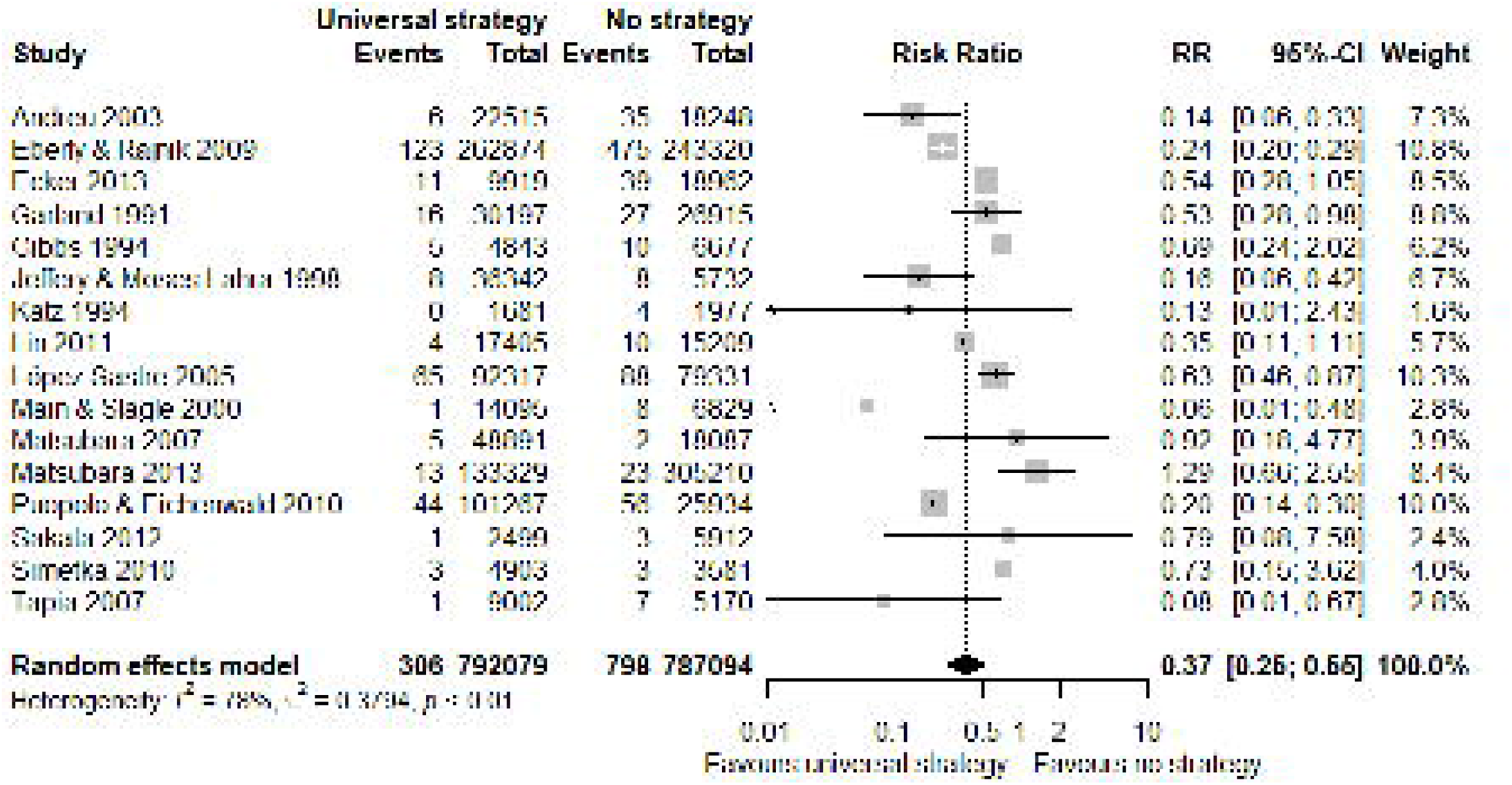
Forest plot of EOGBS infection: universal vs. no strategy Forest plot of risk ratio (with 95%-CI) of EOGBS infection in universal strategies versus no strategy.

**Figure 5:**
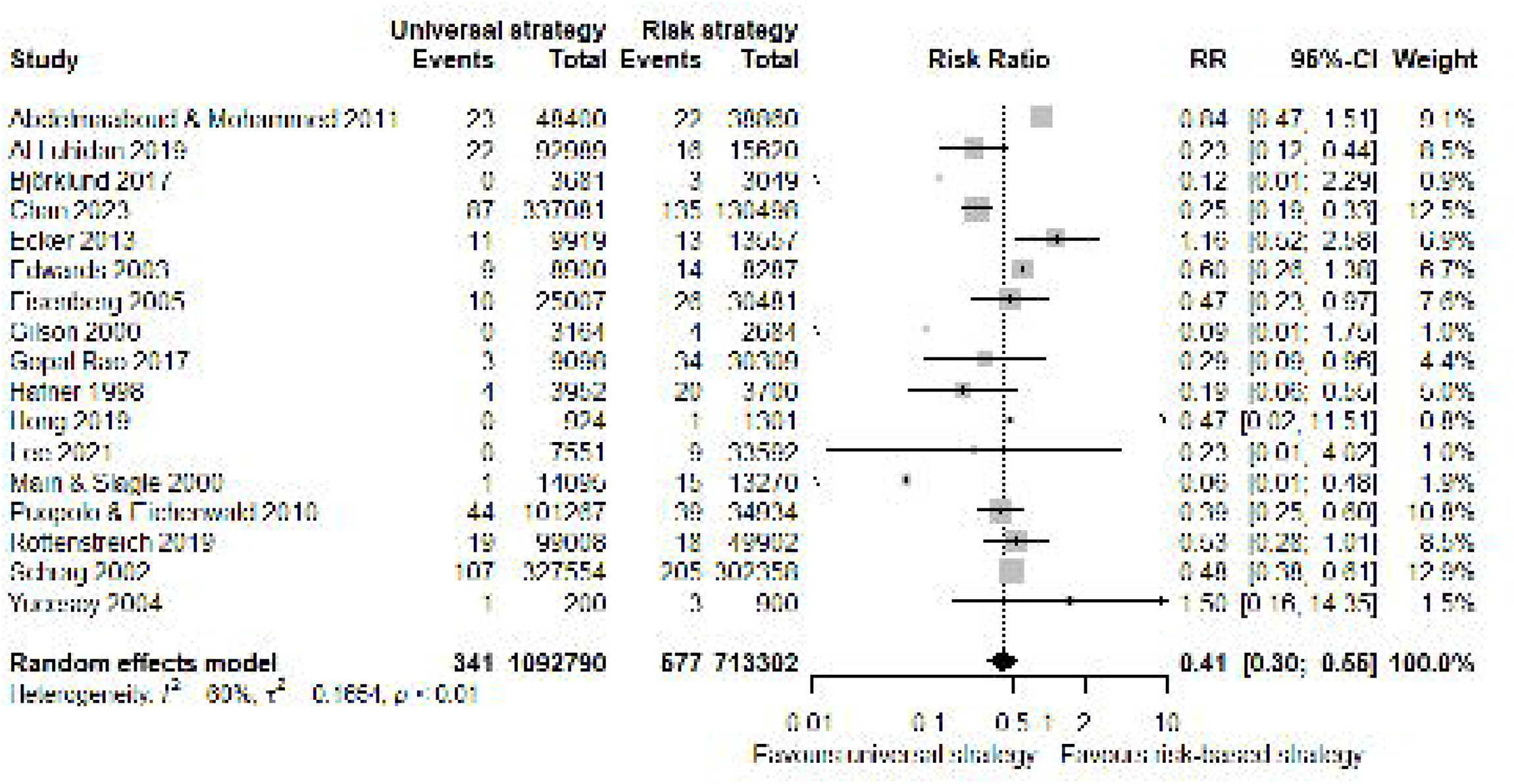
Forest plot of EOGBS infection: universal vs. risk strategies Forest plot of risk ratio (with 95%-CI) of EOGBS infection in universal strategies versus risk-based strategies.

Meta-regression demonstrated that there was a correlation between the baseline EOGBS infection incidence and the differences between any strategy and no strategy (β=-0.463, p<0.001) and between universal and no strategy (β=-0.746, p<0.001). This indicates that the effect of any or universal strategies compared to no strategy is increased if the incidence in the population is higher. This was not the case in the comparison between risk-based and no strategy (β=-0.328, p=0.082).

#### Timing of GBS determination

Vergani et al.^2^ reported no significant differences in EOGBS infection between early and late antepartum determination (RR 1.24 (0.25-6.12)), while El Helali et al. reported that intrapartum determination significantly reduced EOGBS infection compared to antepartum determination (RR 0.21 (0.07-0.64)) (<u>Supplementary file 2, 4 and 5</u>).^43,92^ Meta-regression demonstrated that there was no correlation between timing of GBS determination and the differences between universal and no or risk-based strategies.

#### Additional early-onset infection

Any strategy was associated with a reduced risk of non-GBS EOS compared to no strategy, but individual strategies did not significantly impact non-GBS EOS (<u>Supplementary file 2, 4,</u> <u>5</u>). Similar to EOGBS infection comparisons, any, risk-based and universal strategies were all significantly associated with a reduced risk of all EOS compared to no strategy (<u>Supplementary file 2, 4, 5</u>). Universal strategies were associated with a reduced risk of all EOS compared to risk-based strategies (<u>Supplementary file 2, 4, 5</u>).

#### EOGBS-related mortality

The pooled EOGBS-related mortality rate during periods with no strategy (0.089 (0.047-0.17), n=15 studies, I^2^=86%) was reduced by more than half after implementation of any strategy (0.028 (0.022-0.036, n=19 studies, I^2^=0%) (<u>Table 2</u>). EOGBS-related mortality rate was similar in periods with risk-based (0.026 (0.019-0.037), n=6 studies, I^2^=36%) and universal strategies (0.028 (0.015-0.054, n=10 studies, I^2^=0%) (<u>Table 2</u>).

#### IAP administration

Pooled IAP administration rate during periods with no strategy (8 (3-17) %, n=3 studies, I^2^=100%) more than doubled with any strategy (19 (16-22) %, n=16 studies, I^2^=100%). In the comparison of risk-based (16 (12-20) %, n=11 studies, I^2^=97%) and universal strategies (21 (18-24) %, n=12 studies, I^2^=98%) no significant difference in the rate of IAP administration was found (n=9 studies, RR 1.29 (0.95-1.75), I^2^=99%, Chi^2^-test p<0.001) (<u>Table 3</u>, and <u>Supplementary file 4, 6</u>).

#### Antimicrobial resistance

In the 11 studies reporting on antimicrobial resistance of EOGBS isolates, there was no resistance to ampicillin, penicillin or other β-lactams but varying resistance to other second-line antimicrobials, including erythromycin and clindamycin, but not vancomycin (Supplementary file 2).

## Discussion

Our study adds to the data from previous reviews by expanding the number of included studies and thus the population size, as well as by exploring outcomes not previously reported (non-GBS early-onset sepsis (EOS), mortality and maternal peripartum infection).^12,13^ In this systematic review and meta-analysis of 72 studies that included more than 10 million live births and pregnant women, all strategies (i.e. any, risk-based, universal and ‘other’ strategies) were associated with a lower risk of early-onset Group B Streptococcal (EOGBS) infection. Universal strategies were associated with a lower risk of EOGBS infection compared to risk-based strategies, while intrapartum antibiotic prophylaxis (IAP) rate was not significantly different between strategies without report of antimicrobial resistance of EOGBS isolates to penicillin or ampicillin. Pooled EOGBS-related mortality was halved in periods with any strategy compared to periods with no strategy and non-GBS EOS incidence decreased after implementation of any strategy. This systematic review and meta-analysis confirms the findings of two previous reviews that universal strategies are more effective in preventing EOGBS infection compared to having no strategy or risk-based strategies.^12,13^

In this review, evidence was mostly derived from observational studies conducted in high- and high-middle-income countries with historical controls (pre-post implementation studies). These study limitations are reflected in serious risks of bias, often due to potential confounding and potential time bias^3^. Regardless, after sensitivity analyses, results were similar with respect to EOGBS infection incidence, in which the data was graded as moderate-level certainty evidence. Other limitations included heterogeneity of the strategies used, which led to classifying all that did not fit the terms ‘risk-based’ or ‘universal’ into an ‘other’ category. This category consisted of at least 10 different strategies, which could not be pooled.

The results of this systematic review and meta-analysis need to be seen in context, by including consideration with regard to cost, feasibility and providers’ and women’s views. Universal strategies appear to be optimal in preventing EOGBS, with most protocols determining GBS colonisation between 35 and 37 weeks’ gestation.^4,7,107^ However, inherently, these protocols do not take into account preterm birth occurring before determination, in which EOGBS morbidity and mortality are higher.^4^ It is known that GBS diagnostics with regard to GBS colonisation lose accuracy as time progresses between diagnosis and birth (>6 weeks), indicative of possible transient colonisation.^108^ Earlier screening would, therefore, signify a less accurate prediction of EOGBS infection risk for infants, but our findings do not support this theory. Although intrapartum determination might lower EOGBS infection compared to antenatal determination, intrapartum testing is not yet widely available and more research to understand the potential harms and benefits is needed.

Discussion concerning the potential for IAP strategies to contribute to harm, such as increased non-GBS neonatal sepsis have featured prominently in this space, in addition to possible overtreatment and rising antimicrobial resistance with universal strategies, on account of associations between early antibiotic exposure and altered gut microbiome, asthma and obesity.^14–16,109,110^ Interestingly, instead of an increase in non-GBS EOS, we observed that implementation of any strategy was associated with a lower risk of non-GBS EOS. The studies reporting an increasing incidence of gram-negative sepsis (particularly *E. coli*) primarily focused on very low-birthweight infants, while studies in premature and term infants (not included in our meta-analysis) did not observe such an increase, similar to our findings.^16,30,109,111^ Our findings also suggest that antimicrobial resistance in EOGBS isolates is confined to second-line antimicrobials. Hence, penicillin and ampicillin might still be the best choice for IAP in preventing vertical transmission of GBS, especially since recent evidence suggests that clindamycin is less effective than ampicillin against vertical transmission of GBS.^112^ Moreover, our findings provide some reassurance that universal strategies are not likely to result in significantly higher antibiotic exposure compared to risk-based strategies.

Despite evidence for this systematic review and meta-analysis predominantly deriving from studies conducted in high and high-middle-income countries, GBS prevention strategies might also lower the high EOGBS burden in low- and lower-middle income countries. Low and lower-middle-income countries more commonly have no strategy for IAP, partly due to lack of resources and inadequate infrastructure for diagnostic screening and testing, while they have higher EOGBS morbidity and mortality.^113–115^ In our review, we included one study that reported on the adoption of a strategy in a lower-middle-income country^4^ (India).^83^ After the adoption of a risk-based strategy, EOGBS infection incidence significantly decreased, particularly in premature infants, without any antimicrobial resistance for penicillin and ampicillin in EOGBS isolates.^83^ As these results are similar to the findings in this review, IAP strategies might prove beneficial in these settings, though more research in low- and lower-middle income countries is necessary.

Worldwide annual EOGBS burden is about 205 000 affected newborn infants, of which a significant proportion results in death.^115^ Any IAP strategy aimed to lower this EOGBS burden could reduce risk of EOGBS infection and non-GBS sepsis, while also lowering EOGBS-related mortality. Specifically, universal strategies likely lead to a larger reduction in EOGBS infection compared to risk-based strategies while a similar proportion of pregnant women receive IAP. Considering that EOGBS isolates were not resistant to ampicillin or penicillin, altogether, our findings do not support evidence that IAP strategies, and in particular, universal strategies, are associated with explicit harm, though data was derived from observational studies.

Currently, the randomised multicentre GBS3 (ISRCTN49639731) trial is being conducted in the United Kingdom comparing risk-based and universal strategies (antepartum and intrapartum). It is the first trial to concurrently investigate (cost)-effectiveness of the different strategies and will provide valuable insight into the optimal GBS prevention strategy.

## Data Availability

All data were presented in the manuscript and supplementary files.

## Contributors

Conceptualisation: TJRP, GFH, CA, TL, ABtP, VB and TvdA. Data curation: TJRP. Formal analysis: TJRP. Investigation: TJRP, GFH, YC, CA, TL and VB. Methodology: TJRP, GFH, CA, TL, ABtP, VB and TvdA. Project administration: TJRP, TL, TvdA. Resources and software: Not relevant for this review. Supervision: TL and TvdA. Validation: GFH. Visualisation: TJRP. Writing – original draft: TJRP and GFH. Writing – review & editing: YC, CA, TL, ABtP, VB and TvdA. All authors reviewed and approved the final version before submission. All authors had access to all data used in this study, approved the final version of the manuscript, and accepted the responsibility for the decision to submit the manuscript for publication.

## Data sharing

All data were presented in the manuscript and supplementary files.

## Acknowledgements

This work was funded by the UNDP-UNFPA-UNICEF-WHO-World Bank Special Programme of Research, Development and Research Training in Human Reproduction (HRP), Department of Sexual and Reproductive Health and Research (SRH), a co-sponsored programme executed by the World Health Organization. We want to acknowledge the support of MA Claudia Pees, whom assisted in search strategy formation.

## Footnotes

1 If data was already published in other articles, then the study with the largest sample size was chosen and included.

2 Vergani et al. used a combination strategy consisting of IAP administration to pregnant women with GBS colonisation and to those with risk-factors.

3 The time bias could consist of possible improvements in care over time, but is contrasted by increased viability of premature infants and subsequent survival that could increase the amount of EOGBS cases.

4 This study was not included in the synthesis of EOGBS infection, because no raw data was available. The study characteristics and results are mentioned in the corresponding tables.

