## Supplementary file 1 for "Intrapartum antibiotic prophylaxis to prevent Group B streptococcal infections in newborn infants: a systematic review and meta-analysis comparing various strategies"

### Search Strategy MEDLINE

("streptococcus agalactiae"[MeSH] OR "streptococcus agalactiae"[tw] OR "group b streptococcus"[tw] OR "streptococci agalactiae"[tw] OR "group b streptococci"[tw] OR "group b streptococcal"[tw])

AND

("pregnancy"[MeSH] OR "pregnancy"[tw] OR "pregnancies"[tw] OR "pregnant"[Tw] OR "maternal"[Tw] OR "intrapartum"[Tw] OR "intra partum"[Tw] OR "antepartum"[Tw] OR "ante partum"[Tw] OR "peripartum"[Tw] OR "peri partum"[Tw]OR "Infant, Newborn"[Mesh] OR "Newborn"[Tw] OR "Newborns"[Tw] OR "Neonate"[Tw] OR "Neonates"[Tw] OR "neonatal"[tw] OR "Neonatal Sepsis"[mesh])

AND

("screening"[tw] OR "screenings"[tw] OR "screen"[tw] OR "screens"[tw] OR "screened"[tw] OR "culture based"[tw] OR "culturebased"[tw] OR "risk based"[tw] OR "risk factor"[tw] OR "riskbased"[tw] OR "Guideline" [Publication Type] OR "guidelines"[Tw] OR "guideline"[Tw] OR "prevention and control" [Subheading] OR "prevention"[Tw] OR "preventions"[Tw] OR "prevent"[Tw] OR "prevents"[Tw] OR "preventing"[tw] OR "early onset"[Tw] OR "Polymerase Chain Reaction"[Mesh] OR "Polymerase Chain Reaction"[Tw] OR "Polymerase Chain Reactions"[Tw] OR "PCR"[Tw])

3661 results (13-05-2024)
