## Supplementary file 2 for "Intrapartum antibiotic prophylaxis to prevent Group B streptococcal infections in newborn infants: a systematic review and meta-analysis comparing various strategies"

**Table 2: Timing of GBS determination**

| **Author Year** | **Study design** | **Time analysis** | **Country** | **Country Income** | **Study setting/ Data source** | **IAP criteria: Universal strategy 1** | **IAP criteria: Universal strategy 2** | **IAP agent:** | **Infection definition** | **EOGBS-related mortality incidence per 1000 live births or pregnant women (95%-CI)** | | | |
| --- | --- | --- | --- | --- | --- | --- | --- | --- | --- | --- | --- | --- | --- |
|  |  |  |  |  |  |  |  |  |  | **Early antenatal** | **Late antenatal** | **Antenatal** | **Intrapartum** |
| El Helali et al. 2019 | Retrospective cohort study | Historical | France | High | General hospital | IAP to: i) carriers of rectovaginal GBS culture at 35-37 weeks’ gestation, ii) GBS bacteriuria, iii) previous infant with EOGBS, and iv) if unknown carrier state, PTL, PROM and fever. | IAP to: i) carriers of intrapartum rectovaginal GBS polymerase chain reaction, ii) GBS bacteriuria, iii) previous infant with EOGBS, and iv) if unknown carrier state, PTL, PROM and fever. | Penicillin G 5 million IU IV initially and 2.5 million IU every 4 h. If penicillin allergy, clindamycin or vancomycin, depending susceptibility. | GBS: <7 days in blood or CSF with clinical signs or biological abnormalities consistent with sepsis |  |  | 1.02 (0.52-1.78) | 0.21 (0.057-0.54) |
| Vergani et al. 2002 | Retrospective cohort study | Historical | Italy | High | Tertiary referral centre | IAP to: i) carriers rectovaginal GBS culture at 26-28 weeks’ gestation, and ii) PTL, PROM, fever, GBS bacteriuria, previous infant with GBS infection. | IAP to: i) carriers of rectovaginal GBS culture 35-37 weeks’ gestation, and ii) PTL, PROM, fever, GBS bacteriuria, previous infant with GBS infection. | Ampicillin 2 g IV initially and 1 g every 4 h. Penicillin allergy, erythromycin 500 mg every 6 h. | GBS:  <7 days in blood, CSF, auricular or pharyngeal culture with at least two inflammatory indices. | 0.39 (0.081-0.12) | 0.49 (0.10-1.42) |  |  |

**Table 3: EOGBS-related mortality**

| **Author Year** | **Study design** | **Time analysis** | **Country** | **Country Income** | **Study setting/ Data source** | **IAP criteria: Universal strategies** | **IAP criteria:  Risk-based strategies** | **IAP criteria: Other strategies** | **IAP agent:** | **Infection definition** | **EOGBS-related mortality incidence per 1000 live births or pregnant women (95%-CI)** | | | |
| --- | --- | --- | --- | --- | --- | --- | --- | --- | --- | --- | --- | --- | --- | --- |
|  |  |  |  |  |  |  |  |  |  |  | **No strategy** | **Risk strategy** | **Universal strategy** | **Other strategies** |
| Freitas & Romero 2017 | Retrospective cohort study | Historical | Brazil | High-Middle | Regional maternity hospital |  |  | Based on CDC 2010. | Ampicillin 2g IV initially and 1 g every 4 h. Penicillin allergy, clindamycin 900 mg IV every 8 hours | GBS: <72 hours in blood and antibiotic treatment ≥5 days or death <5 days while on antibiotic treatment. | 0.22 (0.045-0.64) |  |  | 0 |
| Garland 1991 | Retrospective observational study | Concurrent | Australia | High | Public teaching hospital | IAP to: carriers vaginal GBS culture at 32 weeks' gestation. |  |  | Penicillin 1 million IU IV every 6 h. Penicillin allergy, erythromycin 500 mg every 6 h. | EOGBS: in blood, CSF, surface swabs or urine | 0.30 (0.13-0.59) |  | 0.033 (0.00084-0.18) |  |
| Gopal Rao et al. 2017 | Retrospective observational study | Historical | UK | High | General hospital | IAP to: carriers of rectovaginal GBS colonisation at 35-37 weeks' gestation | NICE 2012. |  | Penicillin G 3 g IV initially and 1.5 g every 4 h. Penicillin allergy, clindamycin. | GBS: <7 days in blood, CSF or other sterile fluids. |  | 0.033 (0.00084-0.18) | 0 |  |
| Hafner et al. 1998 | Retrospective and prospective cohort study | Historical | Austria | High | Sociomedical centre | IAP to: carriers of rectovaginal GBS culture at 33-35 weeks' gestation. | IAP to: (premature) PTL, PROM, fever, previous infant with GBS infection, and maternal diabetes mellitus. |  | Amoxicillin with clavulanic acid 2.2 g every 6 h. Penicillin allergy, 600 mg clindamycin. | GBS: in throat, umbilicus, ears or blood and signs of sepsis. |  | 0 | 0 |  |
| Håkansson et al. 2017 | Retrospective cohort study | Historical | Sweden | High | National health registers of all hospitals in Sweden |  | IAP to: PTL, PROM, fever, GBS bacteriuria, and previous infant with GBS infection. |  | NI. | GBS: <7 days in blood or CSF. | 0.015 (0.0069-0.041) | 0.015 (0.0049-0.035) |  |  |
| Isaacs & Royle 1999 | Prospective observational study | Historical | Australia | High | Australian neonatal units of the Australasian Study Group for Neonatal Infection |  |  | Different strategies in hospitals comprising no, risk-based, universal and other strategies. | Usually ampicillin or penicillin. | GBS: <48 hours in blood, CSF or urine with clinical sepsis. | 0.20 (0.066-0.48) |  |  | 0.054 (0.015--0.14) |
| Jeffery & Moses Lahra 1998 | Prospective cohort study | Historical | Australia | High | Tertiary referral hospital | IAP to: i) carriers of vaginal GBS colonisation at 28 weeks' gestation without risk-factors or 24 weeks if known risk-factor for preterm birth, ii) GBS bacteriuria, iii) previous infant with EOGBS infection, and iv) if unknown carrier state, presence of preterm labour. |  |  | Ampicillin 1 g every 6 hours. Penicillin allergy, cephalosporin. | GBS: <48 hours in blood and clinical signs of sepsis. | 0.52 (0.11-1.53) |  | 0 |  |
| Katz et al. 1994 | Retrospective cohort study | Historical | USA | High | Public academic tertiary care medical centre | IAP to: i) carriers of rectovaginal GBS culture at 24-28 weeks' gestation, ii) GBS bacteriuria, and iii) if carrier state unknown, presence of PROM and PTL. |  |  | Ampicillin 2 g IV every 6 hours. Penicillin allergy, cefazolin 500 mg or clindamycin 600 mg every 6 h. | GBS: in blood | 0.51 (0.013-2.81) |  | 0 |  |
| Katz et al. 1999 | Retrospective cohort study | Historical | USA | High | Urban tertiary centre |  |  | IAP to: i) carriers of rectovaginal GBS culture at 28 weeks' gestation and preterm delivery; and ii) (premature) PROM, fever and previous infant with GBS infection. | Ampicillin 2 g IV every 6 h. Penicillin allergy, clindamycin 600 mg every 6 h. | GBS: <7 days in blood or CSF with clinical signs of sepsis. | 0.31 (0.10-0.72) |  |  | 0 |
| López Sastre et al. 2005 | Prospective surveillance study | Historical | Spain | High | Twenty eight acute care teaching hospitals | IAP to: i) carriers of rectovaginal GBS culture at 35-37 weeks' gestation, ii) GBS bacteriuria, and iii) previous infant with GBS infection. |  |  | Societies of Clinical Microbiology and Infectious Disease and Chemotherapy 1998: Ampicillin 2 g IV initially and 1 g every 4 h or penicillin G 5 million IU IV initially and 2.5 million IU every 4 h. Beta-lactam allergy, clindamycin 900 mg every 8 h, or erythromycin 500 mg every 6 h. | GBS: <3 days in blood with one clinical sign and at least one laboratory abnormality consistent with infection. | 0.050 (0.014-0.13) |  | 0.043 (0.012-0.11) |  |
| Matsubara et al. 2013 | Retrospective nationwide questionnaire surveillance | Historical | Japan | High | One hundred fifty-four hospital, 14 managed only outborn infants in absence of obstetric department, 62 NICUs and 76 regional centres | IAP to: i) carriers of GBS colonisation at 33-37 weeks' gestation except for elective caesarean deliveries, ii) previous infant GBS disease, and iii) if unknown carrier state, at delivery irrespective of gestational age. |  |  | NI. | GBS: <7 days in blood, CSF or joint aspirate. | 0.026 (0.011-0.052) |  | 0.030 (0.0082-0.077) |  |
| O'Sullivan et al. 2019 | Prospective active national surveillance study | Historical | UK and Ireland | High | Active surveillance (all paediatricians) and laboratory databases (all) |  | RCOG 2003, NICE 2012. |  | Penicillin G, cephalosporin or vancomycin. | GBS: <7 days in blood, CSF or joint fluid. | 0.048 (0.034-0.066) | 0.030 (0.019-0.043) |  |  |
| Renner et al. 2006 | Retrospective observational study | Historical | Switzerland | High | University hospital |  |  | IAP to: i) carriers of GBS screening at 35-37 weeks + PROM, fever or PTL, ii) unknown carriers state when risk-factor present, and iii) previous infant with GBS sepsis. | NI. | GBS: <7 days in blood. | 0.062 (0.0016-0.35) |  |  | 0 |
| Sagrera et al. 2001 | Retrospective cohort study | Historical | Spain | High | Private hospital | IAP to: i) carriers of rectovaginal GBS colonisation at 35-37 weeks' gestation, ii) unknown carrier state, and iii) previous child GBS and GBS bacteriuria during pregnancy | IAP: PTL, PROM, fever, previous infant with GBS infection, and GBS bacteriuria at end of pregnancy. | IAP to: i) carriers of intrapartum GBS colonisation with PTL, PROM, fever, GBS bacteriuria and previous infant with GBS infection. | Ampicillin 2 g IV initially and 1 g every 4 h. Penicillin allergy, erythromycin or clindamycin. | GBS: <7 days in blood, CSF or in urine with clinical symptoms. |  |  | 0 | 0.33 (0.069-0.97) |
| Share et al. 2001 | Retrospective cohort study | Historical | USA | High | University hospital |  |  | AAP 1997, ACOG 1996. | AAP 1997, ACOG 1996. | GBS: <72h in blood or CSF. | 0 |  |  |  |
| Simetka et al. 2010 | Retrospective cohort study | Historical | Czech Republic | High | University hospital | IAP to: i) carriers of rectovaginal GBS colonisation at 35-37 weeks, ii) GBS bacteriuria, iii) previous infant with invasive GBS, iii) if unknown carrier state, presence of PTL, PROM, fever or positive rapid test for GBS colonisation. |  |  | Penicillin G 5 million IU IV and 2,5-3 million IU every 4 h. Alternatively, ampicillin 2 g IV initially and 1 g every 6 h. Penicillin allergy not at high risk of anaphylaxis, cephalosporins (cefazolin and cefalotin) 2 g IV initially and 1 g every 8 h. Penicillin allergy at high risk of anaphylaxis, clindamycin 900 mg IV every 8 h or vancomycin 1 g IV every 12 h. | GBS: <7d in blood or CSF with clinical symptoms. | 0 |  | 0 |  |
| Trijbels-Smeulders et al. 2007 | Retrospective observational study | Historical | The Netherlands | High | Dutch Paediatric Surveillance Unit database |  |  | Dutch guidelines 1999. | Dutch guidelines 1999. | GBS: <7 d in blood, CSF or other sterile fluids | 0.028 (0.014-0.050) |  |  | 0.028 (0.016-0.045) |
| Vergani et al. 2002 | Retrospective cohort study | Historical | Italy | High | Tertiary referral centre |  | IAP to: PTL, PROM, fever, previous infant with GBS infection, and GBS bacteriuria. | IAP to: i) carriers of rectovaginal GBS culture at 26-28 and 35-37 weeks’ gestation, and ii) PTL, PROM, fever, GBS bacteriuria, previous infant with GBS infection. | Ampicillin 2 g IV initially and 1 g every 4 h. Penicillin allergy, erythromycin 500 mg every 6 h. | GBS:  <7 days in blood, CSF, auricular or pharyngeal culture with at least two inflammatory indices. | 0.47 (0.13-1.19) | 0.097 (0.0025-0.54) |  | 0 |
| Yücesoy et al. 2004 | Prospective, quasi-experimental study | Concurrent | Turkey | High-Middle | Antenatal clinic (in‐ and out‐patient) | IAP to: i) carriers of rectovaginal GBS culture at 35-37 weeks' gestation, and ii) if unknown carrier state, presence of risk-factors. | CDC 1996. |  | Ampicillin 2 g IV initially and 1 g every 4 h. | GBS: <72 hours in blood. |  | 0 | 0 |  |

**Table 4: IAP administration**

| **Author Year** | **Study design** | **Time analysis** | **Country** | **Country Income** | **Study setting/ Data source** | **IAP criteria: Universal strategies** | **IAP criteria:  Risk-based strategies** | **IAP criteria: Other strategies** | **IAP agent** | **Percentage of pregnant women receiving IAP** | | | |
| --- | --- | --- | --- | --- | --- | --- | --- | --- | --- | --- | --- | --- | --- |
|  |  |  |  |  |  |  |  |  |  | **No strategy** | **Risk strategy** | **Universal strategy** | **Other strategies** |
| Björklund et al. 2017 | Retrospective cohort study | Historical | Finland | High | Public financed tertiary delivery unity | IAP to: i) carriers of rectovaginal GBS polymerase chain reaction at admission, and ii) if carrier state unknown on presence of risk-factors. | IAP to: PROM, GBS bacteriuria, GBS colonisation, and previous infant with EOGBS. |  | Penicillin G 5 million IU IV initially and 2.5 million IU every 4 h. Penicillin allergy, cefuroxime 1.5 g IV initially and 750 mg every 8 h or clindamycin 900 mg IV every 8 h. |  | 27.1 (25.5-28.7) | 27.9 (26.5-29.4) |  |
| Coco 2002 | Retrospective cohort study | Historical | USA | High | Family practice residency maternal service | IAP to: carriers of rectovaginal GBS culture after 35 weeks’ gestation. | IAP to: PROM and fever. |  | NI. |  | 18.5 (13.5-24.9) | 20.5 (15.1-27.2) |  |
| Davis et al. 2001 | Retrospective cohort study | Historical | USA | High | Nonprofit group and network health maintenance organisation with data from 2 hospitals | CDC 1996. |  | AAP 1992 | Penicillin G |  |  | 26.7 (25.0-28.5) | 12.8 (11.1-14.7) |
| Hafner et al. 1998 | Retrospective and prospective cohort study | Historical | Austria | High | Sociomedical centre | IAP to: carriers of rectovaginal GBS culture at 33-35 weeks' gestation. | IAP to: (premature) PTL, PROM, fever, previous infant with GBS infection, and maternal diabetes mellitus. |  | Amoxicillin with clavulanic acid 2.2 g every 6 h. Penicillin allergy, 600 mg clindamycin. |  | 11.9 (10.9-13.0) | 13.5 (12.5-14.6) |  |
| Hong et al. 2019 | Retrospective cohort study | Historical | South-Korea | High | Tertiary hospital | CDC 2010. | IAP to: PROM, fever, GBS bacteriuria, and previous infant with GBS sepsis. |  | Cefazolin 2 g IV initially and 1 g every 8 h. Cephalosporin allergy, ampicillin 2 g IV initially and 1 g every 4 h. Penicillin allergy, vancomycin and clindamycin. |  | 10.9 (9.3-12.7) | 21.5 (18.9-24.3) |  |
| Katz et al. 1999 | Retrospective cohort study | Historical | USA | High | Urban tertiary centre |  |  | IAP to: i) carriers of rectovaginal GBS culture at 28 weeks' gestation and PTL, and ii) (premature) PROM, fever and previous infant with GBS infection. | Ampicillin 2 g IV every 6 h. Penicillin allergy, clindamycin 600 mg every 6 h. | 15.7 (15.1-16.3) |  |  |  |
| Locksmith et al. 1999 | Retrospective cohort study | Historical | USA | High | University tertiary care centre | CDC 1996. |  | Until 1993, IAP to: carriers of GBS colonisation at hospital with PTL, premature PROM and another risk-factor (unspecified). | Until 1995, ampicillin 2 g IV initially and 1 g every 6 h. After 1995, penicillin 5 million IU and 2.5 million IU every 4 h. Amoxicillin for premature PROM and GBS colonisation. |  |  | 21.0 (19.8-22.2) | 9.4 (8.8-10.1) |
| Locksmith et al. 1999 | Retrospective cohort study | Historical | USA | High | University tertiary care centre | CDC 1996. |  | After 1993, ACOG guidelines but unknown which ones. IAP to: i) if unknown carrier state, presence of PTL, PROM, fever, and previous infant with GBS sepsis, and ii) strategy above | Until 1995, ampicillin 2 g IV initially and 1 g every 6 h. After 1995, penicillin 5 million IU and 2.5 million IU every 4 h. Amoxicillin for premature PROM and GBS colonisation. |  |  | 21.0 (19.8-22.2) | 12.9 (12.2-13.7) |
| Levine et al. 1999 | Retrospective observational study | Historical | USA | High | The Infection Control Surveillance Database |  |  | CDC 1996 | Ampicillin or cefoxitin. | 3.0 (27.6-32.7) |  |  | 16.0 (14.9-17.2) |
| Schrag et al. 2002 | Retrospective cohort study | Concurrent | USA | High | Multiple hospitals of the Emerging Infections Program Network | IAP to: carriers of GBS culture at least 2 days before delivery. | IAP to: all pregnant women without documentation of test for GBS with PTL, PROM, fever, previous infant with GBS infection, and GBS bacteriuria. |  | NI. |  | 14.6 (13.3-16.1) | 21.4 (19.8-23.0) |  |
| Schuchat et al. 2002 | Retrospective cohort study | Historical | USA | High | Multiple hospitals of the Emerging Infections Program Network | IAP to: carriers of GBS culture at least 2 days before delivery. | IAP to: all pregnant women without documentation of test for GBS with PTL, PROM, fever, previous infant with GBS infection, and GBS bacteriuria. |  | Penicillin, ampicillin, or cefazolin. |  | 9.8 (6.9-13.8) | 13.3 (10.0-17.3) |  |
| Uy et al. 2002 | Retrospective population survey | Historical | USA | High | Tertiary care centre, division of university hospital. |  | CDC 1996. Adhere to risk-based guidelines, but prescription left to individual practitioner. | AAP 1992, ACOG 1992 | AAP 1992: Ampicillin 2 g IV initially, 1-2 g every 4-6 h or penicillin G 5 million IU every 6 h. Penicillin allergy, clindamycin or erythromycin.   Also ACOG 1992 and CDC 1996. | 10.0 (7.2-13.8) | 20.7 (14.9-27.9) |  | 7.3 (4.1-12.7) |
| van Dyke et al. 2009 | Retrospective cohort study | Historical | USA | High | Active Bacterial Core surveillance system 10 US states | Any documented colonisation prenatally or at admission performed 2 days or more before delivery, IAP to carriers of GBS colonisation. |  | Schrag et al. 2002:  IAP to: i) carriers of GBS culture at least 2 days before delivery, and ii) all pregnant women without documentation of test for GBS with following risk factors: PROM, PTL, fever, GBS bacteriuria and previous infant with GBS infection. | Penicillin, ampicillin, cefazolin, clindamycin or vancomycin. |  |  | 31.7 (30.7-32.8) | 26.8 (25.4-28.2) |
| Vergani et al. 2002 | Retrospective cohort study | Historical | Italy | High | Tertiary referral centre |  | IAP to: PTL, PROM, fever, previous infant with GBS infection, and GBS bacteriuria. | IAP to: i) carriers of rectovaginal GBS culture at 26-28 and 35-37 weeks’ gestation, and ii) PTL, PROM, fever, GBS bacteriuria, previous infant with GBS infection. | Ampicillin 2 g IV initially and 1 g every 4 h. Penicillin allergy, erythromycin 500 mg every 6 h. |  | 16.8 (16.1-17.6) |  | 35.8 (35.0-36.6) |
| Wang et al. 2023 | Retrospective cohort study | Historical | China | High-Middle | Eight public hospitals and 31 health centres | IAP to: i) carriers of rectovaginal GBS colonisation at 35-37 weeks' gestation, and ii) carrier state known before 35 weeks' gestation, GBS bacteriuria or history previous infant affected by GBS disease. | IAP to: previous infant with invasive GBS disease, PTL, GBS colonisation, and GBS bacteriuria. |  | Penicillin G 5 million IU IV initially and 2.5 million IU every 4 hours. Penicillin allergy, erythromycin, clindamycin, or vancomycin according to sensitivity. |  | 8.1 (8.0-8.2) | 24.0 (23.8-24.1) |  |
| Youden et al. 2005 | Prospective observational study | Concurrent | Canada | High | Level one paediatric trauma centre | IAP to: carriers of rectovaginal GBS culture at 35-37 weeks’ gestation. | IAP to: (premature) PROM, fever, multiple gestation, previous infant with GBS infection and GBS bacteriuria. |  | Penicillin G 5 million IU initially and 2.5 million IU every 4 h, or ampicillin 2 g initially and 1 g every 4 h. Penicillin allergy, clindamycin 900 mg every 8 hours, erythromycin 500 mg every 6 h, or cefazolin 2 g every 8 h. |  | 23.8 (18.5-30.1) | 14.9 (8.2-25.6) |  |
| Yücesoy et al. 2004 | Prospective, quasi-experimental study | Concurrent | Turkey | High-Middle | Antenatal clinic (in‐ and out‐patient) | IAP to: i) carriers of rectovaginal culture at 35-37 weeks' gestation, or ii) if carrier state unknown with risk factors. | CDC 1996. |  | Ampicillin 2 g IV initially and 1 g every 4 h. |  | 21.2 (18.7-24.0) | 16.0 (11.5-21.8) |  |

**Table 5: Antimicrobial resistance**

| **Author Year** | **Study design** | **Time analysis** | **Country** | **Country Income** | **Study setting/ Data source** | **IAP criteria: Universal strategies** | **IAP criteria:  Risk-based strategies** | **IAP criteria: Other strategies** | **IAP agent** | **EOS definition** | **Antimicrobial** | **Percentage of antimicrobial resistance** | | | |
| --- | --- | --- | --- | --- | --- | --- | --- | --- | --- | --- | --- | --- | --- | --- | --- |
|  |  |  |  |  |  |  |  |  |  |  |  | **No  strategy** | **Risk strategy** | **Universal strategy** | **Other strategies** |
| Alarcon et al. 2004 | Retrospective cohort study | Historical | Spain | High | University Tertiary Public Hospital | CDC 1996. | CDC 1996. |  | CDC 1996. | E. Coli: <7 days in blood or CSF and clinical signs of infection | Ampicillin | 9/16 (56%) |  |  | 17/24 (71%) |
| Chen et al. 2001* | Retrospective cohort study | Historical | USA | High | Tertiary care referral centre |  | IAP to: PTL, PROM and fever. |  | Ampicillin. Penicillin allergy, clindamycin. | S. Aureus: <7 days in blood with 7 days of therapy or at least 2 days if infant died or transferred to another hospital before 7 days. | Ampicillin | 2/2 (100%) | 6/7 (86%) |  |  |
| Chen et al. 2001* | Retrospective cohort study | Historical | USA | High | Tertiary care referral centre |  | IAP to: PTL, PROM and fever. |  | Ampicillin. Penicillin allergy, clindamycin. | Coagulase-negative staphylococci | Ampicillin | 15/15 (100%) | 9/10 (90%) |  |  |
| Chen et al. 2001* | Retrospective cohort study | Historical | USA | High | Tertiary care referral centre |  | IAP to: PTL, PROM and fever. |  | Ampicillin. Penicillin allergy, clindamycin. | E. Coli | Ampicillin | 10/14 (71%) | 6/17 (35%) |  |  |
| Chen et al. 2005* | Retrospective cohort study | Historical | USA | High | Tertiary care referral centre | IAP to: carriers of rectovaginal GBS screening at 35-37 weeks’ gestation | IAP to: PTL, PROM and fever. |  | Ampicillin in risk-factor strategy Penicillin allergy, clindamycin. Penicillin G in universal strategy. Penicillin allergy, erythromycin or clindamycin. | GBS: <7 days in blood. | Ampicillin^1^ Penicillin^2^  Cefazolin^3^  Vancomycin^4^  Erythromycin^5^  Clindamycin^6^ | 0/57 (0%)^1^  0/57 (0%)^2^  0/57 (0%)^3^  0/57 (0%)^4^  4/57 (7%)^5^  0/57 (0%)^5^ | 0/38 (0%)^1^  0/38 (0%)^2^  0/38 (0%)^3^  0/38 (0%)^4^  1/38 (3%)^5^ 0/38 (0%)^6^ | 0/21 (0%)^1^  0/21 (0%)^2^  0/21 (0%)^3^  0/21 (0%)^4^  5/21 (24%)^5^  3/21 (14%)^6^ |  |
| Ecker et al. 2013 | Retrospective cohort study | Historical | USA | High | Regional tertiary care centre | CDC 2002. | NI. |  | Penicillin or ampicillin. | All: ≤7 days in blood, urine or CSF with appropriate treatment. | Ampicillin^1^  Penicillin^2^ | 16/86 (19%)^1^  1/57 (2%)^2^ | 30/67 (45%)^1^  2/25 (8%)^2^ | 13/36 (33%)^1^  0/17 (0%)^2^ |  |
| Ecker et al. 2013 | Retrospective cohort study | Historical | USA | High | Regional tertiary care centre | CDC 2002. | NI. |  | Penicillin or ampicillin. | GBS | Ampicillin^1^  Penicillin^2^ | 0/39 (0%)^1^  0/39 (0%)^2^ | 0/13 (0%)^1^  0/13 (0%)^2^ | 0/11 (0%)^1^  0/11 (0%)^2^ |  |
| Ecker et al. 2013 | Retrospective cohort study | Historical | USA | High | Regional tertiary care centre | CDC 2002. | NI. |  | Penicillin or ampicillin. | Enterococcus | Ampicillin^1^  Penicillin^2^ | 0/4 (0%)^1^  0/4 (0%)^2^ |  | 0/4 (0%)^1^  0/4 (0%)^2^ |  |
| Ecker et al. 2013 | Retrospective cohort study | Historical | USA | High | Regional tertiary care centre | CDC 2002. | NI. |  | Penicillin or ampicillin. | E. Coli | Ampicillin | 9/18 (50%) | 13/20 (65%) | 10/14 (71%) |  |
| Ecker et al. 2013 | Retrospective cohort study | Historical | USA | High | Regional tertiary care centre | CDC 2002. | NI. |  | Penicillin or ampicillin. | Other gram positives | Ampicillin^1^  Penicillin^2^ | 1/14 (7%)^1^  1/14 (7%)^2^ | 1/12 (8%)^1^  2/12 (17%)^2^ | 0/2 (0%)^1^  0/2 (0%)^2^ |  |
| Ecker et al. 2013 | Retrospective cohort study | Historical | USA | High | Regional tertiary care centre | CDC 2002. | NI. |  | Penicillin or ampicillin. | Other gram negatives | Ampicillin | 6/11 (55%) | 5/10 (50%) | 3/5 (60%) |  |
| Edwards et al. 2003 | Retrospective cohort study | Historical | USA | High | General hospital | CDC 1996. |  |  | Ampicillin prior to march 1995 and penicillin thereafter. | All: <7 days in blood. | Ampicillin |  | 11/34 (32%) | 25/41 (61%) |  |
| Edwards et al. 2003 | Retrospective cohort study | Historical | USA | High | General hospital | CDC 1996. |  |  | Ampicillin prior to march 1995 and penicillin thereafter. | GBS | Ampicillin |  | 0/14 (0%) | 0/9 (0%) |  |
| Edwards et al. 2003 | Retrospective cohort study | Historical | USA | High | General hospital | CDC 1996. |  |  | Ampicillin prior to march 1995 and penicillin thereafter. | Coagulase-negative staphylococcus | Ampicillin |  | 3/3 (100%) | 13/13 (100%) |  |
| Freitas & Romero 2017 | Retrospective cohort study | Historical | Brazil | High-Middle | Regional maternity hospital |  |  | Based on CDC 2010 | Ampicillin 2g IV initially and 1 g every 4 h. Penicillin allergy, clindamycin 900 mg IV every 8 hours | GBS: <72 hours in blood and antibiotic treatment ≥5 days or death <5 days while on antibiotic treatment. | Penicillin^1^  Ampicillin^2^  Vancomycin^3^ | 0/7 (0%)^1^  0/7 (0%)^2^  0/7 (0%)^3^ |  |  | - |
| Freitas & Romero 2017 | Retrospective cohort study | Historical | Brazil | High-Middle | Regional maternity hospital |  |  | Based on CDC 2010 | Ampicillin 2g IV initially and 1 g every 4 h. Penicillin allergy, clindamycin 900 mg IV every 8 hours | E. Coli | Ampicillin^1^  3^rd^ generation cephalosporins^2^  Gentamicin^3^ | 4/4 (100%)^1^  0/4 (0%)^2^  ?^3^ |  |  | 1/1 (100%)^1^  0/1 (0%)^2^  ?^3^ |
| Freitas & Romero 2017 | Retrospective cohort study | Historical | Brazil | High-Middle | Regional maternity hospital |  |  | Based on CDC 2010 | Ampicillin 2g IV initially and 1 g every 4 h. Penicillin allergy, clindamycin 900 mg IV every 8 hours | S. Aureus | Methicillin | 0/3 (0%) |  |  | 0/1 (0%) |
| Freitas & Romero 2017 | Retrospective cohort study | Historical | Brazil | High-Middle | Regional maternity hospital |  |  | Based on CDC 2010 | Ampicillin 2g IV initially and 1 g every 4 h. Penicillin allergy, clindamycin 900 mg IV every 8 hours | E. Faecalis | Vancomycin | 0/2 (0%) |  |  | - |
| Lu et al. 2022 | Retrospective cohort study | Historical | Taiwan | High | University hospital with a level 3 NICU | IAP to: carriers of rectovaginal GBS culture at 35-37 weeks' gestation. |  |  | NI. | E. Coli: <72 hours in blood or CSF. | Ampicillin^1^  Gentamicin^2^ | 6/10 (60%)^1^  3/10 (30%)^2^ |  | 15/16 (94%)^1^  9/16 (56%)^2^ |  |
| O'Sullivan et al. 2019 | Prospective active national surveillance study | Historical | UK and Ireland | High | Active surveillance (all paediatricians) and laboratory databases (all) |  | RCOG 2003, NICE 2012. |  | Penicillin G, cephalosporin or vancomycin. | GBS: <7 days in blood, CSF or joint fluid. | Penicillin^1^  Clindamycin^2^  Erythromycin^3^ | 0/377 (0%)^1^  ?^2^  ?^3^ | 0/517 (0%)^1^ ?^2^ ?^3^ |  |  |
| Phares et al. 2008 | Retrospective cohort study | Historical | USA | High | Database Active Bacterial Core surveillance/ Emerging Infections Program Network | CDC 2002. |  | CDC 1996, ACOG 1996, AAP 1997 | CDC 1996, ACOG 1996, AAP 1997, CDC 2002. | GBS: <7 days in blood or CSF. | Penicillin^1^  Ampicillin^2^  Vancomycin^3^  Erythromycin^4^  Clindamycin^5^ |  |  | 0/517 (0%)^1^  0/517 (0%)^2^  0/517 (0%)^3^  ?^4^  ?^5^ | 0/715 (0%)^1^  0/715 (0%)^2^  0/715 (0%)^3^  ?^4^  ?^5^ |
| Puopolo & Eichenwald 2010* | Retrospective cohort study | Historical | USA | High | University hospital | IAP to: carriers of rectovaginal colonisation at 35-37 weeks' gestation. | IAP to: PTL, PROM, fever. |  | Ampicillin or clindamycin for risk-factor strategy.  Penicillin G for universal strategy. Penicillin allergy, clindamycin, cefazolin, erythromycin and vancomycin. | All: <72 hours in blood and neonatologist considered infant infected. Surviving infants treated with appropriate course of antibiotics ≥7 days. | Ampicillin | 17/96 (18%) | 20/78 (26%) | 66/161 (41%) |  |
| Puopolo & Eichenwald 2010* | Retrospective cohort study | Historical | USA | High | University hospital | IAP to: carriers of rectovaginal colonisation at 35-37 weeks' gestation. | IAP to: PTL, PROM, fever. |  | Ampicillin or clindamycin for risk-factor strategy.  Penicillin G for universal strategy. Penicillin allergy, clindamycin, cefazolin, erythromycin and vancomycin. | E. Coli | Ampicillin | 9/15 (60%) | 5/16 (31%) | 22/40 (55%) |  |
| Puopolo & Eichenwald 2010* | Retrospective cohort study | Historical | USA | High | University hospital | IAP to: carriers of rectovaginal colonisation at 35-37 weeks' gestation. | IAP to: PTL, PROM, fever. |  | Ampicillin or clindamycin for risk-factor strategy.  Penicillin G for universal strategy. Penicillin allergy, clindamycin, cefazolin, erythromycin and vancomycin. | Gram-negative species | Ampicillin | 12/19 (63%) | 12/24 (50%) | 38/61 (62%) |  |
| Puopolo & Eichenwald 2010* | Retrospective cohort study | Historical | USA | High | University hospital | IAP to: carriers of rectovaginal colonisation at 35-37 weeks' gestation. | IAP to: PTL, PROM, fever. |  | Ampicillin or clindamycin for risk-factor strategy.  Penicillin G for universal strategy. Penicillin allergy, clindamycin, cefazolin, erythromycin and vancomycin. | Gram-positive species | Ampicillin | 5/77 (6.5%) | 7/60 (12%) | 26/98 (27%) |  |
| Sridhar et al. 2014 | Retrospective cohort study | Historical | India | Low-Middle | Tertiary level neonatal unit of teaching hospital |  | IAP to: PTL, (premature) PROM, fever, 3 or more gloved per vaginal examinations, chorioamnionitis, and GBS bacteriuria. |  | Ampicillin. | EOGBS | Penicillin | No resistance | No resistance |  |  |
| Sutkin et al. 2005 | Retrospective cohort study | Historical | USA | High | Tertiary referral hospital | CDC 1996. |  | AAP 1992 | Penicillin and clindamycin. | GBS: <48 hours in blood, CSF or other tissue. | Penicillin^1^  Ampicillin^2^  Erythromycin^3^  Clindamycin^4^ |  |  | 0/3 (0%)^1^  0/3 (0%)^2^  0/3 (0%)^3^  0/3 (0%)^4^ | 0/14 (0%)^1^  0/14 (0%)^2^  0/14 (0%)^3^  1/14 (7%)^4^ |
| Sutkin et al. 2005 | Retrospective cohort study | Historical | USA | High | Tertiary referral hospital | CDC 1996. |  | AAP 1992 | Penicillin and clindamycin. | E. Coli | Ampicillin^1^  Gentamicin^2^  Cefotaxime^3^ |  |  | 7/11 (64%)^1^  0/11 (0%)^2^  0/11 (0%)^3^ | 7/11 (64%)^1^  0/11 (0%)^2^  0/11 (0%)^3^ |
| Trijbels-Smeulders et al. 2006 | Retrospective cohort study | Historical | the Netherlands | High | 22 Laboratories for Medical Microbiology and from the Netherlands Reference Laboratory for Bacterial Meningitis in 51/93 neonatal and pediatric wards |  |  | Dutch guidelines 1999. | Dutch guidelines 1999 | GBS: ≤7 days in blood/CSF | Penicillin  Amoxicillin  Cefixime  Cefepime  Cripofloxacin  Ceftazidime  Trovafloxacin | No resistance |  |  | No resistance |
| van den Hoogen et al. 2010 | Retrospective cohort study | Historical | the Netherlands | High | University hospital level 3 NICU unit |  |  | Dutch guidelines 1999. | Dutch guidelines 1999 | GBS: <48 hours in blood and clinical signs of infection. | Penicillins and other beta-lactam antibiotics | 0/68 (0%) |  |  | 0/31 (0%) |
| Wang et al. 2023 | Retrospective cohort study | Historical | China | High-Middle | Eight public hospitals and 31 health centres | IAP to: i) carriers of rectovaginal GBS colonisation at 35-37 weeks' gestation, and ii) carrier state known before 35 weeks' gestation, GBS bacteriuria or history previous infant affected by GBS disease. | IAP to: previous infant with invasive GBS disease, PTL, GBS colonisation, and GBS bacteriuria. |  | Penicillin G 5 million IU IV initially and 2.5 million IU every 4 hours. Penicillin allergy, erythromycin, clindamycin, or vancomycin according to sensitivity. | GBS: <72 hours in blood or CSF. | Penicillin^1^ Ampicillin^2^ Clindamycin^3^ Erythromycin^4^ |  | 0/302 (0%)^1^ 0/302 (0%)^2^ 65/203 (32%)^3^ 110/203 (54%)^4^ | 0/54 (0%)^1^ 0/54 (0%)^2^ 27/54 (32%)^3^ 33/54 (61%)^4^ |  |
| Wang et al. 2023 | Retrospective cohort study | Historical | China | High-Middle | Eight public hospitals and 31 health centres | IAP to: i) carriers of rectovaginal GBS colonisation at 35-37 weeks' gestation, and ii) carrier state known before 35 weeks' gestation, GBS bacteriuria or history previous infant affected by GBS disease. | IAP to: previous infant with invasive GBS disease, PTL, GBS colonisation, and GBS bacteriuria. |  | Penicillin G 5 million IU IV initially and 2.5 million IU every 4 hours. Penicillin allergy, erythromycin, clindamycin, or vancomycin according to sensitivity. | S. Bovis: <72 hours in blood or CSF. | Penicillin^1^ Ampicillin^2^ Clindamycin^3^ Erythromycin^4^ |  | 0/17 (0%)^1^ 0/17 (0%)^2^ 5/17 (29%)^3^ 12/17 (71%)^4^ | 0/38 (0%)^1^ 0/38 (0%)^2^ 14/38 (37%)^3^ 23/38 (61%)^4^ |  |
| Wang et al. 2023 | Retrospective cohort study | Historical | China | High-Middle | Eight public hospitals and 31 health centres | IAP to: i) carriers of rectovaginal GBS colonisation at 35-37 weeks' gestation, and ii) carrier state known before 35 weeks' gestation, GBS bacteriuria or history previous infant affected by GBS disease. | IAP to: previous infant with invasive GBS disease, PTL, GBS colonisation, and GBS bacteriuria. |  | Penicillin G 5 million IU IV initially and 2.5 million IU every 4 hours. Penicillin allergy, erythromycin, clindamycin, or vancomycin according to sensitivity. | Other gram-positive bacteria: <72 hours in blood or CSF. | Penicillin^1^ Ampicillin^2^ Erythromycin^3^ |  | 0/45 (0%)^1^ 0/45 (0%)^2^ 14/45 (31%)^3^ | n.p. (23%)^1^ n.p. (12%)^2^ n.p. (12%)^3^ |  |
| Wang et al. 2023 | Retrospective cohort study | Historical | China | High-Middle | Eight public hospitals and 31 health centres | IAP to: i) carriers of rectovaginal GBS colonisation at 35-37 weeks' gestation, and ii) carrier state known before 35 weeks' gestation, GBS bacteriuria or history previous infant affected by GBS disease. | IAP to: previous infant with invasive GBS disease, PTL, GBS colonisation, and GBS bacteriuria. |  | Penicillin G 5 million IU IV initially and 2.5 million IU every 4 hours. Penicillin allergy, erythromycin, clindamycin, or vancomycin according to sensitivity. | E. Coli: <72 hours in blood or CSF. | Ampicillin^1^ Gentamicin^2^ Amoxicillin clavulanate^3^ Second-generation cephalosporins^4^ Third-generation cephalosporins^5^ Cefepime^6^ Carbapenems^7^ Amikacin^8^ Extended spectrum beta-lactamase^9^ |  | 48/77 (62%)^1^ 26/77 (34%)^2^ 7/77 (9%)^3^ 9/77 (12%)^4^ 8/77 (10%)^5^ n.p.^6^ 0/77 (0%)^7^ n.p.^8^ 8/77 (10%)^9^ | 64/79 (81%)^1^ 29/79 (37%)^2^ 11/79 (14%)^3^ 20/79 (25%)^4^ 17/79 (22%)^5^ 5/79 (6%)^6^ 0/79 (0%)^7^ 0/79 (0%)^8^ 11/79 (14%)^9^ |  |
| Wang et al. 2023 | Retrospective cohort study | Historical | China | High-Middle | Eight public hospitals and 31 health centres | IAP to: i) carriers of rectovaginal GBS colonisation at 35-37 weeks' gestation, and ii) carrier state known before 35 weeks' gestation, GBS bacteriuria or history previous infant affected by GBS disease. | IAP to: previous infant with invasive GBS disease, PTL, GBS colonisation, and GBS bacteriuria. |  | Penicillin G 5 million IU IV initially and 2.5 million IU every 4 hours. Penicillin allergy, erythromycin, clindamycin, or vancomycin according to sensitivity. | Other gram-negative bacteria: <72 hours in blood or CSF. | Ampicillin^1^ Gentamicin^2^ Amoxicillin clavulanate^3^ Second-generation cephalosporins^4^ Third-generation cephalosporins^5^ Cefepime^6^ Carbapenems^7^ Amikacin^8^ Extended spectrum beta-lactamase^9^ |  | n.p. (63%)^1^ n.p. (5%)^2^ n.p. (16%)^3^ n.p. (11%)^4^ n.p. (5%)^5^ n.p. (5%)^6^ n.p. (0%)^7^ n.p. (0%)^8^ n.p. (11%)^9^ | n.p. (71%)^1^ n.p. (12%)^2^ n.p. (24%)^3^ n.p. (24%)^4^ n.p. (18%)^5^ n.p. (0%)^6^ n.p. (12%)^7^ n.p. (6%)^8^ n.p. (6%)^9^ |  |
| ? Refers to unknown number of EOS isolates that are resistant in this period.  - Refers to no EOS isolates in this time period. * Refers to data from same hospital. n.p. Refers to unpublished data to maintain confidentiality of individuals in small groups. | | | | | | | | | | | | | | | |

**Table 6: Maternal peripartum infection**

| **Author Year** | **Study design** | **Time analysis** | **Country** | **Country Income** | **Study setting/ Data source** | **IAP criteria: Universal screening** | **IAP criteria:  Risk-factors** | **IAP criteria: Other strategies** | **IAP agent:** | **Infection** | **Maternal peripartum infection incidence (%)** | | | |
| --- | --- | --- | --- | --- | --- | --- | --- | --- | --- | --- | --- | --- | --- | --- |
|  |  |  |  |  |  |  |  |  |  |  | **No strategy** | **Risk strategy** | **Universal strategy** | **Other strategies** |
| Davis et al. 2001 | Retrospective cohort study | Historical | USA | High | Nonprofit group and network health maintenance organisation with data from 2 hospitals | CDC 1996. |  | AAP 1992 | Penicillin G | Amnionitis^1^ Endometritis^2^ Sepsis/bacteraemia^3^ Anaphylaxis^4^ |  |  | 44/2438 (1.8%)^1^ 14/2438 (0.6%)^2*^ 0/2438 (0%)^3**^ 0/2438 (0%)^4^ | 26/1337 (1.9%)^1^ 23/1337 (1.7%)^2*^ 2/1337 (0.2%)^3**^ 0/1337 (0%)^4^ |
| Gilson et al. 2000 | Retrospective cohort study | Concurrent | USA | High | Academic medical centre | IAP to: i) carriers of GBS colonisation at 35-37 weeks' gestation, and ii) if unknown carrier state, presence of risk-factors. | IAP to: PTL, fever, PROM, and previous infant with GBS infection. |  | Before march 1995, ampicillin 2 g IV every 6 h. After march 1995, penicillin G 5 million IU initially and 2.5 million IU every 4 h. Penicillin allergy, clindamycin 900 mg every 8 h. | Chorioamnionitis^1^ Endometritis^2^ |  | 23/407 (5.7%)^1^ 15/407 (3.9%)^2^ | 28/420 (6.7%)^1^ 11/420 (2.6%)^2^ |  |
| Jeffery & Moses Lahra 1998 | Prospective cohort study | Historical | Australia | High | Tertiary referral hospital | IAP to: i) carriers of vaginal GBS colonisation at 28 weeks' gestation without risk-factors or 24 weeks if known risk-factor for preterm birth, ii) GBS bacteriuria, iii) previous infant with EOGBS infection, and iv) if unknown carrier state, presence of preterm labour. |  |  | Ampicillin 1 g every 6 hours. Penicillin allergy, cephalosporin. | Anaphylaxis | 0/5732 (0%) |  | 1/36342 (0%) |  |
| Katz et al. 1999 | Retrospective cohort study | Historical | USA | High | Urban tertiary centre |  |  | IAP to: i) carriers of rectovaginal GBS culture at 28 weeks' gestation and PTL, and ii) (premature) PROM, fever and previous infant with GBS infection. | Ampicillin 2 g IV every 6 h. Penicillin allergy, clindamycin 600 mg every 6 h. | Chorioamnionitis | 578/15620 (3.7%)^*^ |  |  | 271/8748 (3.1%)^*^ |
| Locksmith et al. 1999 | Retrospective cohort study | Historical | USA | High | University tertiary care centre | CDC 1996. |  | Until 1993, IAP to: carriers of GBS colonisation at hospital with PTL, premature PROM and another risk-factor (unspecified). | Until 1995, ampicillin 2 g IV initially and 1 g every 6 h. After 1995, penicillin 5 million IU and 2.5 million IU every 4 h. Amoxicillin for premature PROM and GBS colonisation. | Chorioamnionitis^1^ Endometritis^2^ |  |  | 233/4453 (5.2%)^1*^ 123/4453 (2.8%)^2*^ | 575/7810 (7.4%)^1*^ 314/7810 (4.0%)^2*^ |
| Locksmith et al. 1999 | Retrospective cohort study | Historical | USA | High | University tertiary care centre | CDC 1996. |  | After 1993, ACOG guidelines but unknown which ones. IAP to: i) if unknown carrier state, presence of PTL, PROM, fever, and previous infant with GBS sepsis, and ii) strategy above. | Until 1995, ampicillin 2 g IV initially and 1 g every 6 h. After 1995, penicillin 5 million IU and 2.5 million IU every 4 h. Amoxicillin for premature PROM and GBS colonisation. | Chorioamnionitis^1^ Endometritis^2^ |  |  | 233/4453 (5.2%)^1*^ 123/4453 (2.8%)^2*^ | 599/7917 (7.7%)^1^* 366/7917 (4.6%)^2^* |
| Uy et al. 2002 | Retrospective population survey | Historical | USA | High | Tertiary care centre, division of university hospital. |  | CDC 1996.  Adhere to risk-based guidelines, but prescription left to individual practitioner. | AAP 1992, ACOG 1992 | AAP 1992: Ampicillin 2 g IV initially, 1-2 g every 4-6 h or penicillin G 5 million IU every 6 h. Penicillin allergy, clindamycin or erythromycin.   Also ACOG 1992 and CDC 1996. | Chorioamnionitis | 10/300 (3.3%) | 6/150 (4.0%) |  | 4/150 (2.7%) |
| *p<0.05 **p<0.07 | | | | | | | | | | | | | | |
