## Supplementary file 3 for "Intrapartum antibiotic prophylaxis to prevent Group B streptococcal infections in newborn infants: a systematic review and meta-analysis comparing various strategies"

**Risk of Bias Assessment for individual studies (ROBINS-I)**

| **Author & Year** | **RoB: Confounding** | **Confounding explanation** | **RoB: participant selection** | **Participant selection explanation** | **RoB: Classification of intervention** | **Classification of intervention explanation** | **RoB: Deviations from intended intervention** | **Deviations from intended intervention explanation** | **RoB: Missing data** | **Missing data explanation** | **RoB: Outcome measurement** | **Outcome measurement explanation** | **RoB: Selection of reported results** | **Selection of reported results explanation** | **Overall RoB** | **Overall RoB explanation.** |
| --- | --- | --- | --- | --- | --- | --- | --- | --- | --- | --- | --- | --- | --- | --- | --- | --- |
| Abdelmaaboud & Mohammed 2011 | Serious | Provide demographic data for total population, but potential confounding because baseline characteristics differ between groups. No control for potential confounding (including time). | Low | Both patient populations from same demographic. | Low | Prospectively and well defined interventions. Outcomes determined after intervention. | Unknown | No information on protocol adherence. | Moderate | Retrospective data collection. | Moderate | Retrospective outcome measurement, but outcome measurement likely standardised. Outcome measures unaffected by knowledge of intervention. Methods of outcome assessment comparable because cases identified from same microbiology laboratory’s computerized database. | Critical | Outcomes correspond to standard incidence measures, however incidences do not correspond to total amount of cases. | Critical | This study has critical risk of bias in the domains of reported results. |
| Al Luhidan et al. 2019 | Serious | No demographic data total population and no control for potential confounding (including time) | Low | Both patient populations from same demographic. | Low | Prospectively and well defined interventions. Outcomes determined after intervention. | Unknown | No information on protocol adherence. | Moderate | Retrospective data collection. | Moderate | Retrospective outcome measurement, but outcome measurement likely standardised. Outcome measures unaffected by knowledge of intervention. Methods of outcome assessment comparable because cases identified from same microbiology laboratory database. | Low | Outcomes correspond to standard incidence measures and there were no specific subset of analyses is reported. | Serious | This study has serious risk of bias in the domain of confounding. |
| Alarcon et al. 2004 | Moderate | E. Coli EOS group described. No control for potential confounding (including time). | Low | Both patient populations from same demographic. | Low | Prospectively and well defined interventions. Outcomes determined after intervention. | Unknown | No information on protocol adherence. | Moderate | Missing data on 1/41 cases, unknown why missing, but in line with retrospective nature of study | Moderate | Outcome measures unaffected by knowledge of intervention. Methods of outcome assessment comparable because cases identified from same microbiologic register. Retrospective outcome measurement, but outcome measurement standardised. | Low | Outcomes correspond to standard resistance measures and there were no specific subset of analyses is reported. | Moderate | This study has moderate risk of bias in the domains of confounding, missing data and outcome measurement. |
| Andreu et al. 2003 | Serious | No demographic data total population and no control for potential confounding (including time) | Low | Both patient populations from same demographic. | Serious | Unclear what the criteria were for IAP administration. Outcomes determined after intervention. | Unknown | No information on protocol adherence. | Moderate | Retrospective data collection. | Moderate | Retrospective outcome measurement, but clear definition. Outcome measures unaffected by knowledge of intervention. Methods of outcome assessment comparable because cases identified from same database. | Low | Outcomes correspond to standard incidence measures and there were no specific subset of analyses is reported. | Serious | This study has serious risk of bias in the domains confounding and classification of interventions. |
| Angstetra et al. 2007 | Low | Provide demographic data for total population. Control for some potential confounding (not including time). | Low | Both patient populations from same demographic. | Serious | Interventions prospectively defined, but risk-based screening is undefined. Outcomes determined after intervention. | Unknown | No information on protocol adherence. | Low | Prospective data collection with outcome data available for all participants. | Moderate | Prospective outcome measurement. Outcome measures unaffected by knowledge of intervention. Methods of outcome assessment incomparable because cases identified from different databases. | Low | Outcomes correspond to standard incidence measures and there were no specific subset of analyses is reported. | Serious | This study has serious risk of bias in the domains of classification of interventions. |
| Bekker et al. 2014 | Moderate-serious | No demographic data total population, but control for potential confounding (including time). | Low | Both patient populations from same demographic. | Low | Prospectively and well defined interventions. Outcomes determined after intervention. | Unknown | No information on protocol adherence. | Low | Retrospective data collection with nationwide surveillance. Outcome data available for all participants. | Low | Nationwide surveillance. Outcome measures unaffected by knowledge of intervention. Methods of outcome assessment incomparable because cases identified from same databases. | Moderate-serious | Outcomes correspond to standard incidence measures and there were no specific subset of analyses is reported, though raw incidences are not publishes and incidences differ from those written in the paper. | Moderate-serious | This study has moderate-serious risk of bias in the domains of confounding. |
| Björklund et al. 2017 | Low | Provide some demographic data total population. No control for potential confounding, but short study duration makes time bias unlikely. | Low | Both patient populations from same demographic and close in time. | Low | Prospectively and well defined interventions. Outcomes determined after intervention. | Low | Adherence was only described in screening period, which was 89%. | Moderate | Retrospective data collection. | Moderate | Retrospective outcome measurement, but outcome measurement likely standardised. Outcome measures unaffected by knowledge of intervention. Methods of outcome assessment comparable because cases identified from clinical records database | Moderate | No data on adherence during risk-factor based period. Outcomes correspond to standard incidence measures and there were no specific subset of analyses is reported. | Moderate | This study has moderate risk of bias in the domains of missing data, outcome measurement and reported results. |
| Brozanski et al. 2000 | Serious | No demographic data on total population, but GBS colonisation similar. No control for potential confounding (including time). | Low | Both patient populations from same demographic. | Serious | Interventions prospectively defined, but the comparator group includes a combination strategy and no strategy. Outcomes determined after intervention. | Serious | Adherence was described and about 40% of all pregnant women received IAP without indication | Moderate | Retrospective data collection | Moderate | Retrospective outcome measurement, but outcome measurement likely standardised. Outcome measures unaffected by knowledge of intervention. Methods of outcome assessment comparable because cases identified from medical record database | Low | Outcomes correspond to standard incidence measures and there were no specific subset of analyses is reported. | Serious | This study has serious risk of bias in the domain of confounding, classification of interventions and deviations from intended interventions. |
| Chan et al. 2023 | Moderate-serious | No demographic data on total population, but GBS colonisation similar. Control for potential confounding (including time). | Low | Both patient populations from same demographic. | Low | Prospectively and well defined interventions. Outcomes determined after intervention. | Unknown | No information on protocol adherence. | Moderate | Retrospective data collection. | Moderate | Retrospective outcome measurement, but outcome measurement likely standardised. Outcome measures unaffected by knowledge of intervention. Methods of outcome assessment comparable because cases identified from same laboratory database and reporting/clinical systems. | Low | Outcomes correspond to standard incidence measures, and there were no specific subset of analyses is reported. | Moderate-serious | This study has moderate-serious risk of bias in the domains of confounding. |
| Chen et al. 2005 | Serious | No demographic data on EOGBS group. No control for potential confounding (including time). | Low | Both patient populations from same demographic. | Low | Prospectively and well defined interventions. Outcomes determined after intervention. | Unknown | No information on protocol adherence. | Moderate | Retrospective data collection | Moderate | Retrospective outcome measurement, but outcome measurement likely standardised. Outcome measures unaffected by knowledge of intervention. Methods of outcome assessment comparable because cases identified from same hospital microbiology database. | Low | Outcomes correspond to standard resistance measures and there were no specific subset of analyses is reported. | Serious | This study has serious risk of bias in the domain of confounding. |
| Chen et al. 2001 | Moderate | Provide demographic data on EOGBS group. No control for potential confounding (including time). | Low | Both patient populations from same demographic. | Low | Prospectively and well defined interventions. Outcomes determined after intervention. | Unknown | No information on protocol adherence. | Moderate | Retrospective data collection | Moderate | Retrospective outcome measurement, but outcome measurement likely standardised. Outcome measures unaffected by knowledge of intervention. Methods of outcome assessment comparable because cases identified from same hospital microbiology database. | Low | Outcomes correspond to standard resistance measures and there were no specific subset of analyses is reported. | Moderate | This study has moderate risk of bias in the domains of outcome measurement and missing data. |
| Coco 2002 | Low | Provide demographic data on total population and no relevant confounding is present for IAP administration | Low | Both patient populations from same demographic. | Moderate | Prospectively and well defined interventions. Outcomes is part of intervention. Antibiotic agent unknown | Low | Protocol adherence was very good (97%) in the risk-based group and pretty good (81%) in the universal screening group | Moderate | Retrospective data collection. | Moderate | Retrospective outcome measurement, but outcome measurement likely standardised. Outcome measures unaffected by knowledge of intervention. Methods of outcome assessment comparable because data of IAP collected from same delivery logs, chart reviews, and medical record reports. | Low | Outcomes correspond to standard incidence measures and there were no specific subset of analyses is reported. | Moderate | This study has moderate risk of bias in the domains of classification of intervention, deviations from intervention, missing data and outcome measurement. |
| Darlow et al. 2023 | Serious | No demographic data total population and no control for potential confounding (including time). | Low | Both patient populations from same demographic. | Low | Prospectively and well defined interventions. Outcomes determined after intervention. | Serious | Only 37% of pregnant women eligible for IAP received IAP. | Moderate | Prospective data collection. Authors admit they might have missed a few cases. | Low | Prospective surveillance of outcome measurement. Outcome measures unaffected by knowledge of intervention. Methods of outcome assessment comparable because cases identified from same surveys/databases. | Low | Outcomes correspond to standard incidence measures and there were no specific subset of analyses is reported. | Serious | This study has serious risk of bias in the domains of confounding and deviations from intended intervention. |
| Davis et al. 2001 | Moderate | Provide demographic data total population, which differ between groups. Short study duration makes time bias less likely. | Low | Both patient populations from same demographic. | Low | Prospectively and well defined interventions. Outcomes determined after intervention. | Moderate | Protocol adherence was pretty good, because 74% of carriers were treated | Moderate | Retrospective data collection. Estimates of intrapartum antibiotic prophylaxis are somewhat incomplete and rate probably higher than shown. | Moderate | Retrospective outcome measurement, but outcome measurement likely standardised. Outcome measures unaffected by knowledge of intervention. Methods of outcome assessment comparable because antibiotic administration is registered in same automated databases. | Low | Outcomes correspond to standard incidence measures and there were no specific subset of analyses is reported. | Moderate | This study has moderate risk of bias in the domains of confounding, deviations from intended interventions, missing data and outcome measurement. |
| Eberly & Rajnik 2009 | Serious | No demographic data on total population and no control for potential confounding (including time). | Low | Both patient populations from same demographic. | Low | Prospectively and well defined interventions. Outcomes determined after intervention. | Unknown | No information on protocol adherence. | Moderate | Retrospective data collection. | Moderate | Retrospective outcome measurement, but outcome measurement likely standardised by means of data codes. Outcome measures unaffected by knowledge of intervention. Methods of outcome assessment comparable because cases identified from same data record. | Low | Outcomes correspond to standard incidence measures and there were no specific subset of analyses is reported. | Serious | This study has serious risk of bias in the domain of confounding. |
| Ecker et al. 2013 | Serious | No demographic data total population and no control for potential confounding (including time). | Low | Both patient populations from same demographic. | Serious | Interventions prospectively defined, but risk-based screening is not defined. Outcomes determined after intervention. | Unknown | No information on protocol adherence. | Moderate | Retrospective data collection. | Moderate | Retrospective outcome measurement, but outcome measurement likely standardised. Outcome measures unaffected by knowledge of intervention. Methods of outcome assessment comparable because cases identified from same microbiology laboratory database. | Low | Outcomes correspond to standard incidence measures and there were no specific subset of analyses is reported. | Serious | This study has serious risk of bias in the domains of confounding and classification of intervention. |
| Edwards et al. 2003 | Serious | No demographic data total population and no control for potential confounding (including time). | Low | Both patient populations from same demographic. | Serious | Interventions prospectively defined, but risk-based screening is not defined and different antibiotics used in the study. Outcomes determined after intervention. | Unknown | No information on protocol adherence. | Moderate | Retrospective data collection. | Moderate | Retrospective outcome measurement, but outcome measurement likely standardised. Outcome measures unaffected by knowledge of intervention. Methods of outcome assessment comparable because cases identified from laboratory database. | Low | Outcomes correspond to standard incidence measures and there were no specific subset of analyses is reported. | Serious | This study has serious risks of bias in the domains of confounding and classification of intervention. |
| Eisenberg et al. 2005 | Moderate | Provide demographic data total population. No control for potential confounding (including time) | Low | Both patient populations from same demographic. | Moderate | Interventions determined on the basis of GBS culture within 2 days, which was retrospectively determined. Outcomes determined after intervention. | Moderate | Adherence was only 52% with at least one risk factor. | Moderate | Retrospective data collection. | Low | Retrospective outcome measurement, but active surveillance. Outcome measures unaffected by knowledge of intervention. Methods of outcome assessment comparable because cases identified from birth registry data. | Low | Outcomes correspond to standard incidence measures and there were no specific subset of analyses is reported. | Moderate | This study has moderate risk of bias in the domains of confounding, deviations from intended intervention and missing data. |
| El Helali et al. 2019 | Moderate | Provide demographic data total population, but no control for potential confounding (including time). | Low | Both patient populations from same demographic. | Low | Prospectively and well defined interventions. Outcomes determined after intervention. | Low | Antenatal adherence was 89% and intrapartum adherence was 92%. | Moderate | Retrospective data collection. | Moderate | Retrospective outcome measurement, but outcome measurement likely standardised. Outcome measures unaffected by knowledge of intervention. Methods of outcome assessment comparable because antibiotic administration is registered in same microbiology laboratory. | Low | Outcomes correspond to standard incidence measures and there were no specific subset of analyses is reported. | Moderate | This study has moderate risk of bias in the domains of confounding, missing data and outcome measurement. |
| Factor et al. 1998 | Low | Provide demographic data total population and control for potential confounding (not including time and preterm delivery). | Low | Both patient populations from same demographic. | Low | Prospectively and well defined interventions. Outcomes determined after intervention. | Moderate | Adherence ranged between 42-72% per risk factor during the risk factor based screening protocol. | Moderate | Retrospective data collection. | Low | Retrospective outcome measurement, but surveillance. Outcome measures unaffected by knowledge of intervention. Methods of outcome assessment comparable because cases identified from same computerised database. | Low | Outcomes correspond to standard incidence measures and there were no specific subset of analyses is reported. | Moderate | This study has moderate risk of bias in the domains of deviation from intended intervention and missing data. |
| Freitas & Romero 2017 | Serious | No demographic data total population and no control for potential confounding (including time). | Low | Both patient populations from same demographic. | Low | Prospectively and well defined interventions. Outcomes determined after intervention. | Moderate | Adherence was only 55% for the screening strategy period. | Moderate | Retrospective data collection. | Moderate | Retrospective outcome measurement, but outcome measurement likely standardised. Outcome measures unaffected by knowledge of intervention. Methods of outcome assessment comparable because cases identified from same microbiology laboratory database. | Low | Outcomes correspond to standard incidence measures and there were no specific subset of analyses is reported. | Serious | This study has serious risk of bias in the domain of confounding. |
| Garland 1991 | Low | Provide demographic data for total population and concurrent controls. | Moderate | Patient populations from different groups. | Low | Prospectively and well defined interventions. Outcomes determined after intervention. | Unknown | No information on protocol adherence. | Moderate | Retrospective data collection. | Serious | Retrospective outcome measurement and unclear time definition of EOGBS. Outcome measures unaffected by knowledge of intervention. Methods of outcome assessment unknown. | Low | Outcomes correspond to standard incidence measures and there were no specific subset of analyses is reported. | Serious | This study has serious risk of bias the domain of outcome measurement. |
| Gibbs et al. 1994 | Serious | No demographic data for time period without screening strategy. No control for potential confounding (including time). | Low | Both patient populations from same demographic. | Low | Prospectively and well defined interventions. Outcomes determined after intervention. | Low | High protocol compliance (76-82%). | Moderate | Retrospective data collection. | Serious | Retrospective outcome measurement and unclear definition of EOGBS. Outcome measures unaffected by knowledge of intervention. Outcome assessment unknown. | Low | Outcomes correspond to standard incidence measures and there were no specific subset of analyses is reported. | Serious | This study has serious risk of bias in the domains of confounding and outcome measurement. |
| Gilson et al. 2000 | Low | Provide demographic data for total population and concurrent controls. | Moderate | Patient populations from different groups. | Low | Prospectively and well defined interventions, Outcomes determined after intervention. | Moderate | 72% adherence assessed in screening group. | Moderate | Retrospective data collection. | Moderate | Retrospective outcome measurement, but outcome measurement likely standardised. Outcome measures unaffected by knowledge of intervention. Methods of outcome assessment comparable because cases identified from same neonatology database for diagnosis. | Low | Outcomes correspond to standard incidence measures and there were no specific subset of analyses is reported. | Moderate | This study has moderate risk of bias in the domains of participation selection, deviations from intended intervention, missing data and outcome measurement. |
| Gopal Rao et al. 2017 | Moderate | Provide demographic data for total population, which differ slightly between groups. Controlled for potential confounding (risk of time bias low due to cross-over design). | Low | Both patient populations from same demographic. | Low | Prospectively and well defined interventions. Outcomes determined after intervention. | Low | Information only on screening, adherence 81%. | Moderate | Retrospective data collection. | Low | Retrospective outcome measurement, but active surveillance. Outcome measures unaffected by knowledge of intervention. Methods of outcome assessment comparable because cases identified from same microbiology laboratory records. | Low | Outcomes correspond to standard incidence measures and there were no specific subset of analyses is reported. | Moderate | This study has moderate risk of bias in the domains of confounding and missing data. |
| Gosling et al. 2002 | Serious | No demographic data total population, but concurrent controls. No control for potential confounding | Serious | Patient populations from significantly different populations (hospitals). | Moderate | Interventions not specifically defined. Outcomes determined after intervention. | Unknown | No information on protocol adherence. | Low | Retrospective data collection and outcome data likely available for all participants | Moderate | Retrospective outcome measurement, but via questionnaire survey. Outcome measures unaffected by knowledge of intervention. Outcome assessment unknown. | Low | Outcomes correspond to standard incidence measures and there were no specific subset of analyses is reported. | Serious | This study has serious risk of bias in the domains of confounding and selection of participants. |
| Hafner et al. 1998 | Moderate | Provide demographic data total population, but no control for potential confounding (including time). | Low | Both patient populations from same demographic. | Low | Prospectively and well defined interventions. Outcomes determined after intervention. | Low | Information only on screening, adherence 91%. | Moderate | Retrospective data collection. | Serious | Retrospective outcome measurement, but unclear time definition. Outcome measures unaffected by knowledge of intervention. Methods of outcome assessment different before and after implementation. | Low | Outcomes correspond to standard incidence measures and there were no specific subset of analyses is reported. | Serious | This study has serious risk of bias in the domain of outcome assessment. |
| Håkansson et al. 2017 | Moderate | Provide demographic data total population, but no control for potential confounding (including time). | Low | Both patient populations from same demographic. | Moderate | Prospectively and well defined interventions. Outcomes determined after intervention. Antibiotic agent unknown | Moderate | 33% of delivery units already had practice in use before official recommendation. No actual information on protocol adherence. | Moderate | Retrospective data collection, but outcome data available for most participants. | Low | Outcome measures unaffected by knowledge of intervention. Methods of outcome assessment comparable because cases identified from national registry | Low | Outcomes correspond to standard incidence measures and there were no specific subset of analyses is reported. | Moderate | This study has moderate risk of bias in the domains of confounding, classification of interventions, deviations from intended interventions and outcome measurement. |
| Hong et al. 2019 | Serious | Provide demographic data total population, which differ between groups. No control for these potential confounders, but short study duration makes time bias less likely. | Low | Both patient populations from same demographic. | Low | Prospectively and well defined interventions. Outcomes determined after intervention. | Low | Adherence was 98.6% in the universal screening strategy period. Adherence was not described in the risk-factor based screening period. | Moderate | Retrospective data collection. | Moderate | Retrospective outcome assessment, but clear definition. Outcome measures unaffected by knowledge of intervention. Methods of outcome assessment comparable because cases identified from same database. | Low | Outcomes correspond to standard incidence measures and there were no specific subset of analyses is reported. | Serious | This study has serious risk of bias in the domain of confounding. |
| Horvath et al. 2013 | Serious | No demographic data total population and no control for potential confounding (including time). | Low | Both patient populations from same demographic. | Low | Prospectively and well defined interventions. Outcomes determined after intervention. | Unknown | No information on protocol adherence. | Low | Prospective data collection with outcome data available for all participants. | Moderate | Prospective outcome measurement. Outcome measures unaffected by knowledge of intervention. Methods of outcome assessment unknown. | Low | Outcomes correspond to standard incidence measures and there were no specific subset of analyses is reported. | Serious | This study has serious risk of bias in the domain of confounding. |
| Isaacs & Royle 1999 | Serious | No demographic data total population and no control for potential confounding (including time). | Moderate | Patient populations from different groups. | Low | Prospectively and well defined interventions. Outcomes determined after intervention. | Unknown | No information on protocol adherence. | Low | Prospective data collection with outcome data available for all participants. | Low | Prospective outcome measurement. Outcome measures unaffected by knowledge of intervention. Methods of outcome assessment comparable because cases identified from same study group database. | Low | Outcomes correspond to standard incidence measures and there were no specific subset of analyses is reported. | Serious | This study has serious risk of bias in the domain of confounding. |
| Jeffery & Moses Lahra 1998 | Serious | No demographic data total population and no control for potential confounding (including time). | Low | Both patient populations from same demographic. | Low | Prospectively and well defined interventions. Outcomes determined after intervention. | Unknown | No information on protocol adherence. | Low | Prospective data collection with outcome data available for all participants. | Low | Prospective outcome measurement. Outcome measures unaffected by knowledge of intervention. Methods of outcome assessment comparable because cases identified from same clinical records. | Low | Outcomes correspond to standard incidence measures and there were no specific subset of analyses is reported. | Serious | This study has serious risk of bias in the domain of confounding. |
| Johansson Gudjónsdóttir et al. 2019 | Serious | No demographic data total population and no control for potential confounding (including time). | Low | Both patient populations from same demographic. | Moderate | Prospectively and well defined interventions. Outcomes determined after intervention. Antibiotic agent unknown | Unknown | No information on protocol adherence. | Moderate | Retrospective data collection. | Moderate | Retrospective outcome measurement, but outcome measurement likely standardised. Outcome measures unaffected by knowledge of intervention. Methods of outcome assessment comparable because cases identified from same study group database. | Moderate | Outcomes correspond to standard incidence measures, however raw incidences were not reported. | Serious | This study has serious risk of bias in the domain of confounding. |
| Katz et al. 1994 | Serious | No demographic data total population and no control for potential confounding (including time). | Low | Both patient populations from same demographic. | Low | Prospectively and well defined interventions. Outcomes determined after intervention. | Low | Adherence was 92% in the universal screening strategy period. | Moderate | Retrospective data collection. | Moderate | Retrospective outcome measurement, but outcome measurement likely standardised. Outcome measures unaffected by knowledge of intervention. Methods of outcome assessment comparable because cases identified from same microbiology laboratory. | Low | Outcomes correspond to standard incidence measures and there were no specific subset of analyses is reported. | Serious | This study has serious risk of bias in the domain of confounding. |
| Katz et al. 1999 | Serious | Provide demographic data total population, which differ between groups. No control for potential confounding (including time). | Low | Both patient populations from same demographic. | Low | Prospectively and well defined interventions. Outcomes determined after intervention. | Unknown | No information on protocol adherence. | Moderate | Retrospective data collection. | Moderate | Retrospective outcome measurement, but outcome measurement likely standardised. Outcome measures unaffected by knowledge of intervention. Methods of outcome assessment comparable because cases identified from same medical records chart database. | Low | Outcomes correspond to standard incidence measures and there were no specific subset of analyses is reported. | Serious | This study has serious risk of bias in the domain of confounding. |
| Lee et al. 2021 | Serious | No demographic data total population and no control for potential confounding (including time). | Low | Both patient populations from same demographic. | Low | Prospectively and well defined interventions. Outcomes determined after intervention. | Serious | Considerable overlap between risk-based or screening-based strategy period, therefore comparison risk of bias. | Moderate | Retrospective data collection. | Moderate | Retrospective outcome measurement, but outcome measurement likely standardised. Outcome measures unaffected by knowledge of intervention. Methods of outcome assessment comparable because cases identified from same laboratory database. | Low | Outcomes correspond to standard incidence measures and there were no specific subset of analyses is reported. | Serious | This study has serious risk of bias in the domains of confounding and protocol adherence. |
| Levine et al. 1999 | Serious | No demographic data total population and no control for potential confounding (including time). | Low | Both patient populations from same demographic. | Low | Prospectively and well defined interventions. Outcomes determined after intervention. | Unknown | No information on protocol adherence. | Moderate | Retrospective data collection. | Low | Retrospective outcome measurement, but active surveillance. Outcome measures unaffected by knowledge of intervention. Methods of outcome assessment comparable because cases identified from the same Infection Control Surveillance database. | Low | Outcomes correspond to standard incidence measures and there were no specific subset of analyses is reported. | Serious | This study has serious risk of bias in the domain of confounding. |
| Lin et al. 2011 | Moderate-serious | Provide demographic data only for universal screening group. Trend analysis without controlling for possible confounding. | Low | Both patient populations from same demographic. | Low | Prospectively and well defined interventions. Outcomes determined after intervention. | Low | 90% of GBS screen-positive pregnant women received IAP. | Moderate | Retrospective data collection. | Moderate | Retrospective outcome measurement, but clear outcome definition. Outcome measures unaffected by knowledge of intervention. Methods of outcome assessment comparable because cases identified from medical charts. | Low | Outcomes correspond to standard incidence measures and there were no specific subset of analyses is reported. | Moderate-serious | This study has moderate-serious risk of bias in the domain of confounding. |
| Locksmith et al. 1999 | Moderate | Provide demographic data for total population, which differ between groups. Control for potential confounding (not including time). | Low | Both patient populations from same demographic. | Low | Prospectively and well defined interventions. Outcomes determined after intervention. | Unknown | No information on protocol adherence of total population, only for EOGBS group. | Moderate | Retrospective data collection. | Low | Retrospective outcome measurement, but active surveillance. Outcome measures unaffected by knowledge of intervention. Methods of outcome assessment comparable because cases identified from same databases.. | Low | Outcomes correspond to standard incidence measures and there were no specific subset of analyses is reported. | Moderate | This study has moderate risk of bias in the domains of confounding and missing data. |
| López Sastre et al. 2005 | Serious | No demographic data total population and no control for potential confounding (including time). | Low | Both patient populations from same demographic. | Low | Prospectively and well defined interventions. Outcomes determined after intervention. | Unknown | No information on protocol adherence. | Low | Prospective data collection with outcome data available for all participants. | Low | Prospective surveillance of outcome measure. Outcome measures unaffected by knowledge of intervention. Methods of outcome assessment comparable because cases identified from same databases. | Low | Outcomes correspond to standard incidence measures and there were no specific subset of analyses is reported. | Serious | This study has serious risk of bias in the domain of confounding. |
| Lu et al. 2022 | Moderate | Provide demographic data on EOGBS group, but no control for potential confounding (including time). | Low | Both patient populations from same demographic. | Moderate | Prospectively and well defined interventions. Outcomes determined after intervention. Antibiotic agent unknown | Unknown | No information on protocol adherence. | Moderate | Retrospective data collection. | Serious | Retrospective outcome measurement, but outcome measurement likely not standardised. Outcome measures unaffected by knowledge of intervention. Methods of outcome assessment comparable because cases identified from microbiology database | Low | Outcomes correspond to standard antibiotic resistance measures and there were no specific subset of analyses reported. | Serious | This study has serious risk of bias in outcome assessment. |
| Lukacs & Schrag 2012 | Moderate-serious | No demographic data total population, but control for potential confounding (including time). | Low | Both patient populations from same demographic. | Low | Prospectively and well defined interventions. Outcomes determined after intervention. | Unknown | No information on protocol adherence. | Moderate | Cross sectional data collection. | Low | Cross sectional outcome measurement, but active surveillance. Outcome measures unaffected by knowledge of intervention. Methods of outcome assessment comparable because cases identified from same National Hospital Discharge Survey. | Low | Outcomes correspond to standard incidence measures and there were no specific subset of analyses is reported. | Moderate-serious | This study has moderate-serious risk of bias in the domains of confounding. |
| Main & Slagle 2000 | Moderate | Provide demographic data for total population, differ slightly between groups. No control for potential confounding (including time). | Low | Both patient populations from same demographic. | Low | Prospectively and well defined interventions. Outcomes determined after intervention. | Low | Adherence was 94% in the universal screening period. | Low | Prospective data collection with outcome data available for all participants. | Low | Prospective surveillance of outcome measure. Outcome measures unaffected by knowledge of intervention. Methods of outcome assessment comparable because cases identified from same combined mother-baby comprehensive perinatal database | Low | Outcomes correspond to standard incidence measures and there were no specific subset of analyses is reported. | Moderate | This study has moderate risk of bias in the domain of confounding. |
| Matsubara et al. 2007 | Moderate | Provide demographic data on total population, but no control for potential confounding (including time). | Serious | Patient populations from significantly different populations (hospitals). | Low | Prospectively and well defined interventions. Outcomes determined after intervention. | Unknown | No information on protocol adherence. | Moderate | Retrospective data collection. | Moderate | Retrospective outcome measurement, but via questionnaire survey. Outcome measures unaffected by knowledge of intervention. Outcome assessment unknown. | Low | Outcomes correspond to standard incidence measures and there were no specific subset of analyses is reported. | Serious | This study has serious risk of bias in the domain of selection of participants. |
| Matsubara et al. 2013 | Serious | No demographic data total population and no control for potential confounding (including time). | Low | Both patient populations from same demographic. | Low | Prospectively and well defined interventions. Outcomes determined after intervention. | Unknown | No information on protocol adherence. | Moderate | Retrospective data collection. | Low | Retrospective outcome measurement, but active surveillance. Outcome measures unaffected by knowledge of intervention. Methods of outcome assessment comparable because cases identified from discharge register of each hospital. | Low | Outcomes correspond to standard incidence measures and there were no specific subset of analyses is reported. | Serious | This study has serious risk of bias for the domain of confounding. |
| O'Sullivan et al. 2019 | Serious | No demographic data total population and no control for potential confounding (including time). | Low | Both patient populations from same demographic. | Low | Prospectively and well defined interventions. Outcomes determined after intervention. | Unknown | No information on protocol adherence. | Low | Prospective data collection with outcome data available for all participants. | Low | Prospective active surveillance. Outcome measures unaffected by knowledge of intervention. Methods of outcome assessment comparable because cases identified from the Public Health England national electronic surveillance database. | Low | Outcomes correspond to standard incidence measures and there were no specific subset of analyses is reported. | Serious | This study has serious risk of bias in the domain of confounding. |
| Petersen et al. 2014 | Moderate | Provide demographic data on total population, but no control for potential confounders (including time). | Low | Both patient populations from same demographic. | Moderate | Prospectively and well defined interventions. Outcomes determined after intervention. Antibiotic agent unknown | Unknown | No information on protocol adherence. | Moderate | Retrospective data collection. | Moderate | Retrospective outcome measurement, but outcome measurement likely standardised. Outcome measures unaffected by knowledge of intervention. Methods of outcome assessment comparable because cases identified from the Clinical Microbiology Database and a local database at the hospital. | Low | Outcomes correspond to standard incidence measures and there were no specific subset of analyses is reported. | Moderate | This study has moderate risk of bias in the domains of confounding, classification of interventions, missing data and outcome measurement. |
| Phares et al. 2008 | Serious | No demographic data total population and no control for potential confounding (including time). | Low | Both patient populations from same demographic. | Serious | Unknown what interventions are actually compared. | Unknown | No information on protocol adherence. | Moderate | Retrospective data collection. | Low | Retrospective outcome measurement, but active surveillance. Outcome measures unaffected by knowledge of intervention. Methods of outcome assessment comparable because cases identified from same medical record review and laboratory surveillance database. | Moderate | Outcomes correspond to standard incidence measures, however raw incidences were not reported. | Serious | This study has serious risk of bias in the domains of confounding and classification of intervention. |
| Poulain et al. 1997 | Serious | No demographic data total population and no control for potential confounding (including time). | Low | Both patient populations from same demographic. | Low | Prospectively and well defined interventions. Outcomes determined after intervention. | Moderate | In screening strategy period, 63% adherence | Moderate | Retrospective data collection. | Serious | Retrospective outcome measurement, but outcome measurement likely standardised. Outcome measures measured only when colonised mother. Methods of outcome assessment detailed, but strict definition unknown. Same databases used for case identification. | Low | Outcomes correspond to standard incidence measures and there were no specific subset of analyses is reported. | Serious | This study has serious risk of bias in the domains of confounding and outcome measurement. |
| Puopolo & Eichenwald 2010 | Serious | No demographic data total population and no control for potential confounding (including time). | Low | Both patient populations from same demographic. | Low | Prospectively and well defined interventions. Outcomes determined after intervention. | Unknown | No information on protocol adherence. | Moderate | Retrospective data collection. | Moderate | Retrospective outcome measurement, but outcome measurement likely standardised. Outcome measures unaffected by knowledge of intervention. Methods of outcome assessment comparable because cases identified from same microbiology laboratory electronic database. | Low | Outcomes correspond to standard incidence measures and there were no specific subset of analyses is reported. | Serious | This study has serious risk of bias in the domain of confounding. |
| Renner et al. 2006 | Serious | No demographic data total population and no control for potential confounding (including time). | Low | Both patient populations from same demographic. | Moderate | Prospectively and well defined interventions. Outcomes determined after intervention. Antibiotic agent unknown | Unknown | No information on protocol adherence. | Moderate | Retrospective data collection. | Moderate | Retrospective outcome measurement, but outcome measurement likely standardised. Outcome measures unaffected by knowledge of intervention. Methods of outcome assessment comparable because cases identified from same databases. | Low | Outcomes correspond to standard incidence measures and there were no specific subset of analyses is reported. | Serious | This study has serious risk of bias in the domain of confounding. |
| Rottenstreich et al. 2019 | Serious | No demographic data total population and no control for potential confounding (including time). | Low | Both patient populations from same demographic. | Moderate | Prospectively and well defined interventions. Outcomes determined after intervention. Antibiotic agent unknown | Unknown | No information on protocol adherence. | Moderate | Retrospective data collection. | Moderate | Retrospective outcome measurement, but outcome measurement likely standardised. Outcome measures unaffected by knowledge of intervention. Methods of outcome assessment comparable because cases identified from same computerized medical record database. | Low | Outcomes correspond to standard incidence measures and there were no specific subset of analyses is reported. | Serious | This study has serious risk of bias in the domain of confounding. |
| Sagrera et al. 2001 | Serious | No demographic data total population and no control for potential confounding (including time). | Low | Both patient populations from same demographic. | Low | Prospectively and well defined interventions. Outcomes determined after intervention. | Low | In universal screening period, compliance 93-94% | Moderate | Retrospective data collection. | Moderate | Retrospective outcome measurement, but clear outcome definition. Outcome measures unaffected by knowledge of intervention. Methods of outcome data assessment unknown . | Low | Outcomes correspond to standard incidence measures and there were no specific subset of analyses is reported. | Serious | This study has serious risk of bias in the domain of confounding. |
| Sakata 2012 | Serious | No demographic data total population and no control for potential confounding (including time). | Low | Both patient populations from same demographic. | Low | Prospectively and well defined interventions. Outcomes determined after intervention. | Unknown | No information on protocol adherence. | Moderate | Retrospective data collection. | Moderate | Retrospective outcome measurement, but outcomes clearly defined. Outcome measures unaffected by knowledge of interventions. Methods of outcome assessment comparable because cases identified from same medical record database. | Low | Outcomes correspond to standard incidence measures and there were no specific subset of analyses is reported. | Serious | This study has serious risk of bias in the domain of confounding. |
| Schrag et al. 2002 | Low | Provide demographic data on total population and control for relevant confounders. Concurrent control lower risk of time bias. | Low | Both patient populations from same demographic. | Moderate | Definition of both not really accurate, but methods to increase precision help define interventions. Outcomes determined after interventions, but unknown which antibiotic is given. | Moderate | In screening group adherence 89% and in risk-based group 61% | Moderate | Retrospective data collection, of which 95% of selected births had abstracted charts. | Moderate | Retrospective outcome measurement, but active surveillance. Outcome measures unaffected by knowledge of intervention. Methods of outcome assessment comparable because cases identified from same surveillance database. Unknown timing of EOGBS. | Low | Outcomes correspond to standard incidence measures and there were no specific subset of analyses is reported. | Moderate | This study has moderate risk of bias in the domains of classification of interventions, deviations from intended outcome, missing data and outcome measurement. |
| Schuchat et al. 2002 | Low | Provide demographic data on total population and no relevant confounding is present for IAP administration | Low | Both patient populations from same demographic. | Low | Prospectively and well defined interventions. Outcome is part of intervention. | Moderate | Universal screening adherence 74%, but 45% in risk-based strategy | Moderate | Retrospective data collection, of which 87.6% of selected births had abstracted charts. | Moderate | Retrospective outcome measurement, but outcomes likely standardised. Outcome measures =intervention. Methods of outcome assessment comparable because cases identified from maternal records. | Low | Outcomes correspond to antibiotic administration measures and there were no specific subset of analyses is reported. | Moderate | This study has moderate risk of bias in deviations from intended intervention, missing data and outcome measurement. |
| Share et al. 2001 | Serious | Provide demographic data total population, which differ between groups. No control for potential confounding (including time). | Low | Both patient populations from same demographic. | Low | Prospectively and well defined interventions. Outcomes determined after intervention. | Unknown | No information on protocol adherence. | Moderate | Retrospective data collection. | Moderate | Retrospective outcome measurement, but outcome clearly defined. Outcome measures unaffected by knowledge of intervention. Methods of outcome assessment comparable because cases identified from chart review. | Low | Outcomes correspond to standard incidence measures and there were no specific subset of analyses is reported. | Serious | This study has serious risk of bias in the domain of confounding. |
| Simetka et al. 2010 | Moderate | Provide demographic data on total population, but no control for potential confounders (including time). | Low | Both patient populations from same demographic. | Low | Prospectively and well defined interventions. Outcomes determined after intervention. | Unknown | No information on protocol adherence. | Moderate | Retrospective data collection. | Moderate | Retrospective outcome measurement, but outcome clearly defined. Outcome measures unaffected by knowledge of intervention. Outcome assessment likely standardised. | Low | Outcomes correspond to standard incidence measures and there were no specific subset of analyses is reported. | Moderate | This study has moderate risk of bias in the domains of confounding and outcome measurement. |
| Sridhar et al. 2014 | Serious | No demographic data on EOGBS group. No control for potential confounding (including time). | Low | Both patient populations from same demographic. | Low | Prospectively and well defined interventions. Outcomes determined after intervention. | Unknown | No information on protocol adherence. | Moderate | Retrospective data collection. | Moderate | Retrospective outcome measurement, but outcome measurement likely standardised. Outcome measures unaffected by knowledge of intervention. Methods of outcome assessment comparable because cases identified from hospital case records | Serious | No number of EOGBS cases provided. | Serious | This study has serious risk of bias in the domains of confounding and reporting results. |
| Sutkin et al. 2005 | Serious | Provide demographic data on total population, which differ between groups. No control for potential confounders (including time) | Low | Both patient populations from same demographic. | Low | Prospectively and well defined interventions. Outcomes determined after intervention. | Unknown | No information on protocol adherence. | Moderate | Retrospective data collection. | Moderate | Retrospective outcome measurement, but outcome measurement likely standardised. Outcome measures unaffected by knowledge of intervention. Methods of outcome assessment comparable because cases identified from microbiology database. | Low | Outcomes correspond to standard incidence measures and there were no specific subset of analyses is reported. | Serious | This study has serious risk of bias in the domain of confounding. |
| Tapia et al. 2007 | Moderate | Provide demographic data on total population, but no control for potential confounders (including time). | Low | Both patient populations from same demographic. | Moderate | Prospectively and well defined interventions. Outcomes determined after intervention. Antibiotic agent unknown | Unknown | No information on protocol adherence. | Moderate | Retrospective data collection. | Moderate | Retrospective outcome measurement, but likely standardised and outcomes clearly defined. Outcome measures unaffected by knowledge of intervention. Methods of outcome assessment comparable because cases identified from same clinical records database. | Low | Outcomes correspond to standard incidence measures and there were no specific subset of analyses is reported. | Moderate | This study has moderate risk of bias in the domains of confounding, classification of intervention, missing data and outcome measurement. |
| Towers & Briggs 2002 | Serious | No demographic data total population and no control for potential confounding (including time). | Low | Both patient populations from same demographic. | Low | Prospectively and well defined interventions. Outcomes determined after intervention. | Unknown | No information on protocol adherence. | Low | Prospective data collection with outcome data available for all participants. | Low | Prospective surveillance of outcome measure. Outcome measures unaffected by knowledge of intervention. Methods of outcome assessment comparable because cases identified from same databases. | Low | Outcomes correspond to standard incidence measures and there were no specific subset of analyses is reported. | Serious | This study has serious risk of bias in the domain of confounding. |
| Trijbels-Smeulders et al. 2006 | Moderate | Provide demographic data on EOGBS group. No control for potential confounding (including time). | Low | Both patient populations from same demographic. | Low | Prospectively and well defined interventions. Outcomes determined after intervention. | Unknown | No information on protocol adherence. | Low | Active surveillance. | Low | Retrospective outcome measurement, but active surveillance. Outcome measures unaffected by knowledge of intervention. Methods of outcome assessment comparable because cases identified from Dutch Paediatric Surveillance Unit. | Low | Outcomes correspond to standard resistance measures and there were no specific subset of analyses is reported. | Moderate | This study has moderate risk of bias in the domain of confounding. |
| Trijbels-Smeulders et al. 2007 | Serious | No demographic data total population and no control for potential confounding (including time). | Low | Both patient populations from same demographic. | Low | Prospectively and well defined interventions. Outcomes determined after intervention. | Unknown | No information on protocol adherence. | Moderate | Retrospective data collection. | Low | Retrospective outcome measurement, but active surveillance. Outcome measures unaffected by knowledge of intervention. Methods of outcome assessment comparable because cases identified from two surveillance database. | Low | Outcomes correspond to standard incidence measures and there were no specific subset of analyses is reported. | Serious | This study has serious risk of bias in the domain of confounding. |
| Uy et al. 2002 | Low | Provide demographic data on total population and no relevant confounding is present for IAP administration | Low | Both patient populations from same demographic. | Low | Prospectively and well defined interventions. Outcome is part of intervention. | Moderate | Adherence to risk factors was 59% in risk-based era | Moderate | Retrospective data collection. | Moderate | Retrospective outcome measurement, but outcomes likely standardised. Outcome measures unaffected by knowledge of intervention. Methods of outcome assessment comparable because cases identified from computerised neonatal database. | Low | Outcomes correspond to antibiotic administration measures and there were no specific subset of analyses is reported. | Moderate | This study has moderate risk of bias in the domains of protocol adherence, missing data and outcome measurement. |
| van den Hoogen et al. 2010 | Serious | No demographic data total population and no control for potential confounding (including time). | Low | Both patient populations from same demographic. | Low | Prospectively and well defined interventions. Outcomes determined after intervention. | Unknown | No information on protocol adherence. | Low | Prospective data collection with outcome data available for all participants. | Low | Prospective outcome measurement. Outcome measures unaffected by knowledge of intervention. Methods of outcome assessment comparable because cases identified from national registry. | Critical | Outcomes do not correspond to standard incidence measures because no total live births, that is why calculated by hand. | Critical | This study has critical risk of bias in the domain of reported results. |
| van Dyke et al. 2009 | Moderate | Provide demographic data on total population, but no control for potential confounders (including time). | Low | Both patient populations from same demographic. | Low | Prospectively and well defined interventions. Outcomes determined after intervention. | Low | Adherence ranged between 74-85% in the time periods | Moderate | Retrospective data collection. | Low | Retrospective outcome measurement, but active surveillance. Outcome measures unaffected by knowledge of intervention. Methods of outcome assessment comparable because cases identified from same surveillance database. | Low | Outcomes correspond to standard incidence measures and there were no specific subset of analyses is reported. | Moderate | This study has serious risk of bias in the domains of confounding and missing data. |
| Vergani et al. 2002 | Moderate | Provide demographic data on total population, but no control for potential confounding (including time). | Low | Both patient populations from same demographic. | Low | Prospectively and well defined interventions. Outcomes determined after intervention. | Unknown | No information on protocol adherence. | Moderate | Retrospective data collection. | Moderate | Retrospective outcome measurement, but outcome measurement likely standardised. Outcome measures unaffected by knowledge of intervention. Methods of outcome assessment comparable because cases identified from chart review. | Low | Outcomes correspond to standard incidence measures and there were no specific subset of analyses is reported. | Moderate | This study has moderate risk of bias in the domain of confounding, missing data and outcome measurement. |
| Wang et al. 2023 | Serious | No demographic data on total population and no control for potential confounding (including time). | Low | Both patient populations from same demographic. | Low | Prospectively and well defined interventions. Outcomes determined after intervention. | Unknown | No information on protocol adherence. | Moderate | Retrospective data collection. | Moderate | Retrospective outcome measurement, but outcome measurement likely standardised. Outcome measures unaffected by knowledge of intervention. Methods of outcome assessment comparable because cases identified from same laboratory database and reporting/clinical systems. | Low | Outcomes correspond to standard incidence measures and there were no specific subset of analyses is reported. | Serious | This study has serious risk of bias in the domain of confounding. |
| Wicker et al. 2019 | Serious | No demographic data on total population and no control for potential confounding (including time). | Low | Both patient populations from same demographic. | Moderate | Prospectively and well defined interventions. Outcomes determined after intervention. Antibiotic agent unknown | Unknown | No information on protocol adherence. | Low | Prospective data collection with outcome data available for all participants. | Low | Prospective active surveillance. Outcome measures unaffected by knowledge of intervention. Methods of outcome assessment comparable because cases identified with same method. | Low | Outcomes correspond to standard incidence measures and there were no specific subset of analyses is reported. | Serious | This study has serious risk of bias in the domain of confounding. |
| Youden et al. 2005 | Low | Provide demographic data on total population and no relevant confounding is present for IAP administration with concurrent controls | Low | Both patient populations from same demographic. | Low | Prospectively and well defined interventions. Outcomes is part of intervention. | Moderate | Adherence to screening was 77%. | Low | Prospective data collection with data available for 98% of population. | Low | Prospective outcome measurement. Outcome measures unaffected by knowledge of intervention. Methods of outcome assessment comparable because cases identified with same method. | Low | Outcomes correspond to standard incidence measures and there were no specific subset of analyses is reported. | Moderate | This study has moderate risk of bias in the domain of deviations from intended interventions. |
| Yücesoy et al. 2004 | Low | Provide demographic data on total population. Quasi-experimental study design lowers risk of confounding. All possible confounders are equally distributed and concurrent controls make time bias less likely. | Low | Both patient populations from same demographic. | Low | Prospectively and well defined interventions. Outcomes determined after intervention. | Unknown | No information on protocol adherence. | Low | Prospective data collection with outcome data available for all participants. | Low | Prospective outcome measurement. Outcome measures unaffected by knowledge of intervention. Methods of outcome assessment are the same for both groups, because of concurrent system. | Low | Outcomes correspond to standard incidence measures and there were no specific subset of analyses is reported. | Low | This study has low risk of bias in all the domains. |
