## Supplementary file 4 for "Intrapartum antibiotic prophylaxis to prevent Group B streptococcal infections in newborn infants: a systematic review and meta-analysis comparing various strategies"

### Forest plots for meta-analysis comparisons

Term EOGBS:

Forest plot, term EOGBS, any vs no strategy

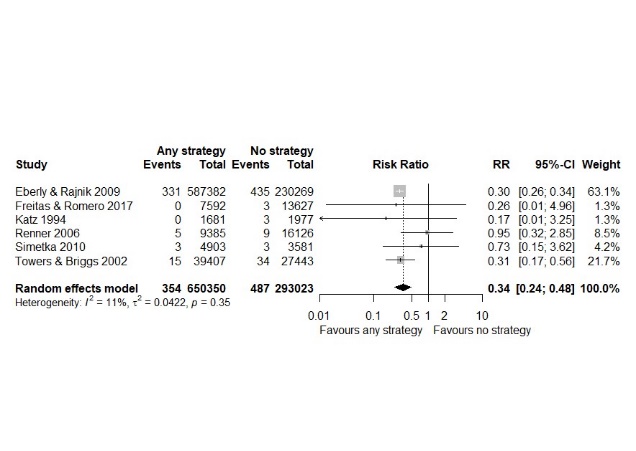

Forest plot, term EOGBS, risk vs no strategy

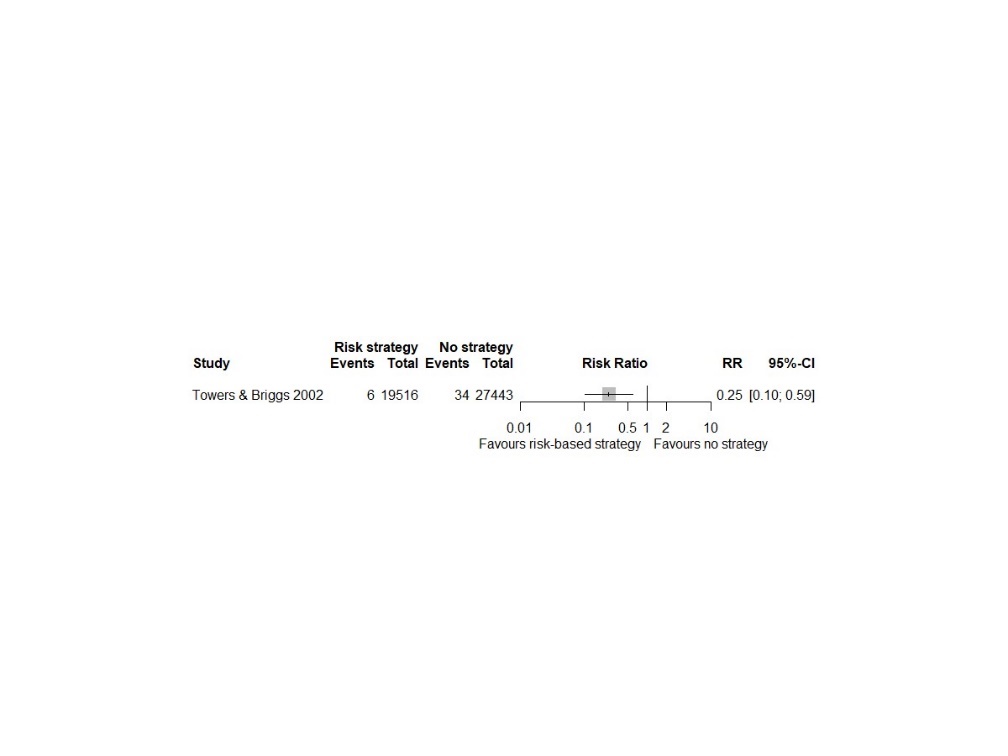

Forest plot, term EOGBS, universal vs no strategy

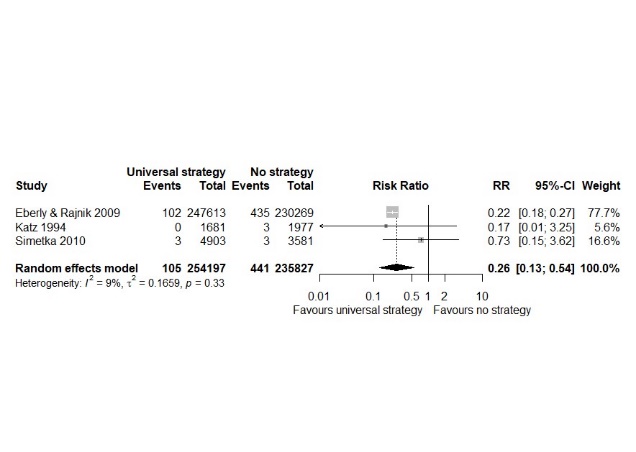

Forest plot, term EOGBS, universal vs risk strategies

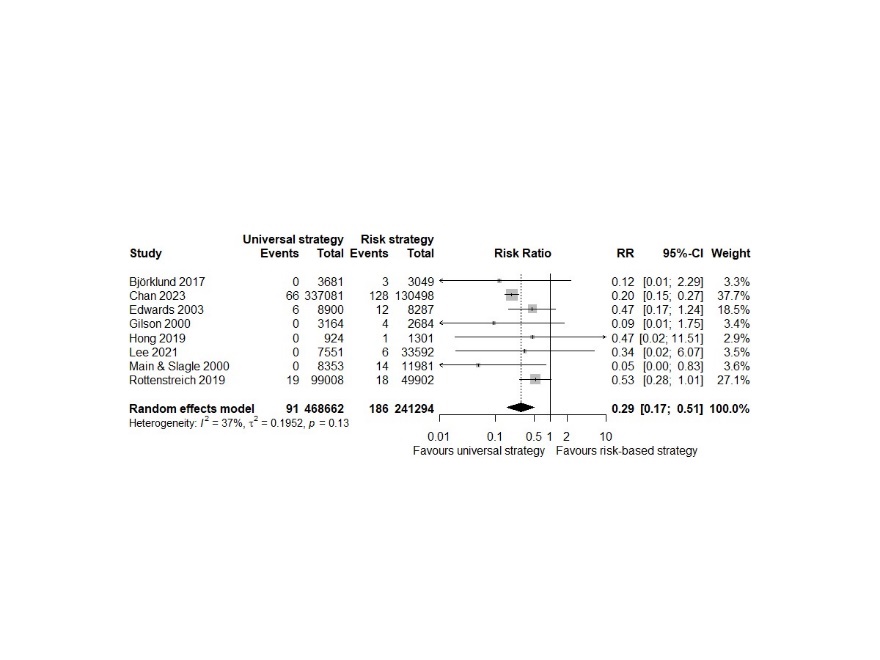

Timing of determination:

Forest plot, EOGBS, early vs late antenatal determination

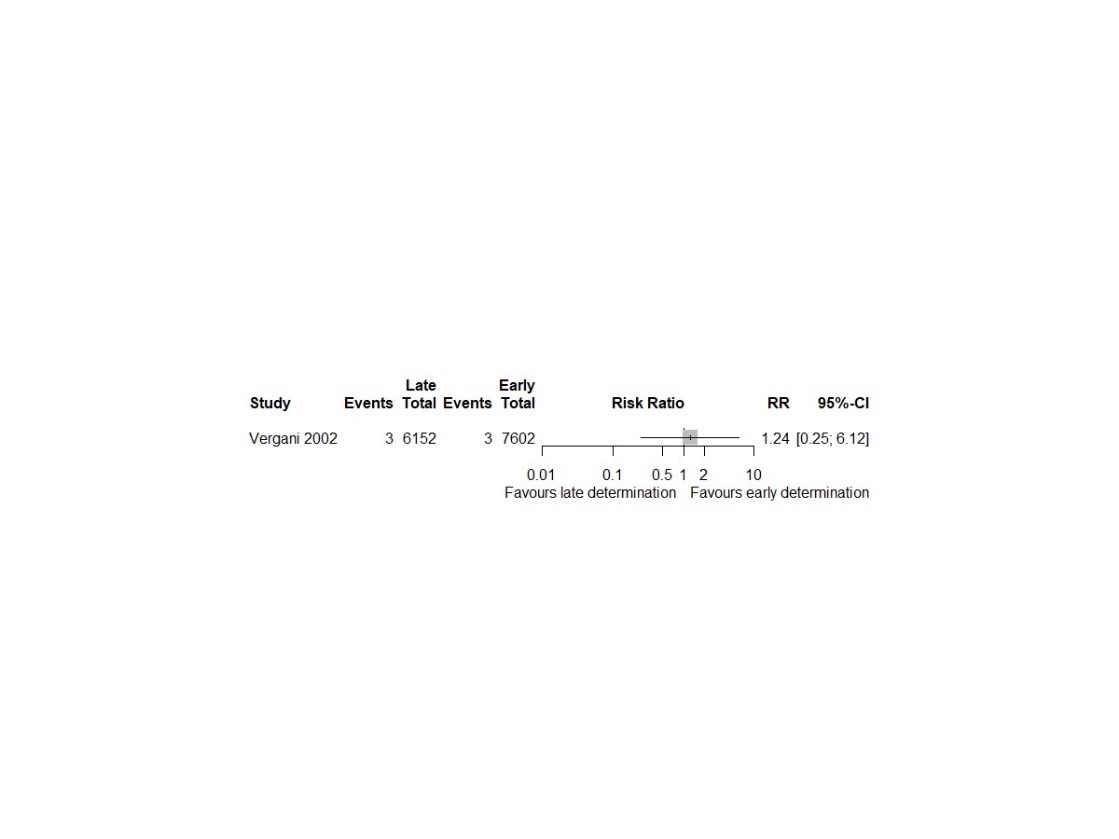

Sensitivity analysis (SA):

Forest plot, SA EOGBS, any vs no strategy

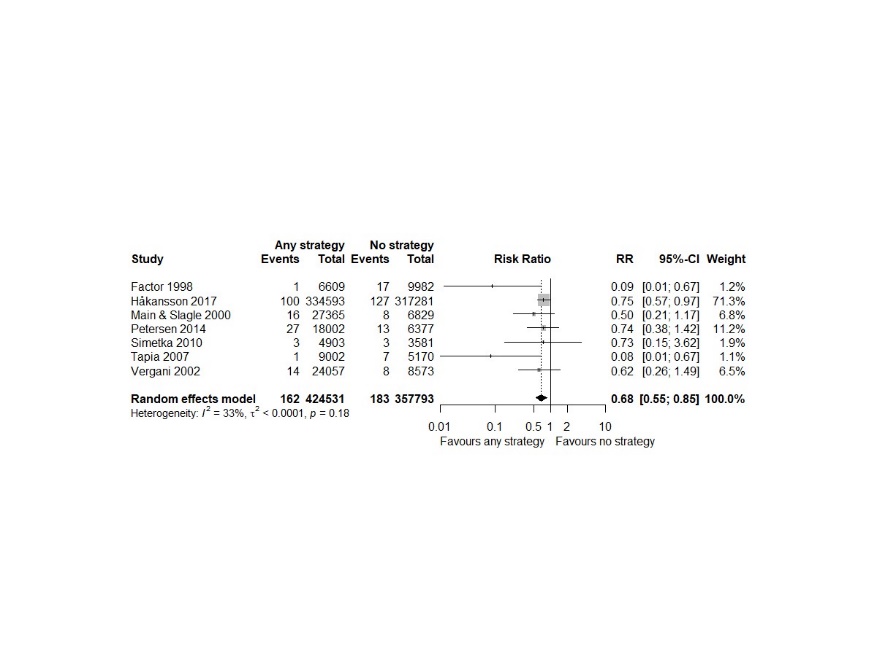

Forest plot, SA EOGBS, risk vs no strategy

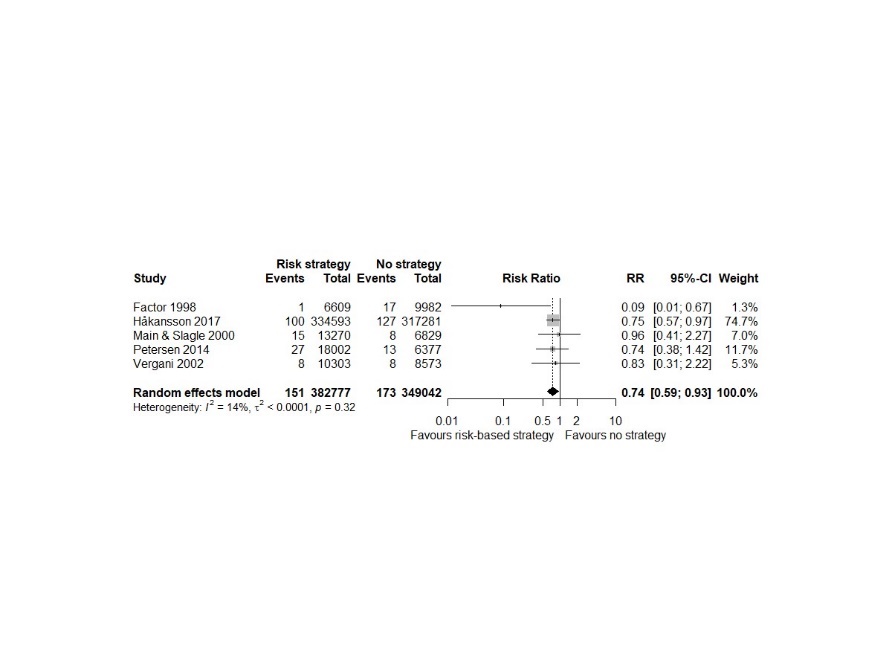

Forest plot, SA EOGBS, universal vs no strategy

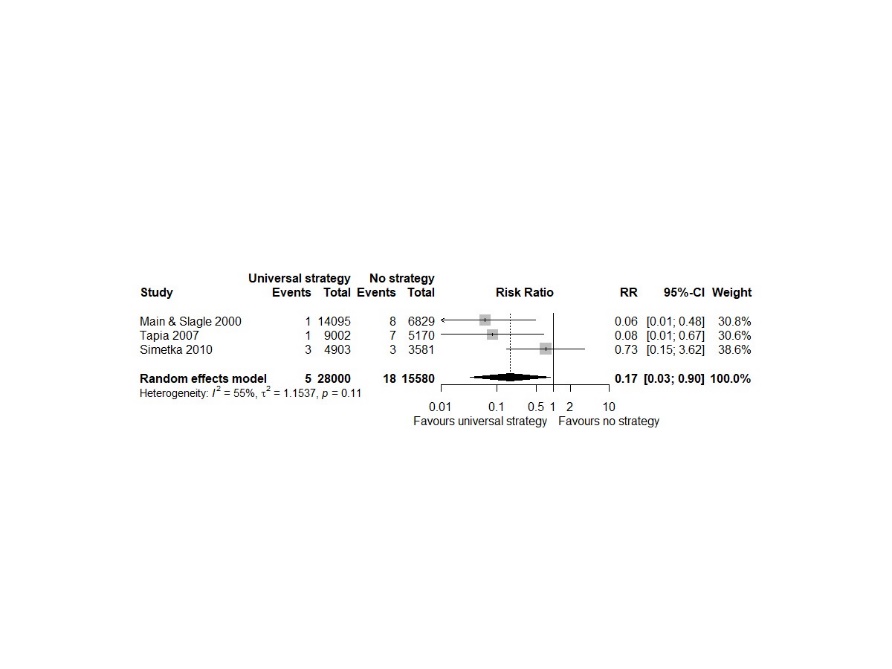

Forest plot, SA EOGBS, universal vs risk strategies

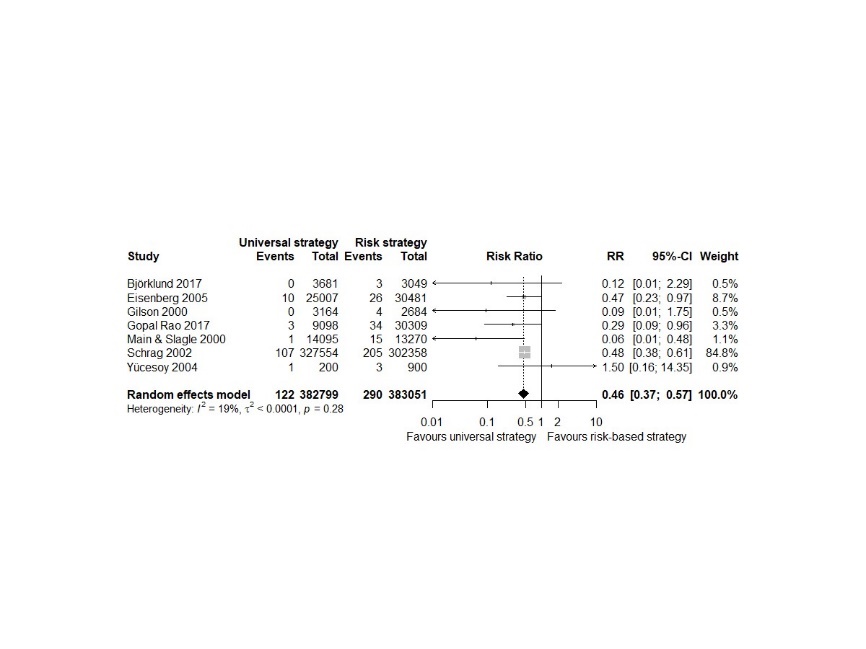

Timing of determination:

Forest plot, EOGBS, intrapartum vs antepartum determination

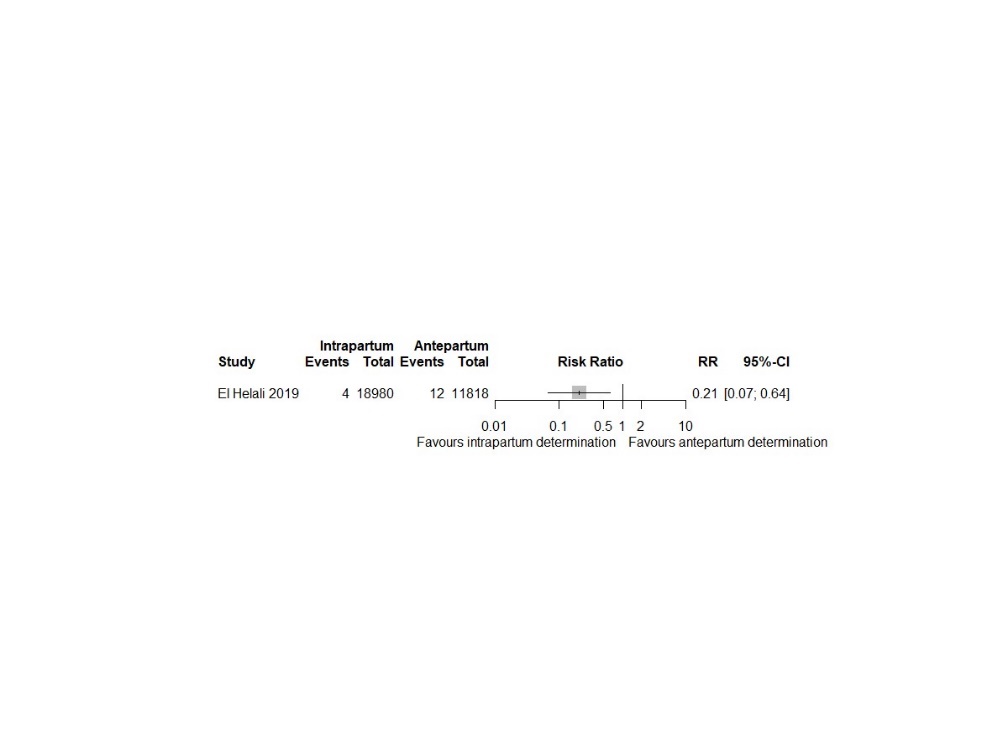

Non-GBS EOS:

Forest plot, Non-GBS EOS, any vs no strategy

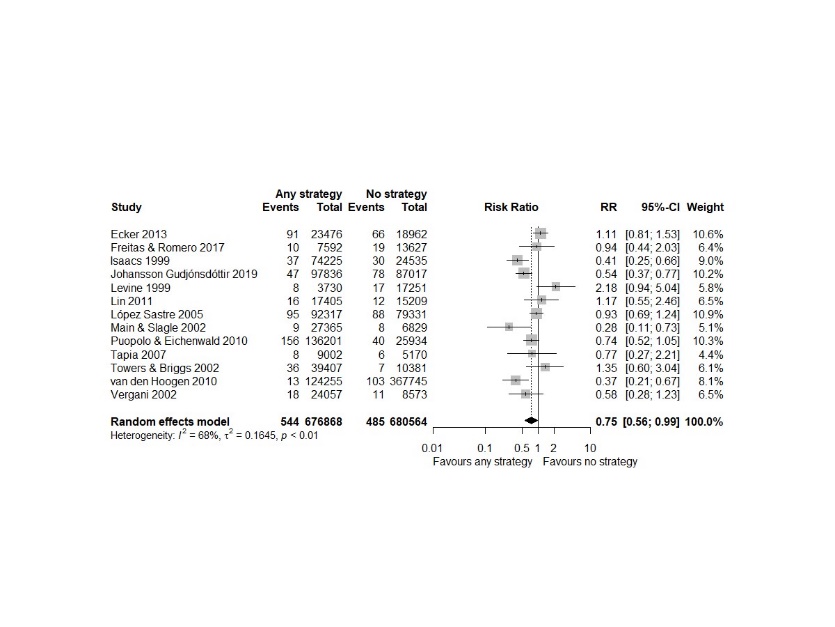

Forest plot, Non-GBS EOS, risk vs no strategy

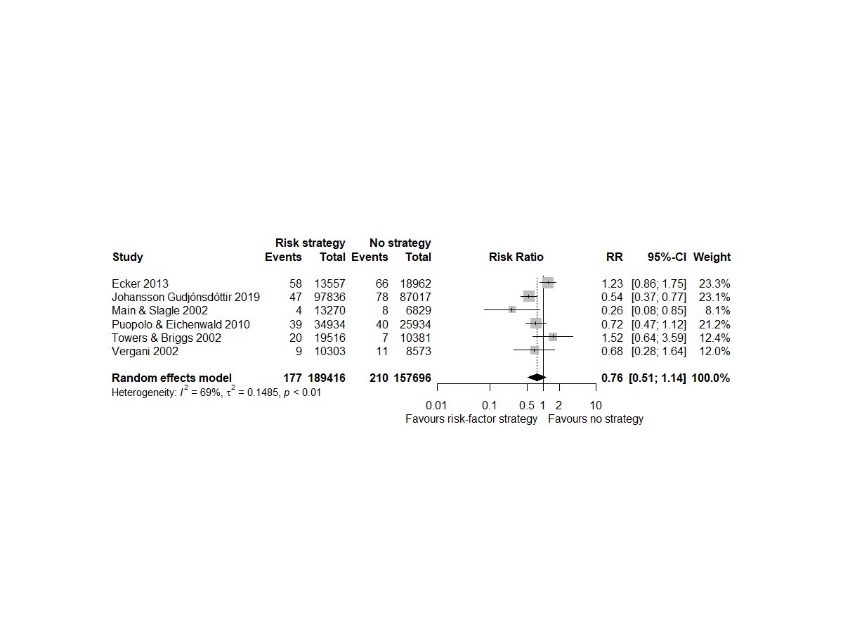

Forest plot, Non-GBS EOS, universal vs no strategy

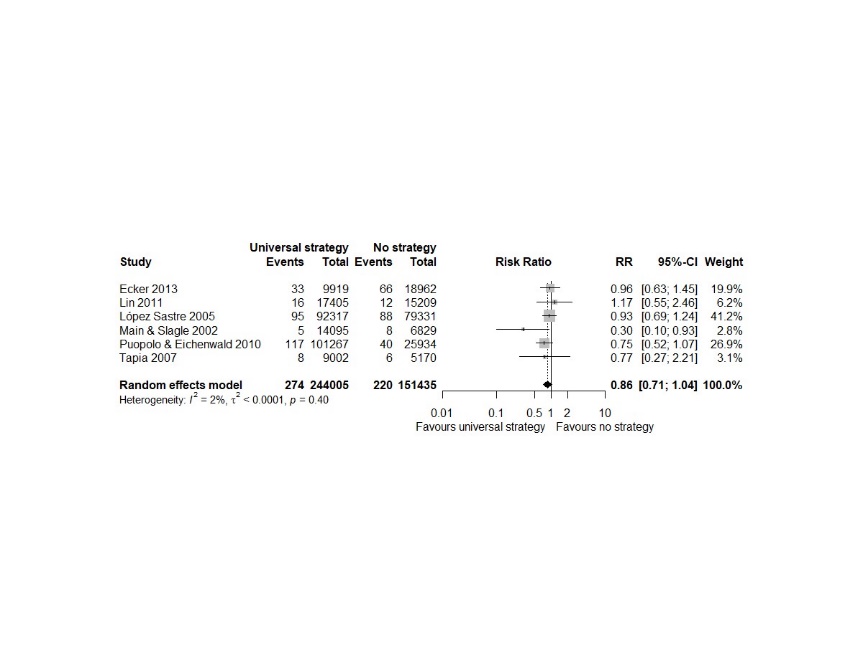

Forest plot, Non-GBS EOS, universal vs risk strategies

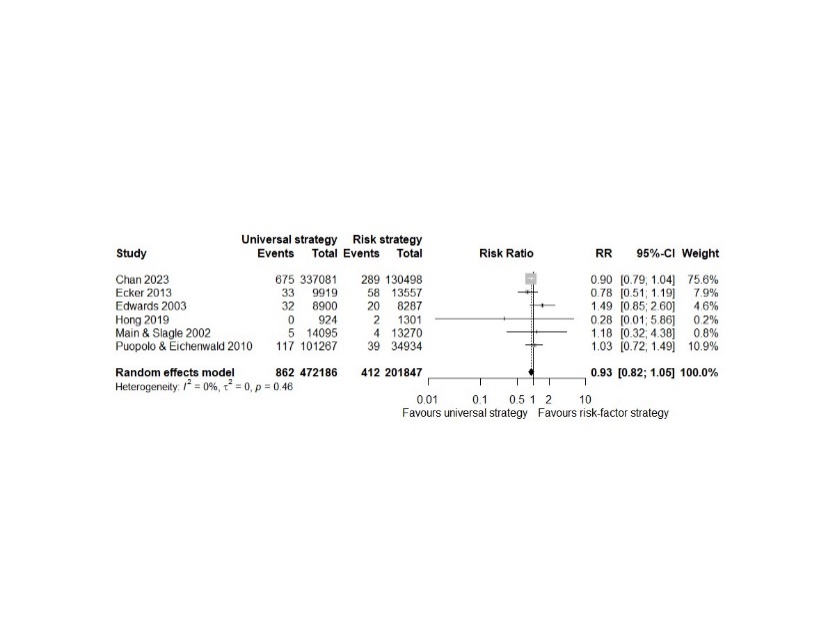

All EOS:

Forest plot, all EOS, any vs no strategy

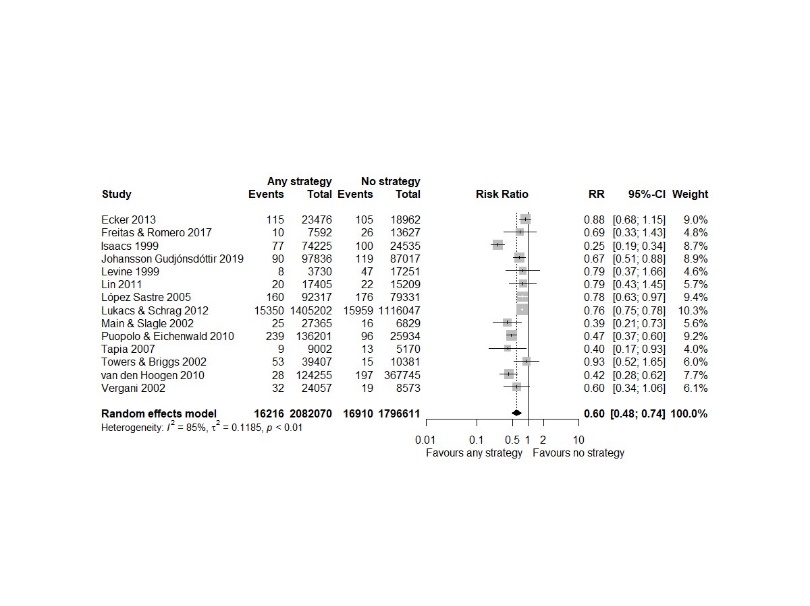

Forest plot, all EOS, risk vs no strategy

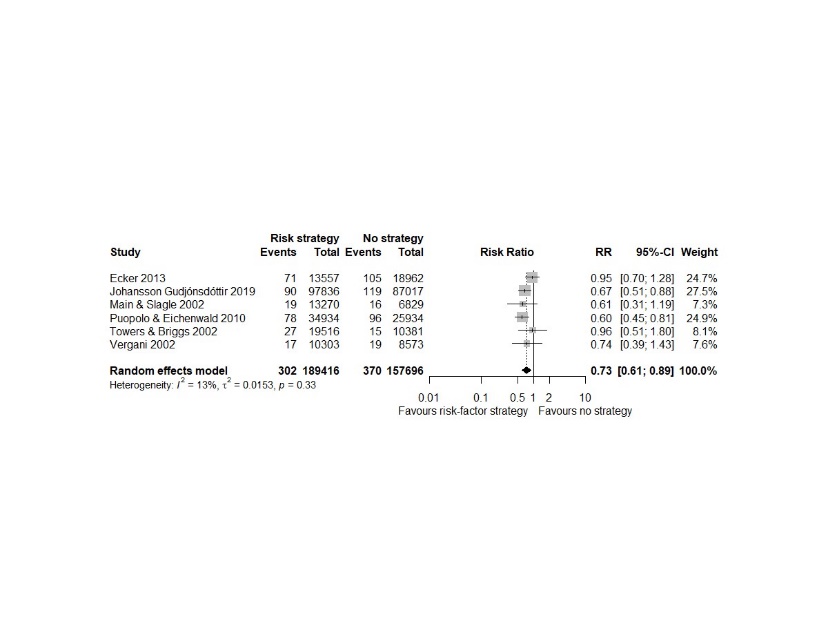

Forest plot, all EOS, universal vs no strategy

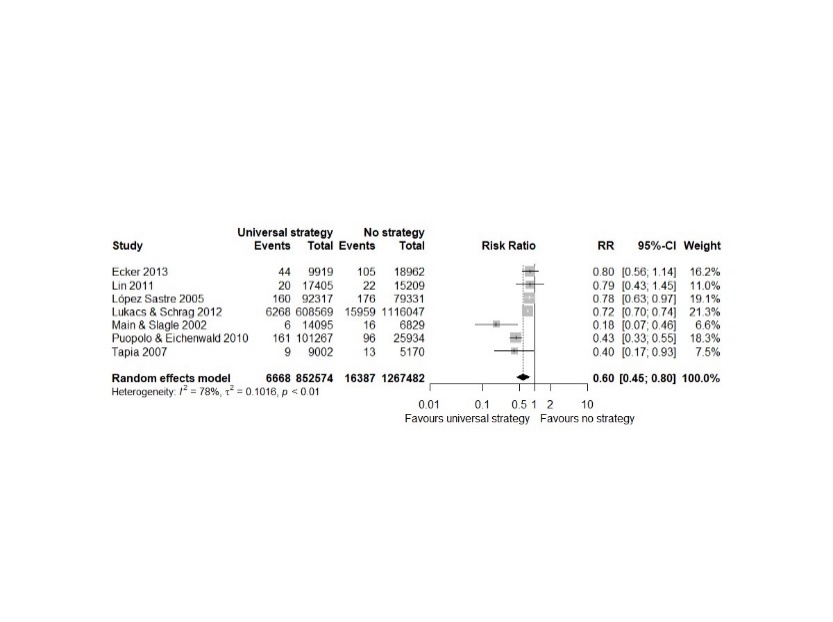

Forest plot, all EOS, universal vs risk strategies

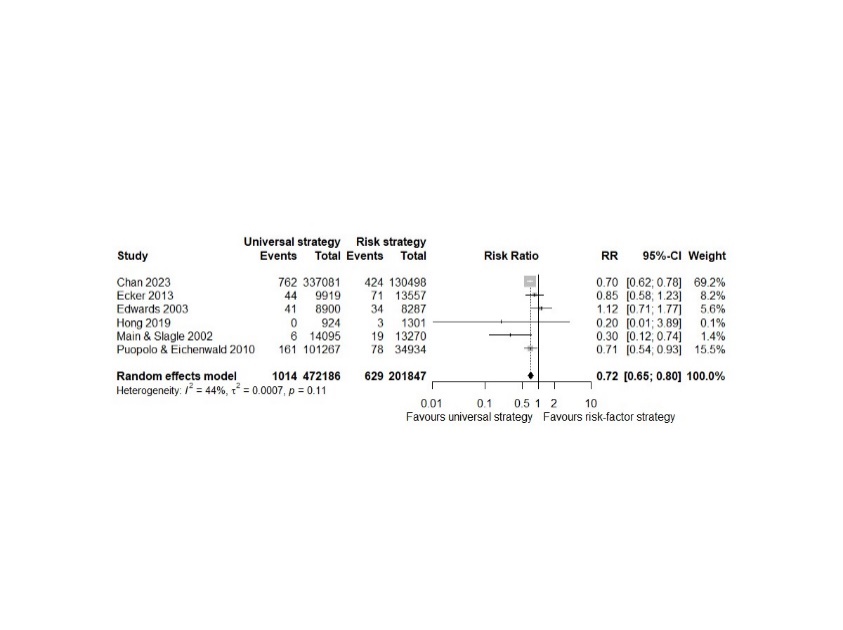

IAP:

Forest plot, IAP, universal vs risk strategies

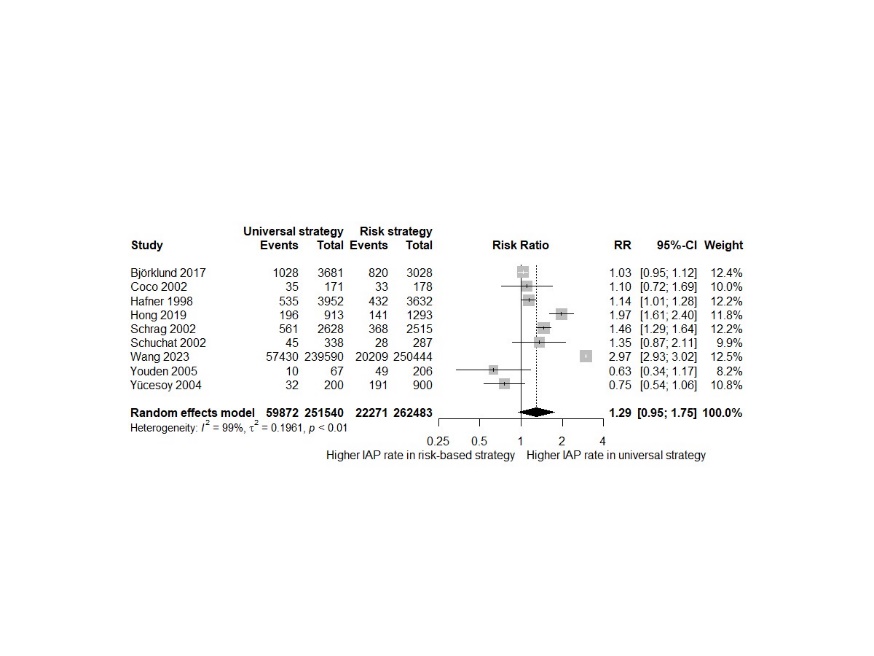
