## Supplementary file 5 for "Intrapartum antibiotic prophylaxis to prevent Group B streptococcal infections in newborn infants: a systematic review and meta-analysis comparing various strategies"

### Publication bias funnel plots for meta-analysis comparisons

Publication bias funnel plot, EOGBS, any vs no strategy

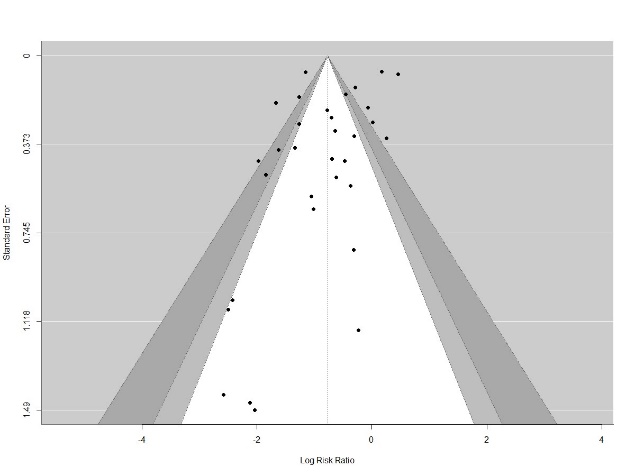

Publication bias funnel plot, EOGBS, risk vs no strategy

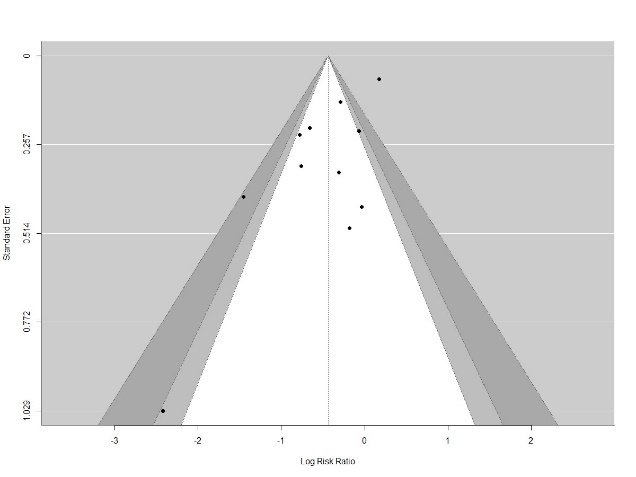

Publication bias funnel plot, EOGBS, universal vs no strategy

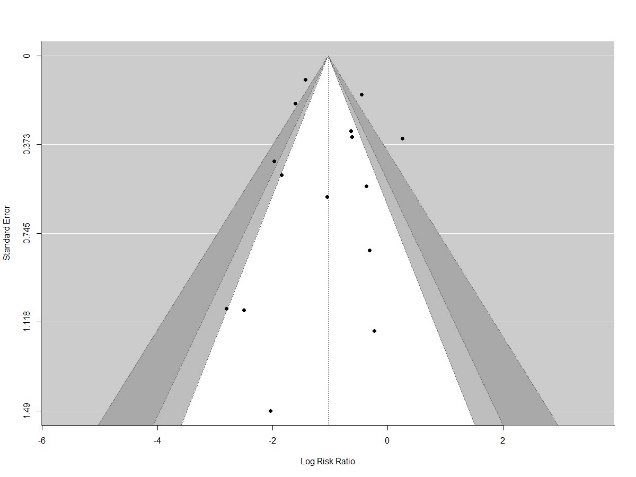

Publication bias funnel plot, EOGBS, universal vs risk strategies

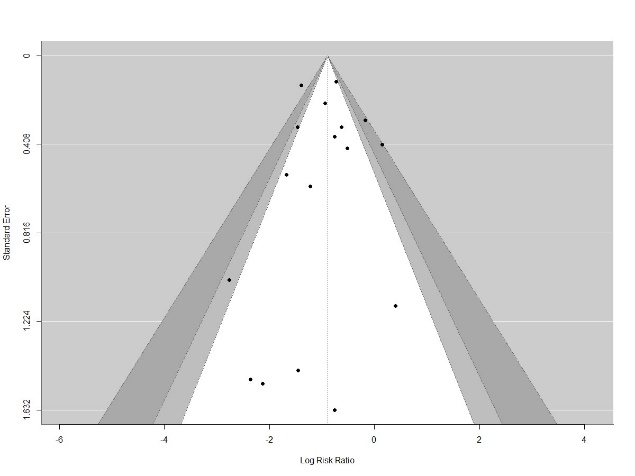

Publication bias funnel plot, Non-GBS EOS, any vs no strategy

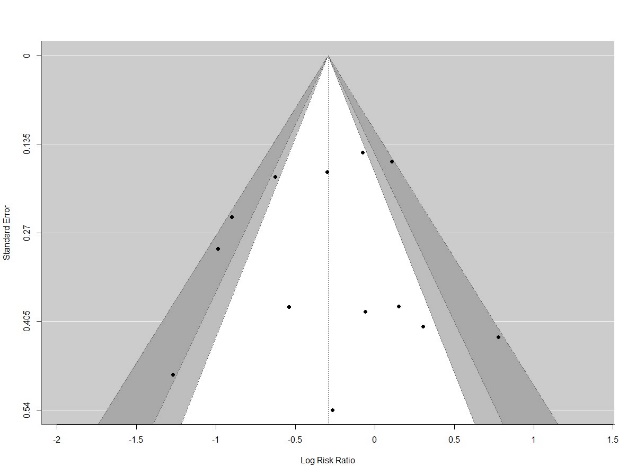

Publication bias funnel plot, All EOS, any vs no strategy

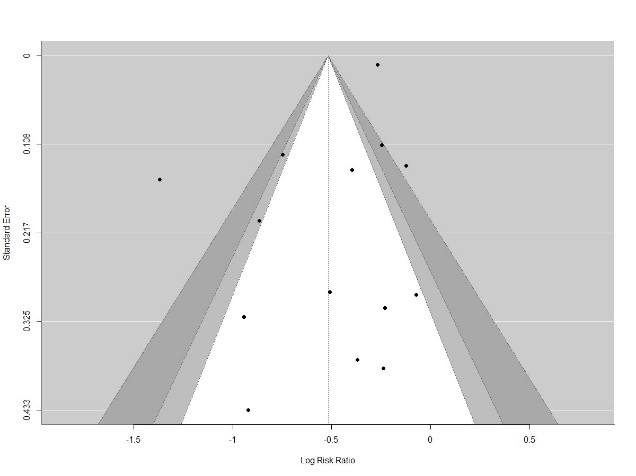
