## Supplementary file 6 for "Intrapartum antibiotic prophylaxis to prevent Group B streptococcal infections in newborn infants: a systematic review and meta-analysis comparing various strategies"

### GRADE Assesment for meta-analysis comparisons

### EOGBS infection (All incidences)

| **Certainty assessment** | | | | | | | **№ of patients** | | **Effect** | | **Certainty** |
| --- | --- | --- | --- | --- | --- | --- | --- | --- | --- | --- | --- |
| **№ of studies** | **Study design** | **Study limitation** | **Inconsistency** | **Indirectness** | **Imprecision** | **Publication bias** | **Any screening strategy** | **No screening strategy** | **Relative (95% CI)** | **Absolute (95% CI)** |  |
| 32 | Observational studies | Very serious^a^ | Serious^b^ | No indirectness^c^ | Not serious^d^ | Not serious^e^ | 18965252065  0.36/1000 live births | 1984/4789425  0.41/1000 live births | **RR 0.46** (0.36 to 0.60) | **0.71** (0.47-0.95) /1000 live births fewer cases | ⨁◯◯◯ Very low |
| a. Most of the studies were assessed to be at serious risk of bias using the ROBINS-I (2 downgrades due to most studies serious study limiation) b. Considerable statistical heterogeneity I^2^=93%, p<0.001, and some overlap in 95%-CI estimates of studies (1 downgrade due to inconsistent finding in the studies). c. Not applicable in this review, because all studies include report a direct comparison. d. The 95%-CI is relatively narrow and excludes a RR of 1.0. Due to large pooled sample size, grading down is unneccesary. e. No apparent publication bias in funnel plot. | | | | | | | | | | | |

| **Certainty assessment** | | | | | | | **№ of patients** | | **Effect** | | **Certainty** |
| --- | --- | --- | --- | --- | --- | --- | --- | --- | --- | --- | --- |
| **№ of studies** | **Study design** | **Study limitation** | **Inconsistency** | **Indirectness** | **Imprecision** | **Publication bias** | **Risk-factor based screening strategy** | **No screening strategy** | **Relative (95% CI)** | **Absolute (95% CI)** |  |
| 11 | Observational studies | Very serious^a^ | Serious^b^ | No indirectness^c^ | Not serious^d^ | Serious^e^ | 799/1589886  0.50/1000 live births | 784/1414837  0.55/1000 live births | **RR 0.65** (0.48 to 0.87) | **0.46** (0.13-0.80) /1000 live births fewer cases | ⨁◯◯◯ Very low |
| a. Most of the studies were assessed to be at serious risk of bias using the ROBINS-I. b. Considerable statistical heterogeneity I^2^=82%, p<0.001, and some overlap in 95%-CI estimates of studies. c. The 95%-CI is relatively narrow and excludes a RR of 1.0. Due to large pooled sample size, grading down is unneccesary. e. The funnel plot demontrates a high probability of publication bias. Egger’s test was singificant for funnnel plot asymmetry. | | | | | | | | | | | |

| **Certainty assessment** | | | | | | | **№ of patients** | | **Effect** | | **Certainty** |
| --- | --- | --- | --- | --- | --- | --- | --- | --- | --- | --- | --- |
| **№ of studies** | **Study design** | **Study limitation** | **Inconsistency** | **Indirectness** | **Imprecision** | **Publication bias** | **Universal screening strategy** | **No screening strategy** | **Relative (95% CI)** | **Absolute (95% CI)** |  |
| 15 | Observational studies | Very serious^a^ | Serious^b^ | No indirectness^c^ | Not serious^d^ | Not serious^e^ | 306/792079  0.39/1000 live births | 798/787094  1.01/1000 live births | **RR 0.37** (0.25 to 0.55) | **0.77** (0.42-1.12) /1000 live births fewer cases | ⨁◯◯◯ Very low |
| a. Most of the studies were assessed to be at serious risk of bias using the ROBINS-I. b. Considerable statistical heterogeneity I^2^=78%, p<0.001, and some overlap in 95%-CI estimates of studies. c. Not applicable in this review. d. The 95%-CI is relatively narrow and excludes a RR of 1.0. Due to large pooled sample size, grading down is unneccesary. e. No apparent publication bias in funnel plot. | | | | | | | | | | | |

| **Certainty assessment** | | | | | | | **№ of patients** | | **Effect** | | **Certainty** |
| --- | --- | --- | --- | --- | --- | --- | --- | --- | --- | --- | --- |
| **№ of studies** | **Study design** | **Study limitation** | **Inconsistency** | **Indirectness** | **Imprecision** | **Publication bias** | **Universal screening strategy** | **Risk-factor based screening strategy** | **Relative (95% CI)** | **Absolute (95% CI)** |  |
| 17 | Observational studies | Very serious^a^ | Not serious^b^ | No indirectness^c^ | Not serious^d^ | Not serious^e^ | 341/1092790  0.31/1000 live births | 577/713302  0.81/1000 live births | **RR 0.41** (0.30 to 0.55) | **0.49** (0.34-0.65) /1000 live births fewer cases | ⨁⨁◯◯ Low |
| a. Most of the studies were assessed to be at serious risk of bias using the ROBINS-I. b. Substantial statistical heterogeneity I^2^=60%, p<0.001, but considerable overlap in 95%-CI estimates of studies. c. Not applicable in this review. d. The 95%-CI is relatively narrow and excludes a RR of 1.0. Due to large pooled sample size, grading down is unneccesary. e. No apparent publication bias in funnel plot. | | | | | | | | | | | |

### EOGBS infection (Term incidences)

| **Certainty assessment** | | | | | | | **№ of patients** | | **Effect** | | **Certainty** |
| --- | --- | --- | --- | --- | --- | --- | --- | --- | --- | --- | --- |
| **№ of studies** | **Study design** | **Study limitation** | **Inconsistency** | **Indirectness** | **Imprecision** | **Publication bias** | **Any screening strategy** | **No screening strategy** | **Relative (95% CI)** | **Absolute (95% CI)** |  |
| 6 | Observational studies | Very serious^a^ | Not serious^b^ | No indirectness^c^ | Not serious^d^ | Unknown^e^ | 354/650350  0.54/1000 live births | 487/293023  1.66/1000 live births | **RR 0.34** (0.24 to 0.48) | **0.64** (0.10-1.17) /1000 live births fewer cases | ⨁⨁◯◯ Low |
| a. Most of the studies were assessed to be at serious risk of bias using the ROBINS-I b. Unimportant statistical heterogeneity I^2^=11%, p-=0.35, and complete overlap in 95%-CI estimates of studies. c. Not applicable in this review, because all studies include report a direct comparison. d. The 95%-CI is relatively narrow and excludes a RR of 1.0. Due to large pooled sample size, grading down is unneccesary. e. No apparent publication bias in funnel plot, but not enough studies to assess. | | | | | | | | | | | |

| **Certainty assessment** | | | | | | | **№ of patients** | | **Effect** | | **Certainty** |
| --- | --- | --- | --- | --- | --- | --- | --- | --- | --- | --- | --- |
| **№ of studies** | **Study design** | **Study limitation** | **Inconsistency** | **Indirectness** | **Imprecision** | **Publication bias** | **Risk-factor based screening strategy** | **No screening strategy** | **Relative (95% CI)** | **Absolute (95% CI)** |  |
| 1 | Observational study | Very serious^a^ | Not relevant^b^ | No indirectness^c^ | Not serious^d^ | Not relevant^e^ | 34/27443  0.30/1000 live births | 6/19516  1.24/1000 live births | **RR 0.25** (0.10 to 0.59) | **0.93** (0.45-1.41) /1000 live births fewer cases | ⨁⨁◯◯ Low |
| a. Study at serious risk of bias. b. Only one study c. Not applicable in this review, because all studies include report a direct comparison. d. The 95%-CI is relatively narrow and excludes a RR of 1.0. Although small sample size, no downgrade, because effect large enough. Only one study. | | | | | | | | | | | |

| **Certainty assessment** | | | | | | | **№ of patients** | | **Effect** | | **Certainty** |
| --- | --- | --- | --- | --- | --- | --- | --- | --- | --- | --- | --- |
| **№ of studies** | **Study design** | **Study limitation** | **Inconsistency** | **Indirectness** | **Imprecision** | **Publication bias** | **Universal screening strategy** | **No screening strategy** | **Relative (95% CI)** | **Absolute (95% CI)** |  |
| 3 | Observational studies | Very serious^a^ | Not serious^b^ | No indirectness^c^ | Not serious^d^ | Unknown^e^ | 105/254197  0.41/1000 live births | 441/235827  1.87/1000 live births | **RR 0.26** (0.13 to 0.54) | **1.11** (0.19-2.04) /1000 live births fewer cases | ⨁⨁◯◯ Low |
| a. Most of the studies were assessed to be at serious risk of bias using the ROBINS-I. b. Unimportant statistical heterogentiy I^2^=9%, p=0.33, and complete overlap in 95%-CI estimates of studies. c. Not applicable in this review. d. The 95%-CI is relatively narrow and excludes a RR of 1.0. Due to large pooled sample size, grading down is unneccesary. e. No apparent publication bias in funnel plot, but not enough studies to assess. | | | | | | | | | | | |

| **Certainty assessment** | | | | | | | **№ of patients** | | **Effect** | | **Certainty** |
| --- | --- | --- | --- | --- | --- | --- | --- | --- | --- | --- | --- |
| **№ of studies** | **Study design** | **Study limitation** | **Inconsistency** | **Indirectness** | **Imprecision** | **Publication bias** | **Universal screening strategy** | **Risk-factor based screening strategy** | **Relative (95% CI)** | **Absolute (95% CI)** |  |
| 8 | Observational studies | Very serious^a^ | Not serious^b^ | No indirectness^c^ | Not serious^d^ | Unknown^e^ | 91/468662  0.19/1000 live births | 186/241294  0.77/1000 live births | **RR 0.29** (0.17 to 0.51) | **0.61** (0.26-0.97) /1000 live births fewer cases | ⨁⨁◯◯ Low |
| a. Most of the studies were assessed to be at serious risk of bias using the ROBINS-I. b. Moderate statistical heterogeneity I^2^=37%, p=0.013, and almost complete overlap in 95%-CI estimates of studies. c. Not applicable in this review. d. The 95%-CI is relatively narrow and excludes a RR of 1.0. Due to large pooled sample size, grading down is unneccesary. e. No apparent publication bias in funnel plot, but not enough studies to assess. | | | | | | | | | | | |

### EOGBS infection (Sensitivity analysis)

| **Certainty assessment** | | | | | | | **№ of patients** | | **Effect** | | **Certainty** |
| --- | --- | --- | --- | --- | --- | --- | --- | --- | --- | --- | --- |
| **№ of studies** | **Study design** | **Study limitation** | **Inconsistency** | **Indirectness** | **Imprecision** | **Publication bias** | **Any screening strategy** | **No screening strategy** | **Relative (95% CI)** | **Absolute (95% CI)** |  |
| 7 | Observational studies | Serious^a^ | Not serious^b^ | No indirectness^c^ | Not serious^d^ | Unknown^e^ | 162/424531  0.38/1000 live births | 183/357793  0.51/1000 live births | **RR 0.68** (0.55 to 0.85) | **0.60** (0.12-1.08) /1000 live births fewer cases | ⨁⨁⨁◯ Moderate |
| a. Most of the studies were assessed to be at moderate risk of bias using the ROBINS-I. b. Moderate statistical heterogeneity I^2^=33%, p=0.18, and complete overlap in 95%-CI estimates of studies. c. Not applicable in this review. d. The 95%-CI is relatively narrow and excludes a RR of 1.0. Due to large sample size, grading down is unneccesary e. No apparent publication bias in funnel plot, but not enough studies to assess. | | | | | | | | | | | |

| **Certainty assessment** | | | | | | | **№ of patients** | | **Effect** | | **Certainty** |
| --- | --- | --- | --- | --- | --- | --- | --- | --- | --- | --- | --- |
| **№ of studies** | **Study design** | **Study limitation** | **Inconsistency** | **Indirectness** | **Imprecision** | **Publication bias** | **Risk-factor based screening strategy** | **No screening strategy** | **Relative (95% CI)** | **Absolute (95% CI)** |  |
| 5 | Observational studies | Serious^a^ | Not serious^b^ | No indirectness^c^ | Not serious^d^ | Unknown^e^ | 151/38277  0.39/1000 live births | 173/349042  0.50/1000 live births | **RR 0.74** (0.59 to 0.93) | **0.43** (-0.13-0.99) /1000 live births fewer cases | ⨁⨁⨁◯ Moderate |
| a. Most of the studies were assessed to be at moderate risk of bias using the ROBINS-I. b. Unimportant statistical heterogeneity I^2^=14%, p=0.32, and complete overlap in 95%-CI estimates of studies. c. Not applicable in this review. d. The 95%-CI is relatively narrow and excludes a RR of 1.0. Due to large sample size, grading down is unneccesary e. No apparent publication bias in funnel plot, but not enough studies to assess. | | | | | | | | | | | |

| **Certainty assessment** | | | | | | | **№ of patients** | | **Effect** | | **Certainty** |
| --- | --- | --- | --- | --- | --- | --- | --- | --- | --- | --- | --- |
| **№ of studies** | **Study design** | **Study limitation** | **Inconsistency** | **Indirectness** | **Imprecision** | **Publication bias** | **Universal screening strategy** | **No screening strategy** | **Relative (95% CI)** | **Absolute (95% CI)** |  |
| 3 | Observational studies | Serious^a^ | Not serious^b^ | No indirectness^c^ | Not serious^d^ | Unknown^e^ | 5/28000  0.18/1000 live births | 18/15580  1.16/1000 live births | **RR 0.17** (0.03 to 0.90) | **0.93** (0.36-1.52) /1000 live births fewer cases | ⨁⨁⨁◯ Moderate |
| a. Most of the studies were assessed to be at moderate risk of bias using the ROBINS-I. b. Substantial statistical heterogeneity I^2^=55%, p=0.11, but complete overlap in 95%-CI estimates of studies. c. Not applicable in this review. d. The 95%-CI is relatively narrow and excludes a RR of 1.0. Due to large sample size, grading down is unneccesary. e. No apparent publication bias in funnel plot, but not enough studies to assess. | | | | | | | | | | | |

| **Certainty assessment** | | | | | | | **№ of patients** | | **Effect** | | **Certainty** |
| --- | --- | --- | --- | --- | --- | --- | --- | --- | --- | --- | --- |
| **№ of studies** | **Study design** | **Study limitation** | **Inconsistency** | **Indirectness** | **Imprecision** | **Publication bias** | **Universal screening strategy** | **Risk-factor based screening strategy** | **Relative (95% CI)** | **Absolute (95% CI)** |  |
| 7 | Observational studies | Serious^a^ | Not serious^b^ | No indirectness^c^ | Not serious^d^ | Unknown^e^ | 122/382799  0.32/1000 live births | 290/383051  0.76/1000 live births | **RR 0.46** (0.47 to 0.57) | **0.64** (0.33-0.95) /1000 live births fewer cases | ⨁⨁⨁◯ Moderate |
| a. Most of the studies were assessed to be at moderate risk of bias using the ROBINS-I. b. Unimportant statistical heterogeneity I^2^=19%, p=0.28, and complete overlap in 95%-CI estimates of studies. c. Not applicable in this review. d. The 95%-CI is relatively narrow and excludes a RR of 1.0. Due to large sample size, grading down is unneccesary. e. No apparent publication bias in funnel plot, but not enough studies to assess. | | | | | | | | | | | |

### EOGBS timing of determination (early vs. late antenatal)

| **Certainty assessment** | | | | | | | **№ of patients** | | **Effect** | | **Certainty** |
| --- | --- | --- | --- | --- | --- | --- | --- | --- | --- | --- | --- |
| **№ of studies** | **Study design** | **Study limitation** | **Inconsistency** | **Indirectness** | **Imprecision** | **Publication bias** | **Late antenatal universal screening strategy** | **Early antenatal universal screening strategy** | **Relative (95% CI)** | **Absolute (95% CI)** |  |
| 1 | Observational study | Very serious^a^ | Not relevant^b^ | Serious^c^ | Serious^d^ | Not relevant^e^ | 3/6152  0.49/1000 live births | 3/7602  0.39/1000 live births | **RR 1.24** (0.25 to 6.12) | **0.09** (-0.62-0.80) /1000 live births **more** cases | ⨁◯◯◯ Very low |
| a. Study at serious risk of bias. b. Only one study c. A combination strategy was used instead of a universal strategy. d. The 95%-CI is wide and includes a RR of 1.0. Sample size is not sufficiently large to detect a precise effect e. Only one study. | | | | | | | | | | | |

### EOGBS timing of determination (intrapartum vs. antepartum)

| **Certainty assessment** | | | | | | | **№ of patients** | | **Effect** | | **Certainty** |
| --- | --- | --- | --- | --- | --- | --- | --- | --- | --- | --- | --- |
| **№ of studies** | **Study design** | **Study limitation** | **Inconsistency** | **Indirectness** | **Imprecision** | **Publication bias** | **Intrapartum universal screening strategy** | **Antepartum universal screening strategy** | **Relative (95% CI)** | **Absolute (95% CI)** |  |
| 1 | Observational study | Serious^a^ | Not relevant^b^ | No indirectness^c^ | Not serious^d^ | Not relevant^e^ | 4/18980  0.21/1000 live births | 12/11818  1.02/1000 live births | **RR 0.21** (0.07 to 0.64) | **0.80** (0.19-1.41) /1000 live births fewer cases | ⨁⨁⨁◯ Moderate |
| a. Study at moderate risk of bias. b. Only one study c. Not applicable in this review, because all studies include report a direct comparison. d. The 95%-CI is relatively narrow and excludes a RR of 1.0. Although small sample size, no downgrade, because effect large enough. e. Only one study. | | | | | | | | | | | |

### Non-GBS EOS

| **Certainty assessment** | | | | | | | **№ of patients** | | **Effect** | | **Certainty** |
| --- | --- | --- | --- | --- | --- | --- | --- | --- | --- | --- | --- |
| **№ of studies** | **Study design** | **Study limitation (risk of bias)** | **Inconsistency** | **Indirectness** | **Imprecision** | **Publication bias** | **Any screening strategy** | **No screening strategy** | **Relative (95% CI)** | **Absolute (95% CI)** |  |
| 13 | Observational studies | Very serious^a^ | Not serious^b^ | No indirectness^c^ | Not serious^d^ | Not serious^e^ | 544/676868  0.80/1000 live births | 485/680564  0.71/1000 live births | **RR 0.75** (0.56 to 0.99) | **0.23** (0.049-0.42) /1000 live births fewer cases | ⨁⨁◯◯ Low |
| a. Most of the studies were assessed to be at serious risk of bias using the ROBINS-I. b. Substantial statistical heterogeneity I^2^=68%, p<0.001, and some overlap in 95%-CI estimates of studies. c. Not applicable in this review. d. The 95%-CI is relatively narrow and excludes a RR of 1.0. Due to large sample size, grading down is unneccesary e. No apparent publication bias in funnel plot. | | | | | | | | | | | |

| **Certainty assessment** | | | | | | | **№ of patients** | | **Effect** | | **Certainty** |
| --- | --- | --- | --- | --- | --- | --- | --- | --- | --- | --- | --- |
| **№ of studies** | **Study design** | **Study limitation** | **Inconsistency** | **Indirectness** | **Imprecision** | **Publication bias** | **Risk-factor based screening strategy** | **No screening strategy** | **Relative (95% CI)** | **Absolute (95% CI)** |  |
| 6 | Observational studies | Very serious^a^ | Not serious^b^ | No indirectness^c^ | Serious^d^ | Unknown^e^ | 177/189416  0.93/1000 live births | 210/157696  1.33/1000 live births | **RR 0.76** (0.51 to 1.14) | **0.27** (-0.099-0.64) /1000 live births fewer cases | ⨁◯◯◯ Very low |
| a. Most of the studies were assessed to be at serious risk of bias using the ROBINS-I. b. Substantial statistical heterogeneity I^2^=69%, p=0.006, but considerable overlap in 95%-CI estimates of studies. c. Not applicable in this review. d. The 95%-CI is wide and includes a RR of 1.0. Sample size is not sufficiently large to detect a precise effect. e. No apparent publication bias in funnel plot, but not enough studies to assess. | | | | | | | | | | | |

| **Certainty assessment** | | | | | | | **№ of patients** | | **Effect** | | **Certainty** |
| --- | --- | --- | --- | --- | --- | --- | --- | --- | --- | --- | --- |
| **№ of studies** | **Study design** | **Study limitation** | **Inconsistency** | **Indirectness** | **Imprecision** | **Publication bias** | **Universal screening strategy** | **No screening strategy** | **Relative (95% CI)** | **Absolute (95% CI)** |  |
| 6 | Observational studies | Very serious^a^ | Not serious^b^ | No indirectness^c^ | Serious^d^ | Unknown^e^ | 274/244005  1.12/1000 live births | 220/151435  1.45/1000 live births | **RR 0.86** (0.72 to 1.04) | **0.17** (-0.056-0.40) /1000 live births fewer cases | ⨁◯◯◯ Very low |
| a. Most of the studies were assessed to be at serious risk of bias using the ROBINS-I. b. Unimportant statistical heterogeneity I^2^=2%, p=0.40, and complete overlap in 95%-CI estimates of studies. c. Not applicable in this review. d. The 95%-CI is wide and includes a RR of 1.0. Sample size is not sufficiently large to detect a precise effect. e. No apparent publication bias in funnel plot, but not enough studies to assess. | | | | | | | | | | | |

| **Certainty assessment** | | | | | | | **№ of patients** | | **Effect** | | **Certainty** |
| --- | --- | --- | --- | --- | --- | --- | --- | --- | --- | --- | --- |
| **№ of studies** | **Study design** | **Study limitation** | **Inconsistency** | **Indirectness** | **Imprecision** | **Publication bias** | **Universal screening strategy** | **Risk-factor based screening strategy** | **Relative (95% CI)** | **Absolute (95% CI)** |  |
| 6 | Observational studies | Very serious^a^ | Not serious^b^ | No indirectness^c^ | Serious^d^ | Unknown^e^ | 862/472186  1.83/1000 live births | 412/201847  2.04/1000 live births | **RR 0.93** (0.82 to 1.05) | **0.068** (-0.21-0.35) /1000 live births fewer cases | ⨁◯◯◯ Very low |
| a. Most of the studies were assessed to be at serious risk of bias using the ROBINS-I.. b. Unimportant statistical heterogeneity I^2^=0%, p=0.46, and complete overlap in 95%-CI estimates of studies. c. Not applicable in this review. d. The 95%-CI is wide and includes a RR of 1.0. Sample size is not sufficiently large to detect a precise effect. e. No apparent publication bias in funnel plot, but not enough studies to assess. | | | | | | | | | | | |

### All EOS

| **Certainty assessment** | | | | | | | **№ of patients** | | **Effect** | | **Certainty** |
| --- | --- | --- | --- | --- | --- | --- | --- | --- | --- | --- | --- |
| **№ of studies** | **Study design** | **Study limitation** | **Inconsistency** | **Indirectness** | **Imprecision** | **Publication bias** | **Any screening strategy** | **No screening strategy** | **Relative (95% CI)** | **Absolute (95% CI)** |  |
| 14 | Observational studies | Very serious^a^ | Serious^b^ | No indirectness^c^ | Not serious^d^ | Not serious^e^ | 16216/5163256  7.79/1000 live births | 16910/1796611  9.41/1000 live birthss | **RR 0.60** (0.48 to 0.74) | **1.13** (0.53-1.74) /1000 live births fewer cases | ⨁◯◯◯ Very low |
| a. Most of the studies were assessed to be at serious risk of bias using the ROBINS-I. b. Considerable statistical heterogeneity I^2^=85%, p<0.001, and some overlap in 95%-CI estimates of studies. c. Not applicable in this review. d. The 95%-CI is relatively narrow and excludes a RR of 1.0. Due to large sample size, grading down is unneccesary e. No apparent publication bias in funnel plot, but not enough studies to assess. | | | | | | | | | | | |

| **Certainty assessment** | | | | | | | **№ of patients** | | **Effect** | | **Certainty** |
| --- | --- | --- | --- | --- | --- | --- | --- | --- | --- | --- | --- |
| **№ of studies** | **Study design** | **Study limitation** | **Inconsistency** | **Indirectness** | **Imprecision** | **Publication bias** | **Risk-factor based screening strategy** | **No screening strategy** | **Relative (95% CI)** | **Absolute (95% CI)** |  |
| 6 | Observational studies | Very serious^a^ | Not serious^b^ | No indirectness^c^ | Not serious^d^ | Unknown^e^ | 302/189416  1.59/1000 live births | 370/157696  2.35/1000 live births | **RR 0.73** (0.61 to 0.89) | **0.57** (0.21-0.94) /1000 live births fewer cases | ⨁⨁◯◯ Low |
| a. Most of the studies were assessed to be at serious risk of bias using the ROBINS-I. b. Moderate statistical heterogeneity I^2^=33%, p=0.18, and complete overlap in 95%-CI estimates of studies. c. Not applicable in this review. d. The 95%-CI is relatively narrow and excludes a RR of 1.0. Due to large sample size, grading down is unneccesary e. No apparent publication bias in funnel plot, but not enough studies to assess. | | | | | | | | | | | |

| **Certainty assessment** | | | | | | | **№ of patients** | | **Effect** | | **Certainty** |
| --- | --- | --- | --- | --- | --- | --- | --- | --- | --- | --- | --- |
| **№ of studies** | **Study design** | **Study limitation** | **Inconsistency** | **Indirectness** | **Imprecision** | **Publication bias** | **Universal screening strategy** | **No screening strategy** | **Relative (95% CI)** | **Absolute (95% CI)** |  |
| 7 | Observational studies | Very serious^a^ | Serious^b^ | No indirectness^c^ | Not serious^d^ | Unknown^e^ | 6668/852574  7.82/1000 live births | 16387/1267482  12.93/1000 live births | **RR 0.60** (0.45 to 0.80) | **1.67** (0.61-2.72) /1000 live births fewer cases | ⨁◯◯◯ Very low |
| a. Most of the studies were assessed to be at serious risk of bias using the ROBINS-I. b. Considerable statistical heterogeneity I^2^=78%, p<0.001, and some overlap in 95%-CI estimates of studies. c. Not applicable in this review. d. The 95%-CI is relatively narrow and excludes a RR of 1.0. Due to large sample size, grading down is unneccesary e. No apparent publication bias in funnel plot, but not enough studies to assess. | | | | | | | | | | | |

| **Certainty assessment** | | | | | | | **№ of patients** | | **Effect** | | **Certainty** |
| --- | --- | --- | --- | --- | --- | --- | --- | --- | --- | --- | --- |
| **№ of studies** | **Study design** | **Study limitation** | **Inconsistency** | **Indirectness** | **Imprecision** | **Publication bias** | **Universal screening strategy** | **Risk-factor based screening strategy** | **Relative (95% CI)** | **Absolute (95% CI)** |  |
| 6 | Observational studies | Very serious^a^ | Not serious^b^ | No indirectness^c^ | Not serious^d^ | Unknown^e^ | 1014/472186  2.15/1000 live births | 629/201847  3.12/1000 live births | **RR 0.46** (0.36 to 0.60) | **0.88** (0.59-1.17) /1000 live births fewer cases | ⨁⨁◯◯ Low |
| a. Most of the studies were assessed to be at serious risk of bias using the ROBINS-I. b. Moderate statistical heterogeneity I^2^=44%, p<=0.11, and complete overlap in 95%-CI estimates of studies. c. Not applicable in this review. d. The 95%-CI is relatively narrow and excludes a RR of 1.0. Due to large sample size, grading down is unneccesary e. No apparent publication bias in funnel plot, but not enough studies to assess. | | | | | | | | | | | |

### IAP administration

| **Certainty assessment** | | | | | | | **№ of patients** | | **Effect** | | **Certainty** |
| --- | --- | --- | --- | --- | --- | --- | --- | --- | --- | --- | --- |
| **№ of studies** | **Study design** | **Study limitation** | **Inconsistency** | **Indirectness** | **Imprecision** | **Publication bias** | **Universal screening strategy** | **Risk-factor based screening strategy** | **Relative (95% CI)** | **Absolute (95% CI)** |  |
| 9 | Observational studies | Serious^a^ | Serious^b^ | No indirectness^c^ | Serious^d^ | Unknown^e^ | 59872/251540  23.8% | 22271/262483  8.5% | **RR 1.29** (0.95 to 1.75) | **-3.7** (-8.5- 1.1) % of pregnant women receiving IAP | ⨁◯◯◯ Very low |
| a. Most of the studies were assessed to be at moderate risk of bias using the ROBINS-I with regard to IAP administration. b. Considerable statistical heterogeneity I^2^=99%, p<0.001, and some overlap in 95%-CI estimates of studies. c. Not applicable in this review. d. The 95%-CI is wide and includes a RR of 1.0. Sample size is not sufficiently large to detect a precise effect. e. Appearace of publication bias in funnel plot, but not enough studies to assess. | | | | | | | | | | | |
